# Paving the way for human vaccination against Rift Valley fever virus: A systematic literature review of RVFV epidemiology from 1999 to 2021

**DOI:** 10.1101/2021.09.29.21264307

**Authors:** Keli N. Gerken, A. Desirée LaBeaud, Henshaw Mandi, Maïna L’Azou Jackson, J. Gabrielle Breugelmans, Charles H. King

## Abstract

**Background:** Rift Valley fever virus (RVFV) is a lethal threat to humans and livestock in many parts of Africa, the Arabian Peninsula, and the Indian Ocean. This systematic review’s objective was to consolidate understanding of RVFV epidemiology during 1999-2021 and highlight knowledge gaps relevant to plans for human vaccine trials.

**Methodology/Principal Findings:** The review is registered with PROSPERO (CRD42020221622). Reports of RVFV infection or exposure among humans, animals, and/or vectors in Africa, the Arabian Peninsula, and the Indian Ocean during the period January 1999 to June 2021 were eligible for inclusion. Online databases were searched for publications, and supplemental materials were recovered from official reports and research colleagues. Exposures were classified into five groups: 1) acute human RVF cases, 2) acute animal cases, 3) human RVFV sero-surveys, 4) animal sero-surveys, and 5) arthropod infections. Human risk factors, circulating RVFV lineages, and surveillance methods were also tabulated. In meta-analysis of risks, summary odds ratios were computed using random-effects modeling. 1104 unique human or animal RVFV transmission events were reported in 39 countries during 1999-2021. Outbreaks among humans or animals occurred at rates of 5.8/year and 12.4/year, respectively, with Mauritania, Madagascar, Kenya, South Africa, and Sudan having the most human outbreak years. Men had greater odds of RVFV infection than women, and animal contact, butchering, milking, and handling aborted material were significantly associated with greater odds of exposure. Animal infection risk was linked to location, proximity to water, and exposure to other herds or wildlife. RVFV was detected in a variety of mosquito vectors during interepidemic periods, confirming ongoing transmission.

**Conclusions/Significance:** With broad variability in surveillance, case finding, survey design, and RVFV case confirmation, combined with uncertainty about populations-at-risk, there were inconsistent results from location to location. However, it was evident that RVFV transmission is expanding its range and frequency. Gaps assessment indicated the need to harmonize human and animal surveillance and improve diagnostics and genotyping. Given the frequency of RVFV outbreaks, human vaccination has strong potential to mitigate the impact of this now widely endemic disease.

**Author Summary:** Rift Valley fever virus (RVFV) is a globally important mosquito-transmitted zoonosis that is also directly transmissible via aerosolization of body fluids from infected animals. RVFV outbreaks cause mass mortality of young livestock and pregnancy losses in both humans and animals. Severe human cases also result in hemorrhagic fever, encephalitis, and death. Loss of livestock additionally threatens the livelihood of people who depend on animals for income and food. In endemic areas, initiation of RVFV outbreaks is connected to weather events that cause excess rainfall, leading to flooding and subsequent mosquito blooms. However, the natural cycle of RVFV transmission is complex, requiring congregation of susceptible mammalian hosts and mosquito vectors in suitable environments. Several human vaccine candidates are in different stages of development, but none are yet licensed for use in human populations. In this systematic review, we assessed the 1999-2021 frequency and distribution of RVFV outbreaks among humans, animals, and vectors to identify potential locations and population targets for a human RVFV vaccine efficacy trial. It focuses on current understanding of RVFV epidemiology and the identification of gaps that pose critical barriers to controlling expansion of RVFV and implementing new protective measures including human vaccination.

## Introduction

Rift Valley fever virus (RVFV) remains an important emerging arboviral pathogen due to its recent geographic spread and its combined disease and financial impacts on vulnerable human populations [1]. Specifically, RVFV is listed as a priority pathogen by the World Health Organization (WHO), the Food and Agriculture Organization of the United Nations (FAO), and the U.S. Centers for Disease Control and Prevention (CDC) because of its ability to cause life-threatening hemorrhagic fever and encephalitis in humans [2, 3], its epidemic potential to cause severe harm to livestock [4, 5], and its potential for non-vector aerosol spread during epizootics and epidemics [6, 7]. The RVF virus is a member of the *Phlebovirus* genus, with three main lineages, with East African, West African, and Southern African groupings. In the absence of efficacious human drugs and/or vaccines against RVF, the virus has the potential to trigger significant public health emergencies in affected areas.

RVFV was first identified in 1931 during an investigation into an epidemic of fatalities among sheep on a farm in the Rift Valley Province of Kenya [8]. Since that time, its spatial range has continued to expand from East Africa into southern Africa, West Africa, and North Africa, and more recently outside Africa to the Arabian Peninsula [6, 7, 9]. RVFV can infect a wide range of mammalian hosts [10] and can be carried by many arthropod vectors. Floodwater *Aedes spp*. are important in maintenance during interepidemic periods, whereas other Diptera serve as secondary vectors during outbreaks in sub-tropical and temperate regions of the world [11–15]. RVFV results in lower case fatality rates (CFR) compared to other hemorrhagic fever viruses [4], but its outbreaks have profound compounding effects on human subsistence (from loss of livestock) and on national economies, related to livestock export bans [16–19].

The critical environmental reservoir of RVFV is presently unknown. Humans become infected either through the bite of an infected vector or through exposure to infectious animal tissues or bodily fluids such as abortus, birthing fluids, meat, milk, or blood aerosolized during slaughtering. Other than *in* utero transmission, there is no evidence for human-human transmission of RVFV [4]. Those infected either remain minimally symptomatic or develop a mild form of the disease Rift Valley fever (RVF) (96% of patients), which is characterized by a febrile syndrome with sudden onset of flu-like fever, muscle pain, joint pain, and headache [4]. Some patients develop neck stiffness, sensitivity to light, loss of appetite and vomiting and therefore RVF can be mistaken for meningitis [20]. While most human cases are relatively mild, a small percentage of patients will develop a much more severe form of the disease [21]. This usually appears as one (or more) of three distinct syndromes: ocular (eye) disease (0.5–2% of patients), meningoencephalitis (fewer than 1% of patients) or hemorrhagic fever (fewer than 1% of patients) [4].

Equally important is the lethal threat to peoples’ livestock. RVFV spread into naïve ecosystems is driven by infected animal movement, and potentially via infected vectors [22], and can result in death of between 70-90% of young ruminants, with sheep the most affected, and loss of pregnancy in nearly 100% of pregnant animals [23].

Outbreaks in humans and in animals do not occur at random. Instead they are strongly linked to excess rainfall and to local flooding events [24] and the consequent rise in mosquito abundance [25–32]. In addition, there is mounting evidence that suggests that there is continuing low-level RVFV transmission to humans and to animals between recognized epidemic periods [33–49]. Undetected infections, particularly in livestock, provide an important reservoir for recurrent outbreaks, leading to a continuing threat of disease in economically marginal communities and risk of further geographical expansion.

With no human-use vaccines available, the Coalition for Epidemic Preparedness Innovations (CEPI) expanded its list of target diseases to include RVF in January 2019. To date, CEPI has supported two programs to advance human RVF vaccines in collaboration with Wageningen Bioveterinary Research (WBVR), a division of Wageningen University, and Colorado State University (CSU). Both vaccine candidates are currently in the preclinical stages of development. Elsewhere, the University of Oxford has developed a single dose ChAdOx1 RVF vectored vaccine that is currently in phase 1 studies to determine the safety and immunogenicity among healthy adult volunteers (https://clinicaltrials.gov/ct2/show/NCT04672824). Previously, the US Army Medical Research Institute of Infectious Diseases (USAMRIID) developed an improved version of inactivated RVF vaccine, TSI-GSD-200, which, from early 1986 to late 1997 was given to 598 at-risk workers in their facilities, which provided effective antibody immunity after a three dose regimen [50]. However, to date, none of these vaccines has been trialed in endemic human populations, and none has undergone phase III clinical trials in order to be licensed for general use. A major challenge in planning clinical trials is the lack of precise understanding of the RVFV epidemiology and the unpredictability of outbreaks. Thus, there was need for an in depth review of existing epidemiological data and identification of remaining key knowledge gaps to further vaccine development.

The specific objective of this study was to consolidate the understanding of recent RVF epidemiology over the recent 1999-2021 period. We sought to catalogue the variability of national/regional incidence and prevalence, human risk factors and populations at risk, the geospatial distribution of RVF serotypes/lineages, and the present day national and regional human and animal RVF/viral hemorrhagic fever surveillance systems. The systematic review also highlighted current knowledge gaps in RVFV epidemiology to establish the major challenges remaining for current efforts in development and testing of human vaccines.

## Methods

To meet our study goals, we performed a systematic review of the available published literature as well as governmental monitoring and media reports of RVF activity during the period 1999-2021. The study results are reported according to the 2020 Preferred Reporting Items for Systematic Reviews and Meta-Analyses (PRISMA 2020) guidelines [51]. See S1 Table for details.

### Inclusion/exclusion criteria

Studies included in the review were those that contained reports of human, animal, and/or vector RVFV infection or exposure among individuals (of any age) in countries across Africa, the Arabian Peninsula, and the Indian Ocean where RVFV transmission was detected during the period January 1^st^ 1999 to June 1^st^ 2021, including historical, observational, and prospective studies. Serosurveys from outside this target area (Spain [52, 53], Poland [54], Korea [55], and Jordan [56]) were also catalogued, although not formally included in our analysis. Reports containing primary human seroprevalence or incidence data, reported RVF outbreaks or cases, details of RVF transmission, RVFV lineage distribution, and/or risk factors were included. Reports of infections among regional livestock and wildlife during the same period were also included.

The primary objective was to obtain population-based survey data. However, georeferenced location-specific case reports were also included in spatiotemporal analysis. There was no restriction according to the language of publication.

Excluded studies were those that reported laboratory-based studies or intervention trials among experimental animals in controlled settings. Reports, reviews, or opinion articles without primary data were also excluded.

### Information sources and search strategy

To maximize detection of eligible RVFV transmission studies, we searched the online databases PubMed, Web of Science, African Journals Online, The Cumulative Index to Nursing and Allied Health Literature (CINAHL), The Scientific Electronic Library Online (SciELO), Elsevier, ResearchGate, and the Program for Monitoring Emerging Diseases (ProMED) listserv site. Animal outbreak data recorded by the World Organization for Animal Health (OIE) was recovered from their databases, the 2005-2021 WAHIS (https://wahis.oie.int/#/home), and the older Handistatus II database (https://web.oie.int/hs2). Other sources of papers and reports that were also retrieved included: i) polling colleagues involved in RVF research or control for any non-indexed ‘grey literature’, ii) using Google Scholar referrals for ‘similar papers’, iii) scanning of literature found in personal archives, and iv) obtaining non-indexed citations found among the reference lists of the papers reviewed in our study. Details of the information sources and search strategy used are available in S1 Text.

### Selection process

Review of titles and abstracts was performed by two trained reviewers who independently searched for data content meeting study requirements. The studies found potentially suitable for inclusion after title-abstract review were then obtained for full-text review from online or library sources. Where a single report contained data on multiple individual community surveys, each of these surveys was separately abstracted and given a unique ID number for inclusion in sub-group comparison analyses. Studies with insufficient details of the incidence or prevalence of RVFV infection were not included, and cases of duplicate publication or extended analysis of previously published data were removed from the list of selected references for this review. Full listings of included and excluded references are provided as Supporting Information files S2 and S3 Tables.

### Data collection process

Included papers were abstracted by two independent reviewers, and their relevant features entered into a purpose-built database created with Google Forms. The entries were then stored in a shared master spreadsheet in Google Sheets. The included papers and data registry downloads were archived in electronic text (pdf) or spreadsheet formats at the Department of Pediatrics, Stanford University School of Medicine, and at the Center for Global Health and Diseases, Case Western Reserve University.

### Data items

For the human and animal RVFV exposures or clinical cases reported, information was entered into the database based on UN geographic region (https://unstats.un.org/unsd/methodology/m49), country, and the sub-national administrative location details that were reported. Islands in the Indian Ocean, normally classified as ‘Eastern Africa’ by the UN, were grouped separately for our analyses. The beginning and ending month and year of the reported RVF activity or exposure study were recorded, as well as information about whether the event was a recurrence of transmission in a previously affected area, whether there had been flooding before the event, and whether the period involved an El Nino-Southern Oscillation (ENSO) event. When outbreaks spanned more than one calendar year, the year of onset was used to classify the event. Study design and the eligible study population were recorded for each report and whether the report involved human cases, domestic or wildlife animals, or arthropod vectors, and the number of affected individuals by species. Where available, age and gender distributions were captured for both animal and human studies, along with any potential risk factors that were studied, and which were found to be significant by that study. Factors evaluated for human exposure risk were age and age class, gender, occupation, socio-economic status, and participation in the animal care activities of feeding, herding, sheltering, or milking livestock, as well as assisting birth, butchering, or skinning, and disposal of aborted material, or consumption of raw milk. Factors evaluated for animal RVFV exposure were age and age class, sex, breed, body condition, herd size, grazing strategy, and environmental conditions including proximity to water, local vegetation, rainfall, and mosquito control measures. Our data extraction tool was designed to capture factors related to the area the animal was raised (water sources nearby, mosquito exposures, rainfall, vegetation), herd factors (grazing strategy, mosquito control measures, herd size) and individual risk factors such as body condition and breed. In some animal studies, the age of the animal (by dental examination) and sex could be objectively assessed at the time of sampling. We grouped common animal exposures and determined how many studies had assessed for the risk factor and how many individual studies found statistically significant results by bivariate or multivariate analyses. Not all studies separated the species of animals involved within each exposure, and these have been reported as “not subdivided by species.” Information was recorded on the diagnostic tests used to identify acute cases or exposures, and on each study’s criteria used to identify suspected and confirmed animal or human cases of RVF. Available information regarding local RVF surveillance methods was noted, as well as any information about the RVFV lineage involved in the reported cases. Where individual outbreaks were reported in more than one publication or governmental bulletin, case numbers and mortality for that outbreak were taken from the latest reports.

### Study risk of bias assessment

We used an abridged version of the Liverpool Quality Appraisal Tool (LQAT) [57, 58] to assess study quality and risk of bias of included studies because of the tool’s flexibility in accommodating different study designs and in creating potential bias assessments specific to our diverse set of studies. Scoring involved assessment of possible subject selection bias, response bias, follow up bias, bias in risk factor assessment, bias in outcome assessment, or possible bias in reporting the outcome. Scoring also included whether the study’s analysis involved adjustment for potential confounders. The LQAT score, combined with recorded information about study design and power, were used to classify reports as weak (score 1-5), moderate (score 6-8), or strong evidence (score 9 or 10) regarding local risk for disease or exposures. Each study was assessed independently by two reviewers with discrepancies resolved in consultation with a third reader.

### Effect measures

We enumerated the cumulative number of outbreaks reported within national and sub-national borders over time, and the observed incidence or period prevalence by location. However, because of significant heterogeneity created by reporting bias, relative risk comparisons among the multiple locations were considered unreliable. Continent-wide RVF risk assessment for Africa, based on climate, weather, population, and landscape factors has been recently well studied [59–64] and so was not repeated here. We have, however, updated prior meta-analysis assessment [65] of individual human risk factors to provide evidence regarding specific sub-groups who might serve as suitable high-risk subjects for a human vaccine trial. Because the chances of infection were based on post hoc determination of exposures, summary odds ratios derived by random effects statistical modeling were used to determine the strength of these associations in the meta-analysis.

### Synthesis methods

Outbreaks and exposures were classified into five groups: 1) acute human RVF cases, 2) acute animal cases, 3) human RVFV exposure data (based on sero-surveys), 4) animal exposure data, and 5) arthropod infection data. These were tabulated over time and space using frequency measures and geo-located, when necessary, using Google Earth Pro software (available at https://www.google.com/earth/versions/). The two decades of inclusion (1999-2010 and 2001-2021) were assessed jointly and then separately to determine the evolution of disease, changes in detection of disease through improved diagnostics, and the effects of more robust surveillance. Regional maps at the national and sub-national level were then prepared using QGIS software (https://www.qgis.org/en/site/) using base maps supplied by the Database of Global Administrative Areas (GADM) (version 3.4, (April 2018, www.gadm.org/data, licensed for non-commercial purposes; see www.gadm.org/license). Surveillance systems in animals, humans, and vectors were categorized either as early warning systems, detection in hospital-based surveys, use of sentinel herds for surveillance, or farmer reported syndromic surveillance. For meta-analysis of individual human level risk factors, results from those included studies with relevant human exposure data were entered into Comprehensive Meta-Analysis software, v.3 (CMA, Biostat, Englewood, NJ) for calculation of pooled summary estimates of exposure effects, along with their confidence intervals. Heterogeneity levels were scored using Higgins’s and Thompson’s I^2^ statistic [66]. Summary estimates of intervention effects were computed using Der Simonian and Laird random-effects modeling [67] implemented by the CMA software. Summary data were presented visually by Forest plots showing the respective odds ratio and 95% confidence interval (CI_95%_) for the pooled analysis. Assessment for potential publication bias was carried out by visual inspection of funnel plots, and statistically by calculating the Egger test. To explore heterogeneity and factors that could potentially modify the summary estimates of effect, we performed subgroup analyses stratified by study risk of bias, and by year of publication. For the sensitivity analysis, each meta-analysis was retested with the exclusion of one study at a time to assess the possibility of a disproportionate impact of any individual study on summary estimates.

## Results

### Studies and RVFV reports selected for inclusion

Initial screening of online publication databases yielded 7097 listings for RVF or RVFV during the period January 1999-June 2021 for our initial review (Fig 1). These were supplemented by 771 georeferenced outbreak case reports obtained from OIE databases World Animal Health Information System (WAHIS, https://wahis.oie.int/#/home) for events after 2004, and from OIE Handistatus II (https://web.oie.int/hs2/report.asp?lang=en) for earlier 1999-2003 events. Other sources, including private archives, governmental reports, ProMed listings, listings from previous systematic reviews [65, 68], and non-indexed citations (found in Google Scholar and in bibliographies of papers under review), provided an additional 667 reports for consideration for possible inclusion. After removal of duplicates, 1976 unique articles or reports were selected for full text review. After the full text review was performed, 285 events reported in 281 publications, plus the 771 events identified in OIE databases and 48 from grey literature, identified a total of 1104 unique human or animal RVFV transmission events for inclusion in the analysis.

**Fig 1.**
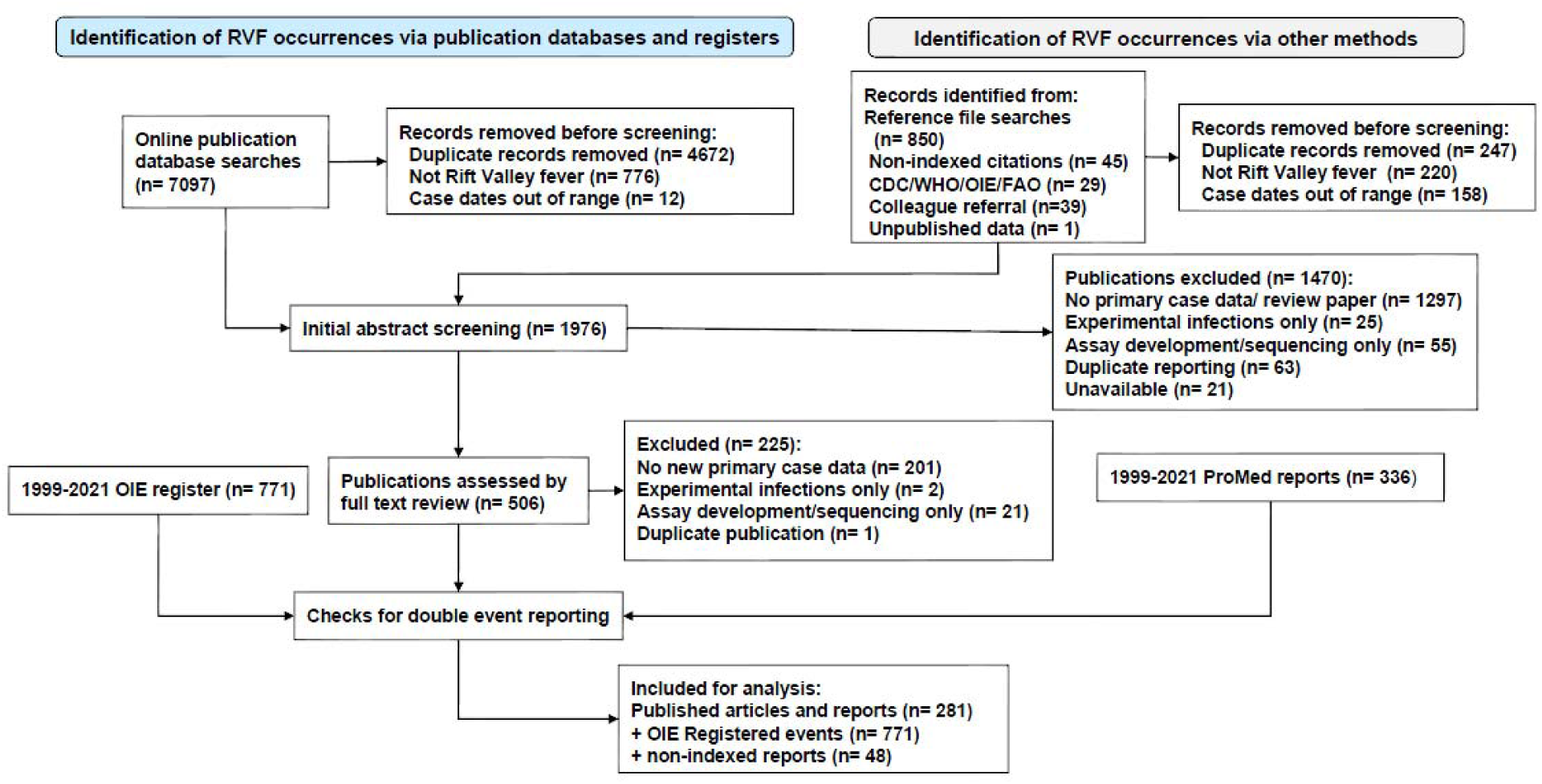
PRISMA systematic review flow diagram of data collection and evaluation. Online searches for publications and data registers (left side flow) were supplemented by governmental outbreak reports, non-indexed citations found in local archives, and citations found within the reference lists of the papers that were reviewed (right side flow).

### Report characteristics

Of the 285 included reports selected from the published literature, 91% were in peer-reviewed journals, 7% were in governmental reports, and 2% were in abstracts or published correspondence. Following the regional designations of the United Nations, 43% of these papers reported RVF activity in East Africa, 13% in West Africa, 13% in the Arabian Peninsula, 10% in North Africa, and 10% in Southern Africa. The countries of Kenya (n= 42 reports), Saudi Arabia (n= 32), Tanzania (n = 24), South Africa (n= 22), Madagascar (n= 14), Sudan (n= 13), Mauritania (n= 11), and Mozambique (n= 11) were the most frequent subjects of RVF outbreak reports and surveys. Thirty percent of reports documented active epidemics/epizootics, 61% were exposure studies performed during interepidemic periods, and 7% were post-epidemic survey studies. One hundred sixty-five (58%) were cross-sectional surveys of humans or animals, 60 (21%) were acute case series, 35 (12%) involved prospective cohorts, and 4 (1%) were case-control studies. Forty-two (15%) reported on vector testing in affected areas. Among published studies, 195 (68%) were non-randomized, 79 (28%) involved some form of random sampling, and 11 (4%) did not clearly indicate how sampling was done. In terms of study quality, the median LQAT score for published studies (where higher scores indicated higher quality studies) was 6 (range = 2-11, IQR 5-7), meaning the majority of studies had moderate-to-high risk of bias in assessing risk by location or by sub-population. Details of all included studies, including individual risk-of-bias scores are provided in S2 Table.

### Where RVFV transmission or RVF epidemics/epizootics occurred, 1999-2021

Overall, 39 countries had evidence of RVFV circulation in humans, animals, or vectors during the 1999-2021 period, based on detection of probable or confirmed acute cases, positive PCR testing, or serosurvey results (Fig 2 and S4 Table). Eighty-three reports documented 124 locations in 19 countries that had 4,353 probable or confirmed acute human RVF cases and 755 deaths (Fig 3), whereas 107 reports documented acute RVF animal events (470 OIE confirmed cases) in 31 countries between 1999 and 2021 (Fig 4). The median year of reported events was 2010. Outbreaks of clinical disease among humans or animals occurred at average rates of 5.8/year and 12.4/year, respectively, with Mauritania, Madagascar, Kenya, South Africa, and Sudan having the most human outbreak years. When month of onset was given for active epizootics or epidemics, in East Africa, 77% (27/35) of outbreaks began between November and January; for southern Africa, 80% (8/10) began between March and June; for West Africa, 92% (12/13) began between July and October; for the Arabian Peninsula, 93% (13/14) began between August and October; and for North Africa, 67% (6/9) began between September and October.

**Fig 2.**
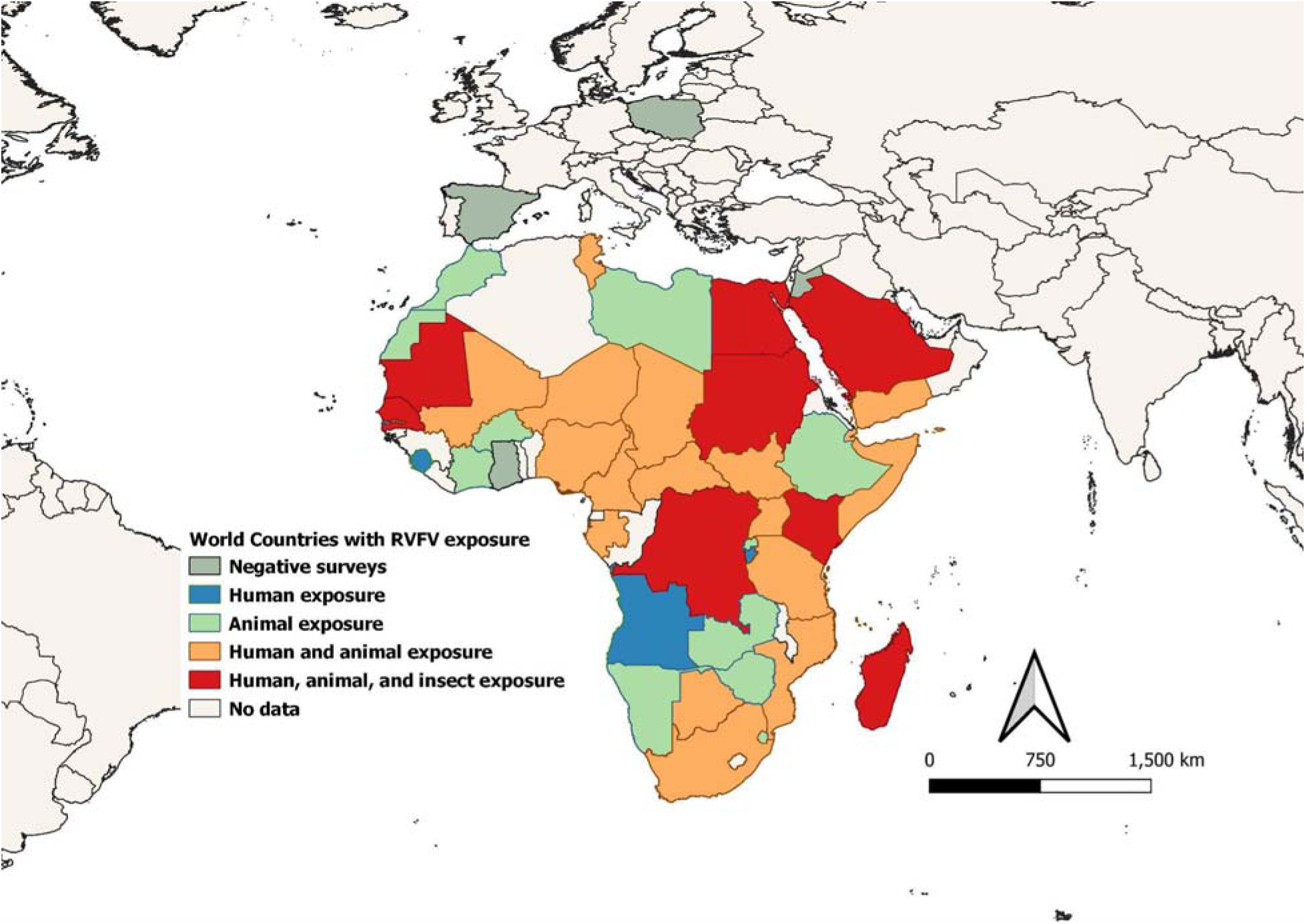
Regional map of countries exposed to RVFV infection based on findings of studies included in the systematic review. Countries were categorized as to whether there was evidence of human, animal, or arthropod RVFV infection during the 1999-2021 era. Base map is from Database of Global Administrative Areas (GADM) (version 3.4, (April 2018, www.gadm.org/data.html)

**Fig 3.**
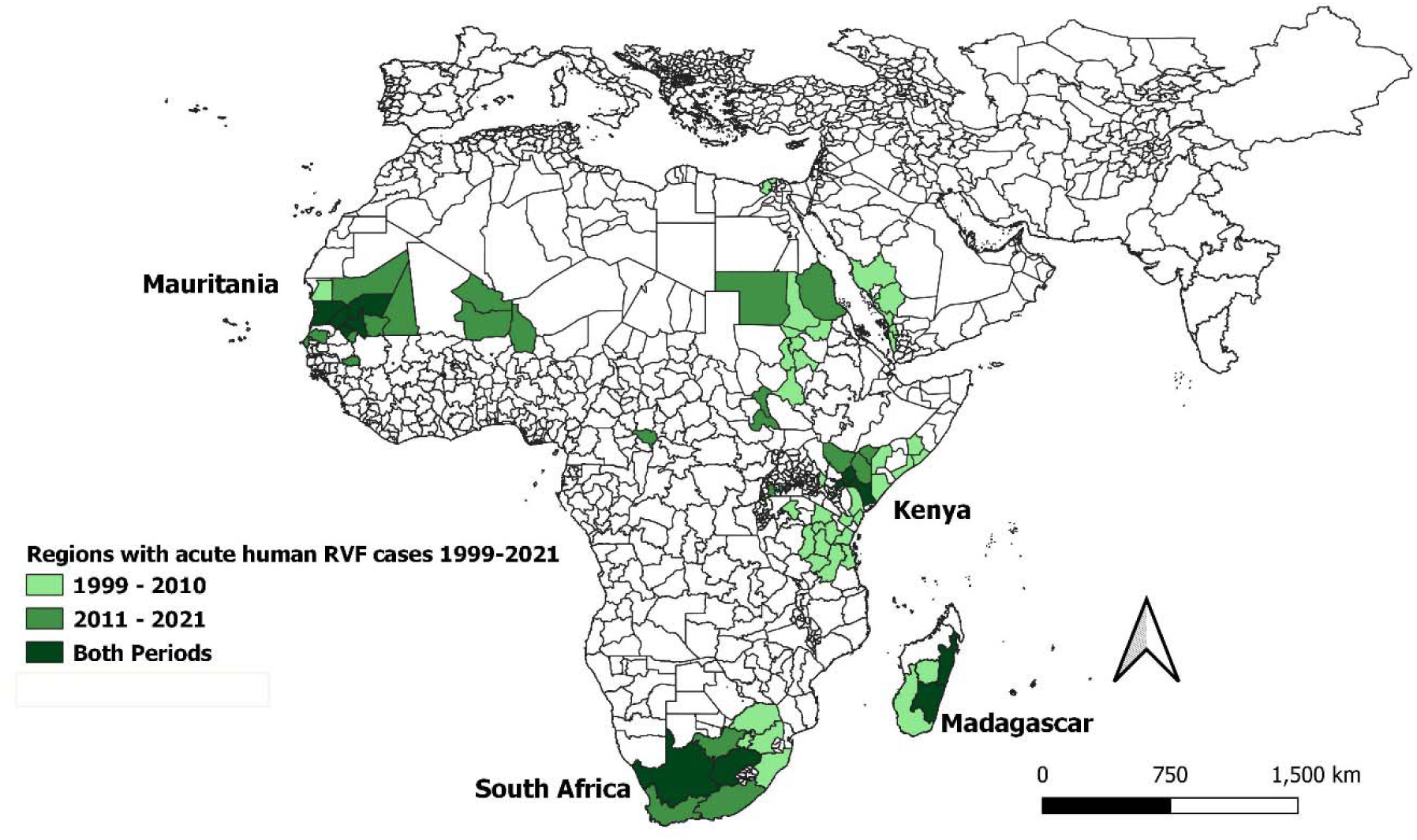
Sub-national administrative regions experiencing acute human cases of RVF during the years 1999-2021. Locations are shaded according to whether they experienced outbreaks before or after 2011, or if they had outbreaks during both periods. Base map is from Database of Global Administrative Areas (GADM) (version 3.4, (April 2018, www.gadm.org/data.html)

**Fig 4.**
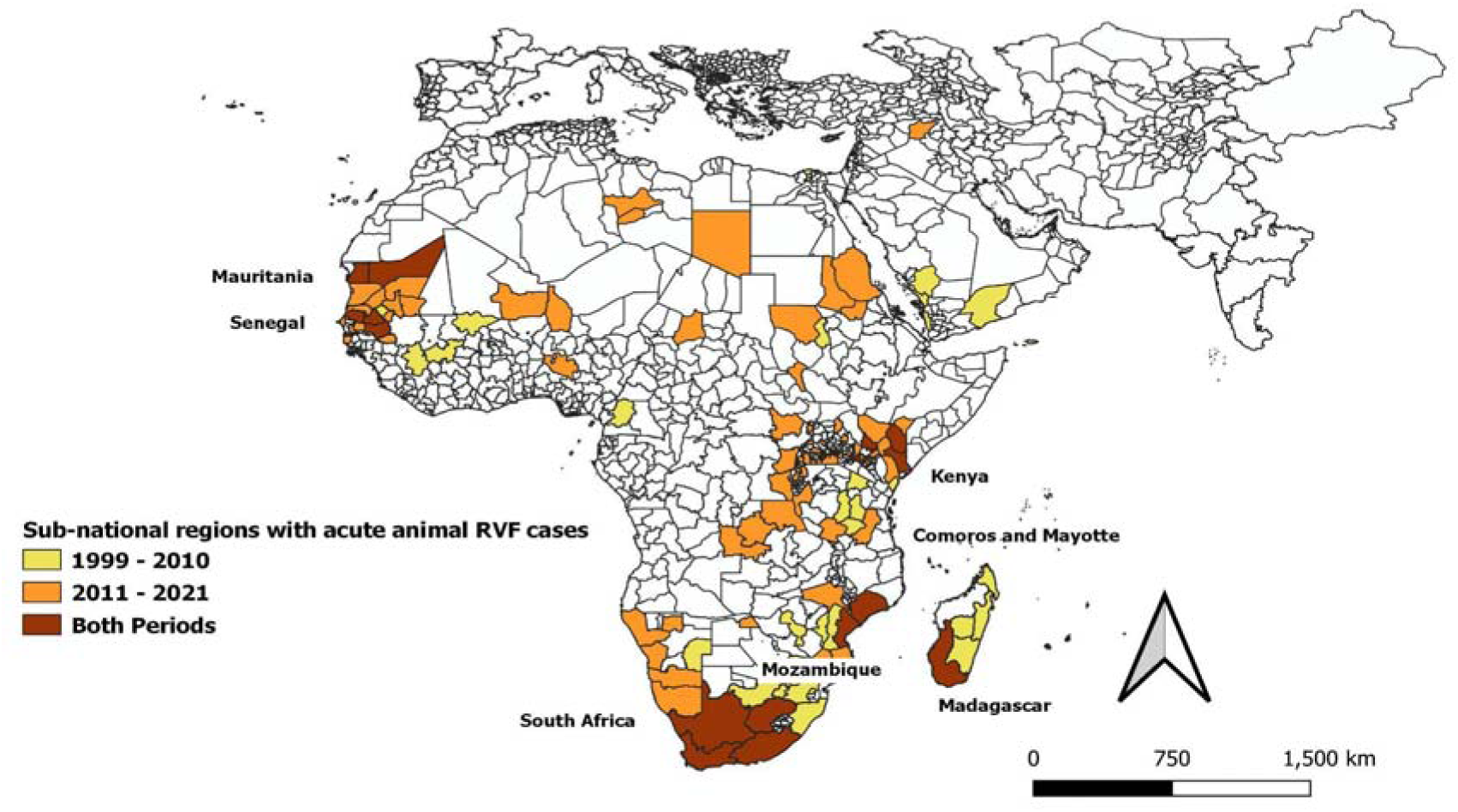
Sub-national administrative regions experiencing acute animal cases of RVF during the years 1999-2021. Locations are shaded according to whether they experienced outbreaks before or after 2011, or during both periods of time. Base map is from Database of Global Administrative Areas (GADM) (version 3.4, (April 2018, www.gadm.org/data.html)

Maps in Figs 3 and 4 show the sub-national administrative regions where acute human and animal cases were reported during the earlier (1999-2010) or latter (2011-2021) halves of our study period, or both. The countries with multiple repeated human outbreaks across both early and later time intervals were Kenya (n=4), Mauritania (n=5), South Africa (n= 4) and Madagascar (n= 2). S5 Table lists dates, place names, GPS locations, reported human case counts, incidence rates per 100,000, reported numbers of deaths, and estimated case fatality risks.

In terms of epizootic activity, Fig 4 highlights the countries that experienced repeated acute RVF outbreaks among animals during one or both decades of the 1999-2021 period. These were Kenya (n= 4), Madagascar (n= 2), Mauritania (n= 6), Mayotte (n= 2), Mozambique (n=2), Senegal (n=3), and South Africa (n=4).

### Serosurveys of human and animal RVFV exposure

Of the population surveys performed during interepidemic periods to detect evidence of prior local RVFV circulation, 50 surveyed human populations, 79 surveyed livestock populations, and 23 tested local wild animals. Of these, eight jointly surveyed both humans and livestock, and nine surveyed both wildlife and livestock. A summary of available diagnostic assays used to identify anti-RVFV antibodies is presented below in Table 1.

**Table 1.**
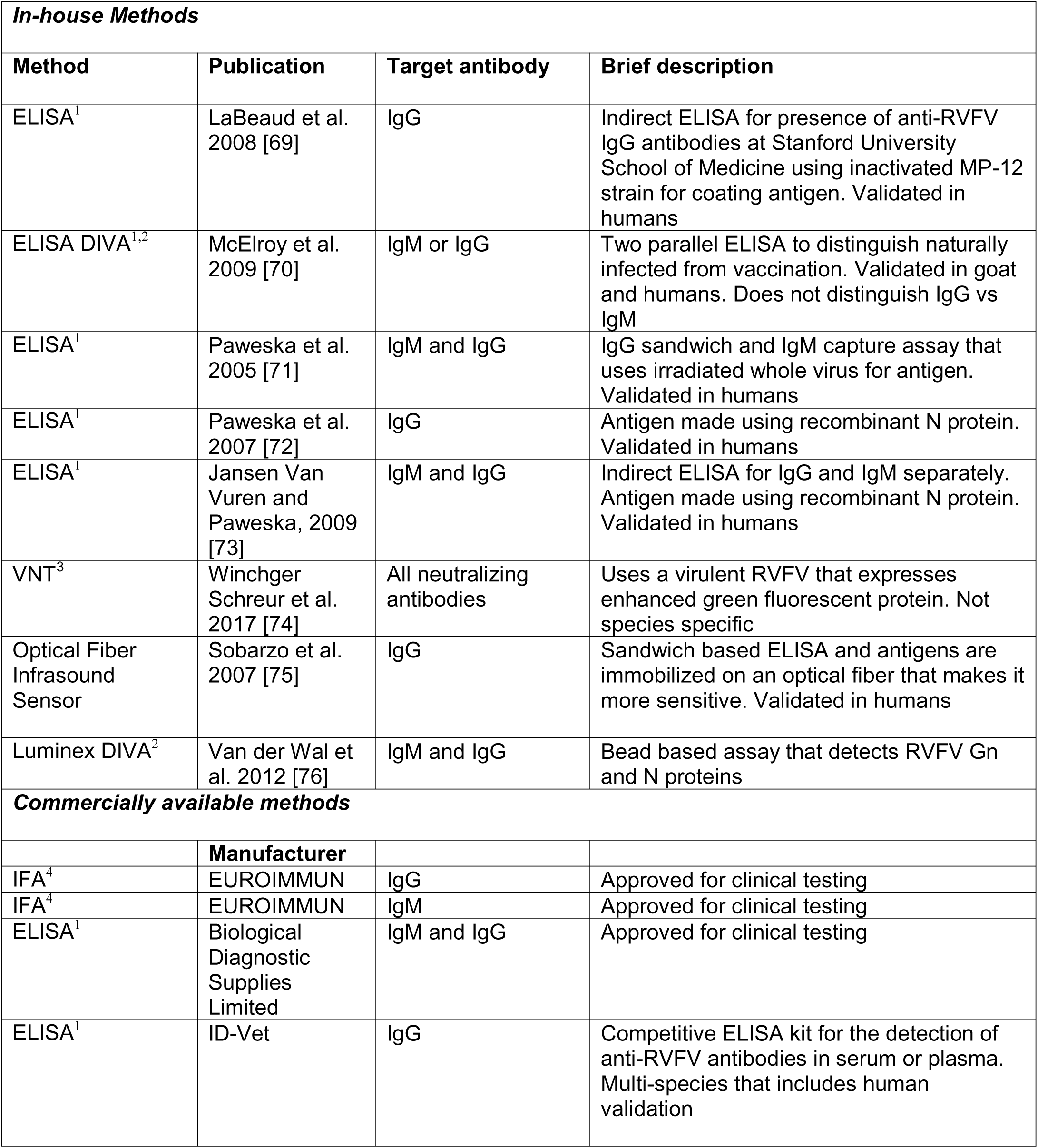
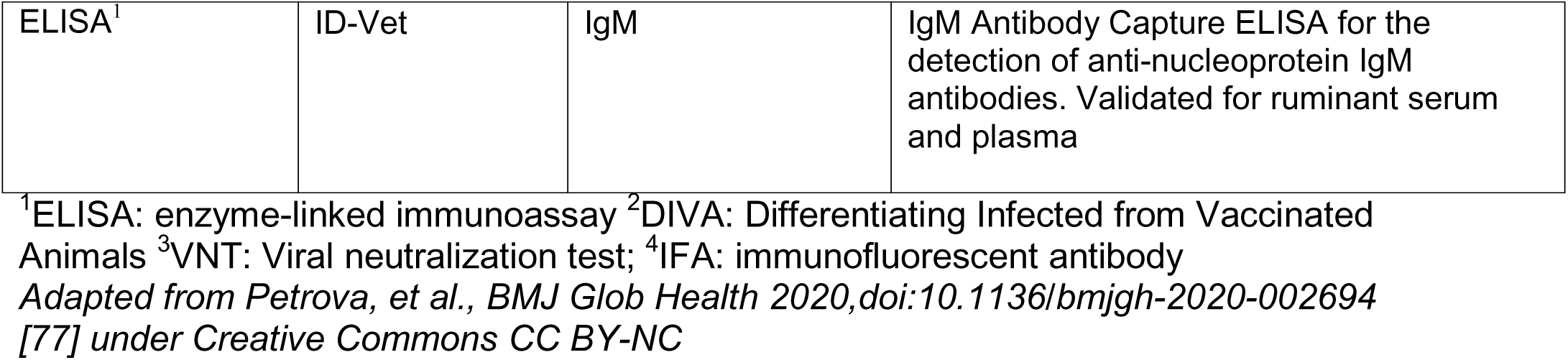
Currently used serologic assays for detection of RVFV exposure in humans.

In several animal surveys performed during interepidemic periods, low rates of livestock and wildlife anti-RVFV IgM seropositivity or IgG seroconversion were detected in areas of Namibia, Senegal, South Africa, and Kenya [47, 78–80] suggesting unsuspected ongoing RVFV transmission in those locations without detection of concurrent clinical RVF cases. Also of note were animal studies that showed serologic evidence of RVFV circulation among animals or humans within areas that had no prior evidence of RVFV circulation. These were, among animals only, in Libya [OIE], Tunisia [81], and Western Sahara [82] in North Africa; but in either humans or animals, in Sierra Leone [36], Côte d’Ivoire [83], Djibouti [84], Rwanda [85, 86], and eSwatini [OIE], i.e., geographical areas in sub-Saharan Africa with no previously documented transmission. Anti-RVFV antibody testing of human patients in health centers in Ghana [87] and Jordan [56] did not detect evidence of local exposure.

### RVFV lineage studies

Twenty-six papers reported on sequence analysis of RVFV isolates from recent and more distant outbreaks in an attempt to identify the geographic extent of specific viral lineages in circulation at the time. Although variants have been seen to cluster in East African, West African, and Southern African groupings, the results from different studies are heterogeneous, and nomenclature for these variants has not yet been standardized. The included studies on strain and lineage are summarized in S6 Table. Differences are noted depending on the segment(s) of the viral genome analyzed, the clustering algorithm used, and the reference isolate used as a ‘type specimen’ for comparison.

Sub-lineages of the Eastern group were found in the 1997-1998 Kenya/Tanzania RVF outbreak, as well as the 2000-2001 Saudi Arabia/Yemen outbreak [88, 89]. These were most similar to the RVFV isolates from humans, animals, and insects during the Kenya 2006-2007 epidemic (when variants termed Kenya-1, Kenya-2, and Tanzania-1 co-circulated) [88, 90]. Later 2008-2009 isolates from outbreaks in Comoros [91], Mayotte, and Madagascar [92, 93] were also from the Eastern group. Although Eastern African group isolates were recovered from Mauritania in 2010 [94] and Senegal in 2013 [95] in West Africa, outbreaks in these countries in other years have identified a separate West African lineage grouping of RVFV isolates [96, 97]. Grobbelaar and colleagues [98] have used the sequence from the viral Gn surface peptide to compare 198 isolates recovered between 1944-2010 from across Africa and the Arabian Peninsula, and have defined 15 different lineages, which they have labeled A through O. South African isolates in 2008 were grouped in the C lineage, related to the Kenya-1 lineage. However, during 2009-2011, South African isolates were predominantly found in a new, separate lineage H [99], while one isolate in Northern Cape Province was from lineage K. Lineage K was later identified in a small South African outbreak in 2018 [100], and in an RVFV-infected Chinese expatriate working in Angola in 2016 [101].

### Human outbreak characteristics

During human outbreaks, surveillance and reporting efforts varied greatly from location to location. We observed that among human outbreak reports, the calculated incidence rates of probable or confirmed human RVFV infections among local or regional residents varied between 0.01 and 91 per 100,000 population, with a median value of 2.5. Case fatality risks for these probable or confirmed cases varied from zero to 65% (median = 14.3%) in locations where more than one case was reported (n= 107 locations from 41 publications, see S5 Table).

The cumulative human exposure to RVFV infection is also heterogeneous across locations. In endemic areas, this cumulative exposure is usually estimated from cross-sectional surveys based on the population prevalence of human anti-RVFV IgG seropositivity at different ages. Among the 25 well-designed population-based serosurveys (scored as having low risk-of-bias) among our included reports (n= 44), IgG seroprevalence during interepidemic periods ranged from (0.15%) in Madagascar in 2014-2015 [40] to 22% in eastern Kenya in early 2006 [103] and again in 2013-2014 [104], with an overall median intra-epidemic value of 5.2% (IQR 1.8% - 13%) seropositivity across all 25 surveys.

Acute human RVF outbreaks were reported across Africa, the Arabian Peninsula, and the Indian Ocean from 2000-2021 (Fig 5). The highest number of reported human RVF events occurred in 2006-7, starting in East Africa, moving to Sudan (North Africa) and then to the islands in the Indian Ocean during 2007-2008, as part of an evolving epidemic and epizootic of the East African RVFV lineage. In East Africa, new RVFV outbreaks have recurred annually since 2016, whereas no new human cases have been reported in the Arabian Peninsula since 2001. RVF flared in southern Africa in multiple episodes between 2008 and 2011 and West African countries Mauritania and/or Senegal have reported cases across multiple years, 2003. 2010, 2012-3, 2015-6, and again in 2020.

**Fig 5.**
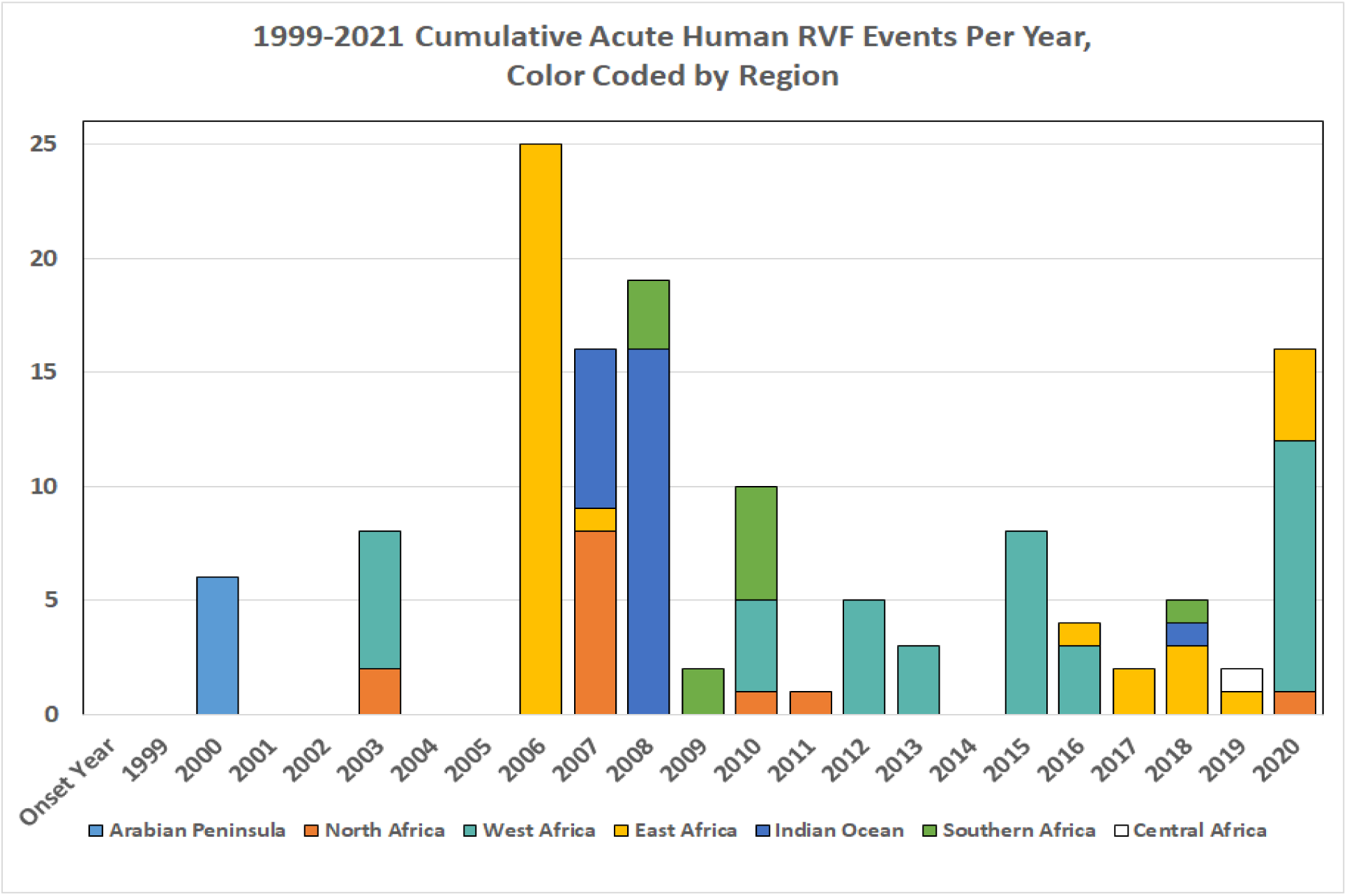
Acute human RVF events per year during 1999-2021, by geographic regions across Africa, the Indian Ocean, and the Arabian Peninsula.

A larger number of acute animal RVF events (N = 273), compared to human events (N = 132), was reported during the 1999-2021 period studied. The reported animal events in southern Africa (N= 31) and in the Indian Ocean islands (N= 19) were concentrated during the 2008-2011 period (Fig 6). By contrast, animal events in West Africa and East Africa were reported during most years of the targeted time interval (cumulative N= 80 and N=118, respectively), with the highest yearly tally being 41 reported events in 2018 (Fig 6). No animal event was reported in the Arabian Peninsula after 2010, while only a low number of events were reported in Central and North African countries between 2002-2019 (cumulative N= 8 and N= 9, respectively; Fig. 6).

**Fig 6.**
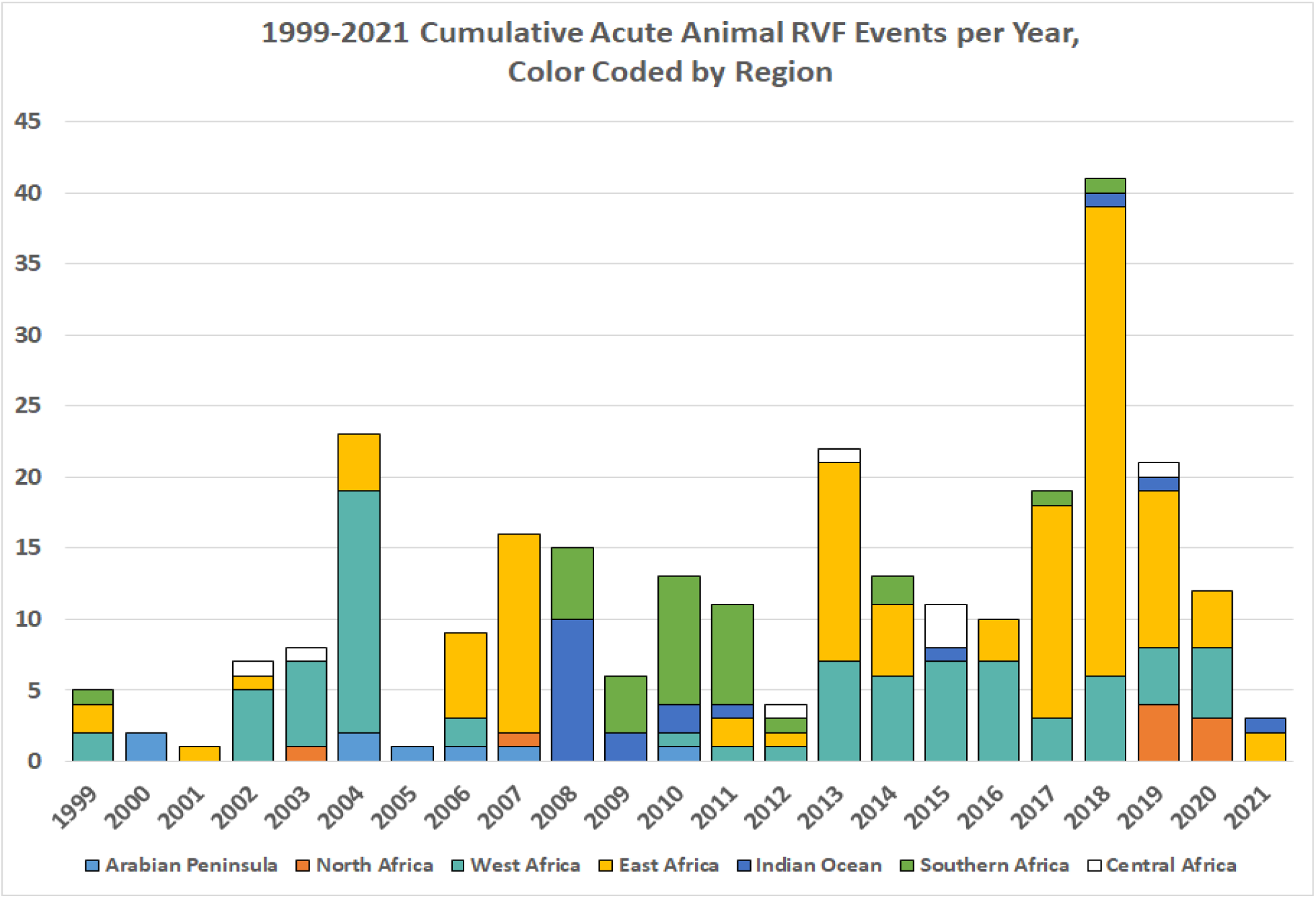
Acute animal RVF events per year during 1999-2021, by geographic regions across Africa, the Indian Ocean, and the Arabian Peninsula.

Of 28 papers that reported both human and livestock data, 17 contained information on concurrent co-local RVF epidemic and epizootic activity. These reports documented combined human/animal RVFV transmission events in Saudi Arabia [105, 106] and Yemen [107] in 2000-2001; in Egypt [108] and Mauritania [109] in 2003; in Kenya [5, 90] and Tanzania [90, 110] in 2006-2007; in Mauritania in 2010 [94, 111] and again in 2012 [112]; in Senegal in 2013-2014 [95]; in Niger [97] and in Uganda [113] in 2016; and in Kenya [114] , Mayotte [115], and South Africa [116] in 2018.

### Evidence of vector presence and competence for interepidemic RVFV transmission

In S7 Table we summarize 31 published studies from 1999-2021 that tested arthropod vector species for RVFV infection potential. Our systematic review identified 21 locations (in 8 countries) with RVFV-positive vectors, including mosquitos from *Aedes, Culex, Anopheles, and Mansonia* spp. and from *Hyalomma* spp. ticks (Fig. 2). Notably, in 6 out of 10 trapping surveys performed during interepidemic years in endemic zones, live RVFV or RVFV RNA was detected in captured vector mosquitoes or ticks [117–122]. As the lifespan of mosquito vectors is relatively short, the presence of RVFV viral RNA in these vectors likely represents sustained circulation in each of the given locations.

### Human risk of RVFV-related disease

Among our included studies from the systematic review, 84 reports assessed human risk factors for RVFV exposure. The factors most commonly assessed were occupation (N= 33 papers), gender (N= 30), age (N= 89), contact with animals (N= 38), travel (N= 15), as well as proxies for mosquito exposure such as work or residence in proximity to water sources (N= 12), and personal behaviors related to mosquito avoidance (N= 29).

Quantitative assessment of the role of gender involved estimation of chances for RVFV exposure by men and women in endemic settings. There were 31 studies that reported RVFV exposure (cases or anti RVFV seropositivity) according to gender (Fig 7). There was moderate heterogeneity among these studies (I^2^ = 39%), and random effects modeling estimation yielded a summary pooled OR estimate of 1.41 (CI_95%_ 1.24, 1.60, p < 0.001) for males vs. females in their exposure rates (Fig 7). This value, based on studies from 1999-2021, remained very similar to that calculated by Nicholas and colleagues [65] (OR = 1.36) in their meta-analysis of studies from 1984-2011.

**Fig 7.**
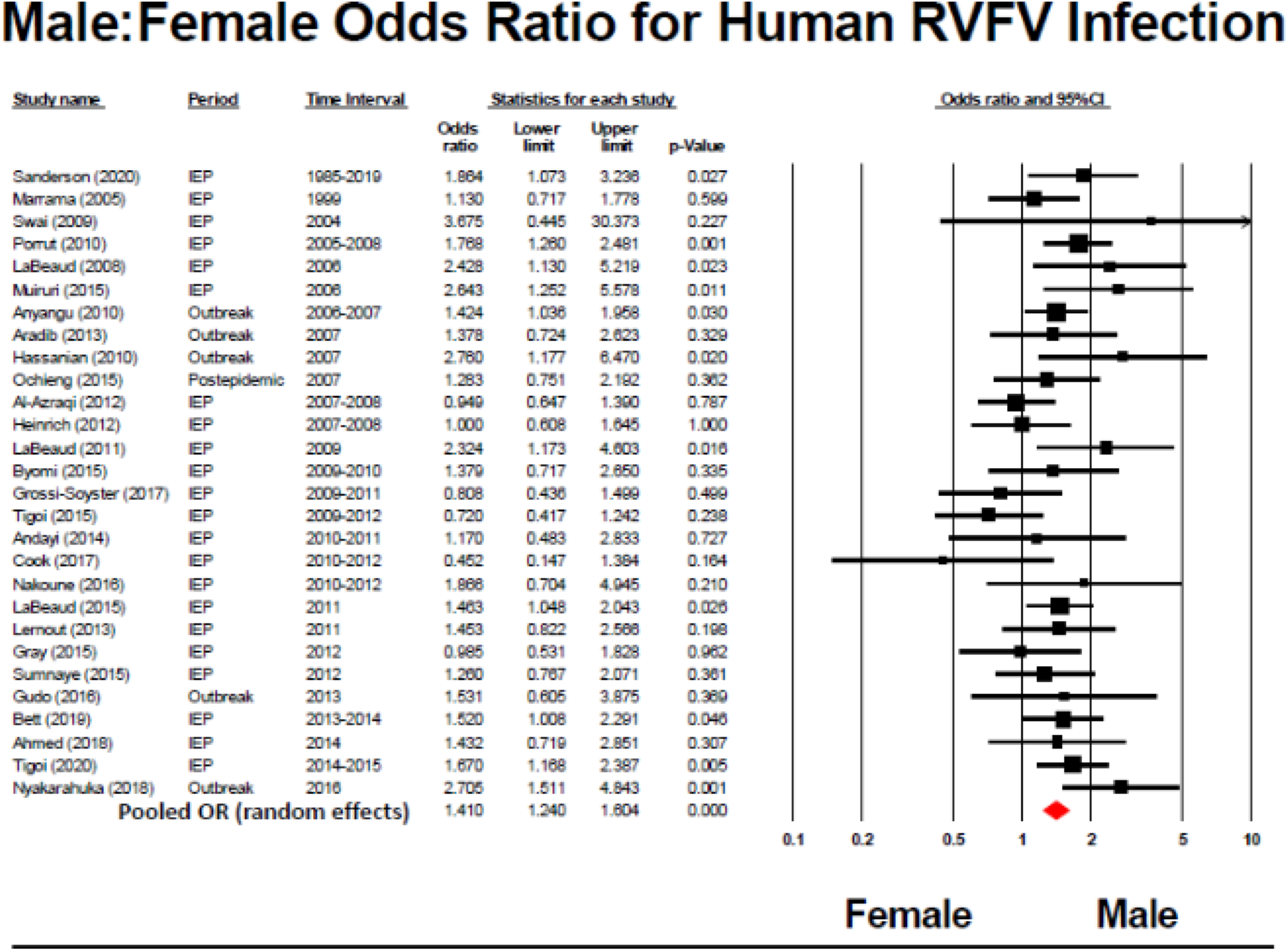
Meta-analysis of the impact of human gender on odds of RVFV exposure in at-risk populations.

### Greater human risk associated with animal contact

Eleven well-structured human surveys (having moderate to low risk of bias, i.e., LQAT scores≥ 6) compared rates of RVFV exposure among local residents according to the extent of their daily contact with animals, particularly ruminant livestock. Heterogeneity was high among these studies (I^2^ = 68%). However, random effects meta-analysis, summarized in the Forest plot in Fig 8, yielded a summary OR estimate signifying a 46% increase in the odds of RVFV exposure among people with regular animal exposure (random effects model pooled OR = 1.46, CI_95%_ = 1.09, 1.94, p= 0.01) in at risk populations. In separate task analyses, S1-S3 Figures show Forest plots for summary estimates of associations between butchering, sheltering, and milking livestock activities and risk of RVFV exposure among at-risk populations. The estimates were: for butchering, OR = 3.7 (CI_95%_ = 2.5, 5.4, p < 0.001, I^2^ = 75%); for sheltering, OR = 3.4 (CI_95%_ = 2.1, 5.4, p < 0.001, I^2^ = 57%); and for milking, OR = 5.0 (CI_95%_ = 2.8, 8.8, p < 0.001, I^2^ = 77%), indicating significantly higher risk with these exposures. Among these pooled studies, there was no evidence of publication bias (by funnel plot or by Egger’s statistic). There were no yearly trends in exposure effects, nor did sensitivity analysis indicate evidence of any single dominant study for each exposure-related outcome.

**Fig 8.**
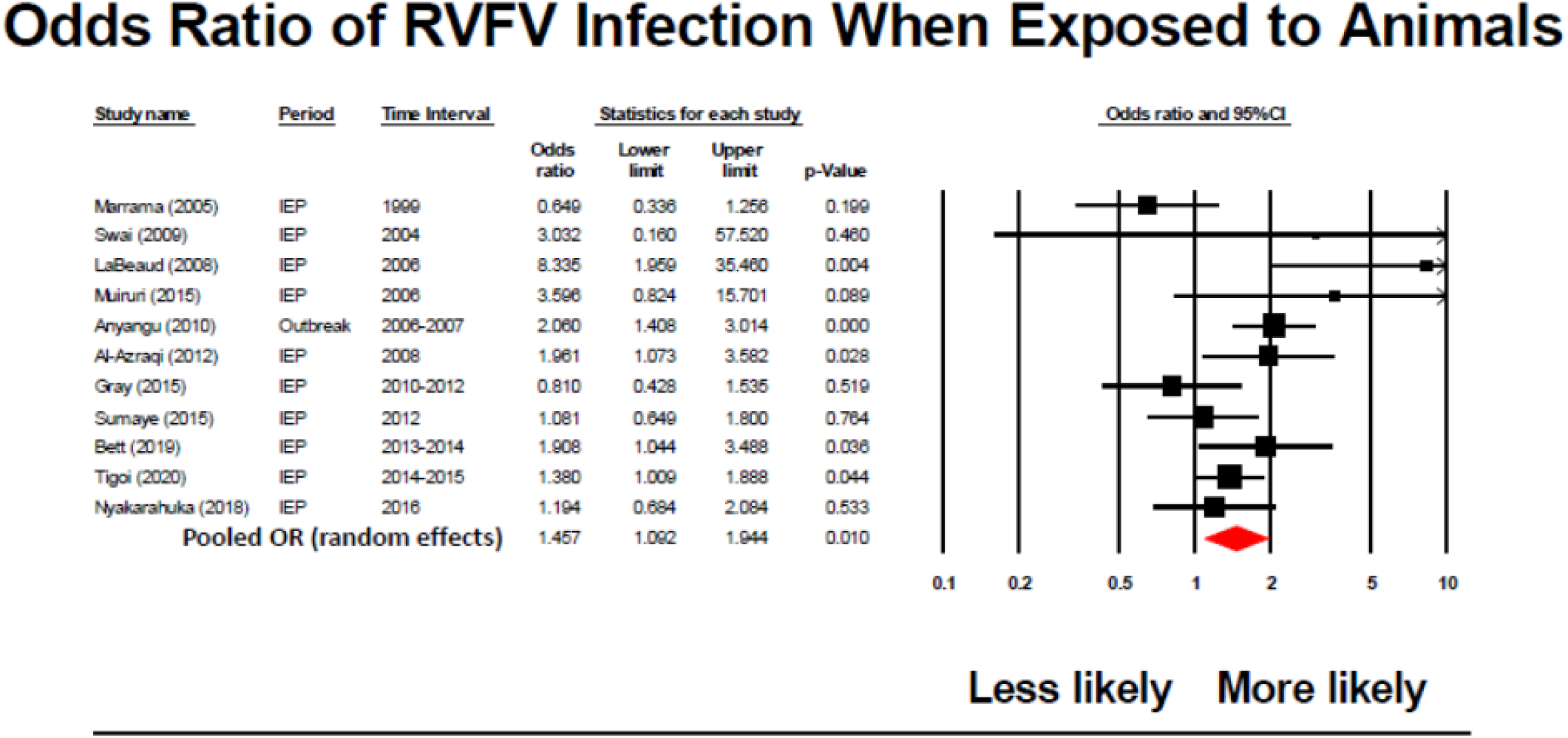
Meta-analysis of the impact of animal exposure on odds of RVFV exposure in at-risk populations.

Thirty-nine included studies assessed human RVFV-infection risk factors related to different animal exposures and 17 of those studies linked animal exposures to acute human RVFV infections (S8 Table). In meta-analyses performed according to specific animal contact tasks, handling aborted material during an RVF outbreak had a pooled odds ratio of human infection (random effects model pooled OR = 3.65, CI_95%_ = 2.32, 5.73, p < 0.001, I^2^ = 73%).

Our survey also provided individual reports of an additional 21 significant animal contact risk factors that had not been commonly assessed. In a study in Tanzania during the 2006-2007 East Africa outbreak, 40% of 115 RVFV cases reported having contact with animal products including meat and milk from sick animals, compared to just 28% of the cases reporting having slaughtered an animal [123]. Specifically, consumption of meat from sick animals was associated with a nearly four-fold increased risk of RVF-associated death during the 2006-7 outbreak in Kenya [68].

### Occupational and socioeconomic factors associated with RVFV exposure

Our systematic review identified seven studies overall that found statistically significant risks related to socioeconomic factors such as education level, ethnicity, and household size. Briefly, the less wealthy and less educated were overrepresented among RVFV exposed people (S9 Table).

Forty-five of the 506 papers assessed occupation as a human risk factor and 14 references found occupation to be a statistically significant factor (S10 Table). For clarification, those who were classified as ‘herders’ may not necessarily live a pastoralist lifestyle, so they have been given separate status. We found three studies that reported high seroprevalence among “housewives”, which might be explained by their frequent handling of raw meat and animal products for cooking.

### Risk Factors for RVF severity

Out of the 135 included papers assessing human RVFV exposure and disease, 12 presented data on human risk factors associated with more severe RVF disease presentation (S8 Table). Persons with severe disease were usually more likely to have had animal contact, [21, 124, 125]. In outbreaks in Kenya [124, 125], specific animal contact activities such as handling or consuming products from obviously sick animals (OR for more severe disease = 2.53, CI_95%_ = 1.78, 3.61, population attributable risk percentage [PAR%] = 19%) and touching an aborted animal fetus (OR = 3.83, CI_95%_ = 1.68, 9.07, PAR% = 14) were strongly linked to risk of severe (rather than mild) human RVF disease. Older age and death of a family member were also linked to more severe human disease [124]. In Sudan, males aged 15-29 years were overrepresented among patients who presented with severe disease, as compared to females of the same age [126]. Environmental factors, such as excess rainfall and muddy soil linked to emergence of mosquito blooms have also been linked to severe human disease in both Sudan and Kenya [127, 128].

Severity of human RVF was associated with the presence of concomitant co-infections. A fatal outcome in a travel related case was attributed to the presence of concurrent hepatitis A virus infection [45]. Additionally, a study in South Africa during the 2010 RVFV outbreak indicated that existing HIV-positive infection status was associated with risk of the encephalitic form of RVF disease [129], a finding similar to experience among HIV-infected patients in Tanzania in 2007, where all patients with HIV-positive status developed encephalitis, and of whom 75% died [123].

Four studies performed in three countries (Kenya, Sudan, Saudi Arabia), examined the risk factors linked to death from RVF. In Kenya in 2007, consuming and handling products from sick animals and village and district location were linked to risk of death (OR = 3.67, CI_95%_ = 1.07, 12.64, PAR% = 47%) [125, 130]. In Saudi Arabia, a retrospective analysis of the 2000-2001 outbreak showed that specific clinical signs were independently linked to death. These included jaundice, bleeding, and neurologic symptoms (P < 0.0002) [131]. One study demonstrated an increased level of RVFV replication in fatal RVFV [132].

### Risk factors for animal RVFV outbreaks

Because human risk of RVFV exposure is closely associated with the local presence of infected livestock, we reviewed factors related to animals’ risk of RVFV infection.

#### Herd Immunity Levels

Our systematic review included results from 174 animal studies, in which 144 tested domestic livestock and 26 tested wild animals for RVFV exposure and/or acute infection. Of all livestock studies included, 32 were conducted during an active epizootic, 10 were conducted just after an outbreak in the post-epidemic phase, and 97 were conducted during interepidemic years. For wildlife surveys, 5 were carried out during an active epizootic, 5 were done in the post-epidemic phase, and 14 were performed during interepidemic periods. Of all animal risk factors, sex and age were the most studied, and older animals and those that had lived through outbreaks were more likely to have been exposed. A wide range of anti-RVFV IgG seroprevalence was observed among livestock in endemic countries (4% in Senegal [95] to 39% in Tanzania [133]) and rates were dependent on species and population sampled, the timing of sampling, and the sampling strategy used. As a result, seroprevalence listed at a given time and place did not imply a specific level of long-term herd immunity. A summary of livestock risk factors is presented in S11 Table.

#### Herd Level Risks

For herd level risks, the observed effects of herd size has varied across studies. A 2013-2014 study in a high risk area of Kenya showed that medium sized herds of 50-100 animals had a significantly higher seroprevalence compared to small herds (< 50 animal) or very large herds (>100 animals) [134], though this did not account for the differences in animal rearing strategies dependent on herd size. The statistical significance of herd size was lost when village was accounted for as a random effect [74]. In Tunisia, the first assessment of serology in camels in 2017-2018 showed that camels living in smaller herds intended for meat production had a higher seroprevalence than those used for military purposes or tourism. Additionally, this study found that camels that had contact with ruminants had significantly higher rates of RVFV exposure [81].

The differing levels of RVFV susceptibility across livestock species (S11 table) further complicates the herd immunity threshold related to animal-to-human RVFV spillover. Additionally, livestock had variable contact with other livestock from neighboring herds and regions. An extensive social contact analysis from the 2008-2009 Madagascar outbreak showed that bartering practices, in which cattle can have multiple contacts within a village, was a significant seroconversion risk factor [135]. Such bartering practice can support inter-village RVFV circulation, whereas formal trade networks or cross-border smuggling were more likely to be responsible for trans-national and international spread [136, 137].

#### Individual Animal Risks

For individual-level animal risk, studies examining cattle had the most significant findings. Abortion was the most recognized clinical sign associated with RVFV infection and recent abortion was significantly associated with RVFV exposure [138]. This experience was the first to identify the 2018 RVFV outbreaks in Kenya when farmer-reported surveillance was implemented [127]. None of the included studies found an association between underlying animal body condition and RVF risk.

### Wild animals’ role in RVFV transmission

RVFV has been found in a variety of wild mammals [48]. Their role in transmission to humans has been documented, as well as their role as viral amplifiers that contribute to increased risk of livestock infection. Wild ruminants, especially buffalo, have been found to have significant herd seroprevalence during inter-epidemic period in endemic areas [46, 139, 140]. For species-specific risk factors, interspecific network centrality, home range and reproductive life-history traits were associated with RVFV occurrence. S12 Table summarizes the wild animal seropositivity and acute infections identified in our literature review.

### Animal Vaccination

Among the included papers in this review, we identified 17 studies in six countries that included vaccinated animals. Countries that had vaccinated cattle, sheep, or goats for RVFV were Kenya, Tanzania, South Africa, Egypt, and Saudi Arabia [5, 141–145]. In Egypt, between 2013 and 2015, seroprevalence of anti-RVFV was 14.9% among immunized cattle, compared to 7.9% among unimmunized cattle [146]. Although livestock are required to be vaccinated before they are exported to Saudi Arabia, a study at a livestock quarantine facility in Djibouti found an anti-RVFV IgM-positive seroprevalence of 1.2% in sheep and goats (small ruminants grouped as ‘shoats’) and 0.3% in cattle [141, 147]. A 2009 study in Mecca, Saudi Arabia found 55.8% anti-RVFV IgG in ostensibly vaccinated sheep, but still found a 2.6% rate of IgM positivity, indicating possible ongoing circulation (despite herd vaccine status) or recent vaccination [148].

### Additional considerations for RVF risks

The range of RVFV-endemic areas is expanding, and the virus appears to travel well [39, 149, 150]. In endemic countries, internal travel to or from outbreak epicenters is typically included in suspect case definitions. A recent review by Grossi-Soyster and LaBeaud [151] has highlighted the risk of RVFV transmission to travelers, and although most travelers will not have direct contact with livestock, their mosquito exposure behavior needs to be assessed.

**Reported surveillance methods** in animal, humans or vectors are summarized in S13 Table. Our review revealed 52 papers specifically reporting on RVFV surveillance. Their methods are not uniformly implemented, but include: i) leveraged RVFV testing of samples taken during surveillance for other diseases such as malaria, dengue, HIV, or Lassa fever [38, 152–154]; ii) the use of sentinel livestock herds in high-risk areas; iii) increased case surveillance based on monitoring of weather-related early warning signs; and iv) international collaborations with neighboring regions to conduct surveillance. These surveillance systems do not necessarily operate in parallel, but instead may overlap and may only be implemented at times of perceived increased risk.

Fig 10 depicts the different types of early warning and surveillance systems and their relative certainty. S13 table further summarizes available information from open-source platforms on the early warning and surveillance systems used for RVFV transmission during 1999-2021.

**Fig 9.**
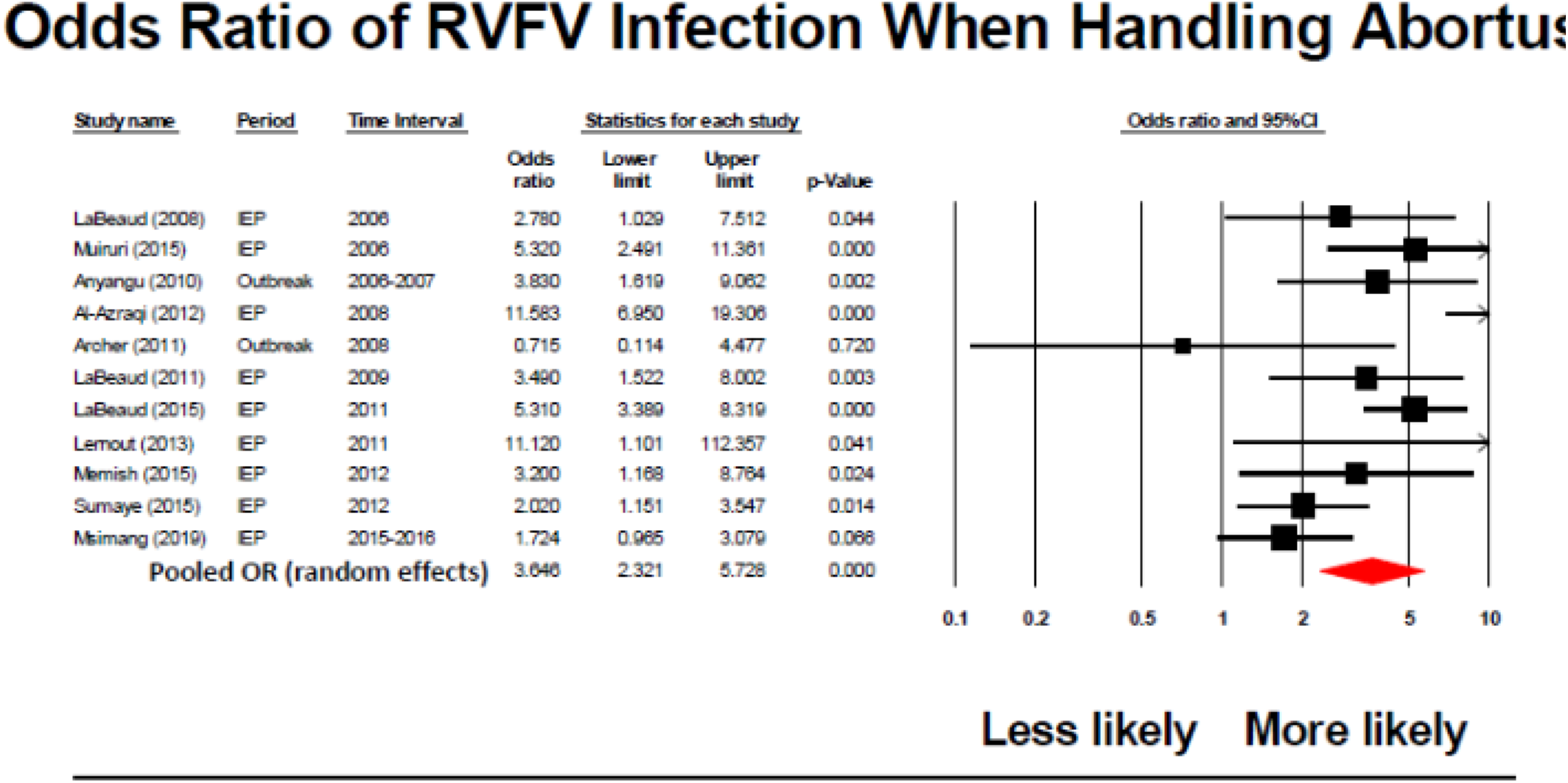
Meta-analysis of the impact of handling RVF-related animal abortions on odds of human RVFV exposure in at-risk populations.

**Fig 10.**
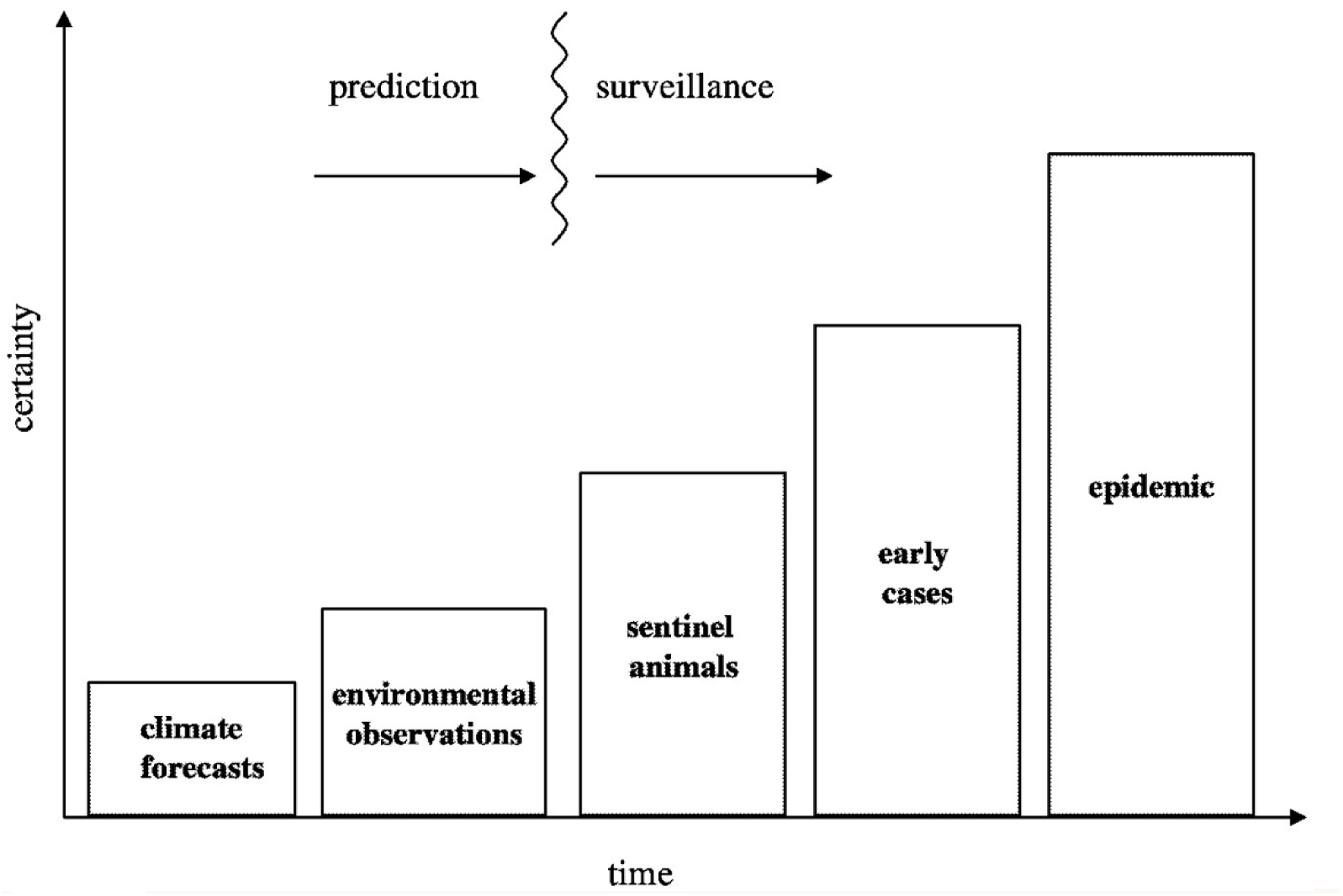
Progressive scale of surveillance that can used to indicate an impending RVFV disease outbreak. Republished with permission of The National Academies Press, from Under the Weather: Climate, Ecosystems, and Infectious Disease (2001), Chapter 7: Towards the Development of Disease Early Warning Systems. P. 87; permission conveyed through Copyright Clearance Center, Inc. under license ID #1142222

### RVFV Diagnostics

When implementing surveillance for detecting RVFV transmission, one major constraint is the limited availability of accurate diagnostics in places where RVF disease occurs. For both human and animals, reliance on a clinical presentation to prioritize RVFV testing is complicated by the significant clinical diagnostic overlap with other febrile infection syndromes.

Although detection of circulating RVFV RNA by RT-PCR can be a definitive diagnosis of active infection, because the human and livestock viremic period is typically only 4-6 days (up to 14 days if hemorrhagic human disease), serologic testing is more extensively used to investigate the epidemiology of recent and past infection. We extracted data from 207 included review reports on the type of assays recently employed for diagnosis of acute RVF. No studies tested livestock and wild animals together for acute infection. The assays used to detect acute RVFV infections are summarized in Table 3.

**Table 2.** Rift Valley fever virus lineage determinations for isolates recovered in recent epidemics, based on cluster analysis of gene sequences for the viral Gn surface protein. (Based on Grobbelaar, et al. 2011 [98])

| Grouping | Grobbelaar System | Year | Country | Species | Lineage Notes | Reference |
| --- | --- | --- | --- | --- | --- | --- |
| **East African** | lineage C | 2000-01 | Saudi Arabia, Yemen | Human | Very similar to 1997-98 isolates/variants Kenya-1, Kenya-2, and Tanzania-1 | [88, 89] |
|  | lineage C | 2006-07 | Kenya | Humans, Livestock, Vectors |  | [88, 90] |
|  | N/A | 2007 | Comoros | Humans | Similar to Kenya 2006-7 | [91] |
|  | lineage C |  | Madagascar |  |  | [92, 93] |
| **West African** |  |  | Mauritania, Senegal, Niger |  | Related to Kenya-1 lineage | [96, 97] |
| **Southern African** | lineage C | 2008-09 | South Africa |  |  | [98] |
|  | lineage H | 2009-11 | South Africa |  |  | [102] |
|  | lineage K | 2009-11 | South Africa |  |  | [102] |
|  | lineage K | 2018 | South Africa |  |  | [100] |
|  | lineage K | 2016 | Angola/China |  |  | [101] |

**Table 3:** Summary of acute diagnostic assays employed in included studies, by species.

| Number of Included Studies that Used the Following Methods for Detection of Acute RVFV Infection |  |  |  |  |  |  |  |  |  |  |  |  |
| --- | --- | --- | --- | --- | --- | --- | --- | --- | --- | --- | --- | --- |
| Species | IgM | IgM, direct<sup>1</sup> | IgM, PCR | IgM, PCR, direct<sup>1</sup> | IgM, PCR IgG SC<sup>2</sup> | IgM, IgG SC<sup>2</sup> | PCR IgG SC<sup>2</sup> | Direct<sup>1</sup> only | PCR only | PCR, direct<sup>1</sup> | IgG SC<sup>2</sup> only | Total |
| Livestock | 26 | 0 | 4 | 0 | 1 | 3 | 2 | 0 | 5 | 0 | 3 | 44 |
| Humans | 21 | 2 | 23 | 4 | 2 | 0 | 0 | 2 | 9 | 1 | 0 | 64 |
| Humans, livestock | 1 | 1 | 5 | 3 | 1 | 0 | 0 | 0 | 1 | 0 | 0 | 12 |
| Humans, livestock, wildlife | 0 | 0 | 0 | 1 | 0 | 0 | 0 | 0 | 0 | 0 | 0 | 2 |
| Wildlife | 0 | 0 | 1 | 0 | 0 | 0 | 0 | 1 | 0 | 1 | 1 | 4 |
| Method Total | 48 | 3 | 33 | 8 | 4 | 3 |  | 3 | 15 | 2 | 4 |  |

<sup>1</sup>Direct: Direct detection methods include immunofluorescence assay, cell culture (virus isolation), electron microscopy, and immunohistochemistry
<sup>2</sup>SC: IgG Seroconversion of IgG (positive IgG during the study period AND a previously documented negative status from the same individual)

## DISCUSSION

During RVF epidemics, assessment of incidence, prevalence, and associated risk factors for infection are often based on case series of suspected patients, rather than well-designed, population-based surveys that enumerate laboratory-confirmed cases vs. non-cases. Epidemic and epizootic outbreak surveys are further hampered by limited access to the affected rural communities, which are often experiencing severe flooding caused by excessive rainfall, and very limited access to diagnostic testing. In addition, survey incidence/prevalence calculations may be imprecise because of lack of accurate census information. As a result of these factors and the irregular frequency of disease outbreaks, the epidemiology of RVFV transmission and the risk factors for RVFV-related human disease are not well defined. In some endemic areas, post-epidemic serosurveys have shown high community exposure to RVFV [69, 104, 153–158], even though symptomatic RVF had been only rarely reported by local health care services. Based on our consolidated picture of recent RVFV epidemiology and the knowledge gaps we have identified, future operational research and clinical trial design should consider the factors discussed below, with the understanding that currently available data have some significant limitations in accurately defining human risk for RVF.

Our review indicated that during the last two decades, new patterns of RVF epidemiology have emerged. The virus continues to expand its range across Africa countries and into new regions within endemic countries [59, 159]. It should be noted that among studies that were technically excluded from our review, there were several completely negative serosurveys of animals performed in Spain [52], the Canary Islands [53], Mauritius [160], Zanzibar [161], South Korea [55], and Poland [54]. Expansion of RVFV transmission has been aided by livestock movement [34, 162–165] and the increasing frequency of extreme weather events involving heavy rainfall and flooding [60]. The discovery of vertical transmission within arthropod vector species other than floodwater *Aedes* spp. mosquitoes [166] indicates the existence of multiple pathways for local RVFV persistence in different ecosystems during time periods that are in between recognized outbreaks. An additional non-vector route of RVFV transmission to humans, i.e., maternal-fetal transmission, has also been identified recently [167–169], and it has been established that RVFV can be an abortifacient in humans [170]. Thus, treating RVF as a concern only during large scale outbreaks in known hotspots fails to capture the full burden of disease and its underlying transmission, and fails to detect new areas of emergence.

The review identified 9 actionable critical knowledge gaps for vaccine development and testing. Until this knowledge is improved, clinical trial planning and therefore vaccine development will continue to face practical implementation challenges that can’t easily be overcome:

1. Gap 1: Defining the likely interval between significant RVF outbreaks is challenging
2. Gap 2: The unpredictability of livestock herd immunity
3. Gap 3: Differences in seasonality across endemic/enzootic locations.
4. Gap 4: Data challenges related to bias based on limited study designs and inconsistent reporting.
5. Gap 5: Delineating human risk factors.
6. Gap 6: Need for consistent naming of RVFV lineages and strains.
7. Gap 7: Need for consistent RVFV testing frameworks.
8. Gap 8: Evolving better approaches to diagnostics.
9. Gap 9: The need for One Health approaches for RVFV detection and control.

**Gap 1: Defining the likely interval between significant RVF outbreaks is challenging** The limited understanding of transmission persistence during interepidemic periods is a major gap affecting our ability to control continued expansion of RVFV, both within and between countries. Prospective studies during interepidemic periods are needed to provide a deeper understanding of how RVFV survives then subsequently thrives in an endemic area. Study findings could then suggest how mitigation efforts, such as vaccination of high-risk humans, can be implemented even before existing surveillance systems detect an outbreak.

Defining periodicity of outbreaks is challenging because, without active case finding, passive detection is an imperfect, trailing indicator of transmission, as it requires a large enough case number of RVF clinical syndromes (e.g., abortions in livestock) to identify events. Over the last two decades, increased availability and sensitivity of RVFV diagnostic assays, including genotyping, have contributed to increased recognition of viral transmission. However, without baseline interepidemic surveys in both high and low risk areas it is not possible to determine whether RVF outbreak waves are due to persistence or to re-introduction of RVFV transmission. As the outbreak intervals for RVFV remain undefined, the lack of studies during interepidemic periods limit our ability to understand viral maintenance and make it difficult to qualify endemicity in support of mitigation efforts such as vaccination. More useful data would include concomitant active surveillance of vectors, livestock, and humans, and information on rainfall, temperature, mosquito larval sources, and the frequency of risk-related human behaviors. Because RVF livestock outbreaks are up to five times more likely to occur where outbreaks have previously occurred, and human outbreaks are more likely to occur in the same locations as livestock outbreaks [5, 171], areas with prior RVFV transmission shown in Figs. 3 and 4 would be best locations to characterize interepidemic transmission.

Kenya, South Africa, Madagascar, and Mauritania have had recurrent human and domestic animal RVF outbreaks in the last two decades and are thus most likely to experience additional outbreaks in the near future, and thus could be most suitable for a human vaccination trial. Notably, areas with recurrent outbreaks tend to have more robust surveillance which may account for the higher frequency of RVF detection in those locations (i.e., ascertainment bias). This could impact the predictability of outbreaks, and strongly affect the validity of vaccine trial endpoints. Based on findings identified in this systematic review, RVF can be more likely in a given location, compared to a similar nearby location due to many factors, including the total number of susceptible hosts (influenced by herd immunity), the presence of wildlife reservoirs [172], the water retention properties of the soil and rainfall patterns [63], local cultural practices that may increase risk (e.g., consumption of blood) or decrease risk (e.g., refusing to eat meat from dead animals) [173], and mitigation efforts that persist between outbreaks. As these factors can be confounded, an in-depth understanding of RVFV epidemiology is needed to design investigations so that resources for operational research can be maximized.

**Gap 2: The unpredictability of livestock herd immunity** Current prevention and readiness efforts for RVFV outbreaks rely mostly on early warning prediction and a rapid livestock vaccination response, both of which occur with variable reliability. Recovery from infection and vaccination with live-attenuated vaccines are thought to result in lifelong neutralizing antibodies in livestock [174] and indeed, if enough livestock could be vaccinated for RVFV, then human spillover would be unlikely [175]. However, livestock have a rapid turnover rate within and between herds as animals are slaughtered and sold, or arrive naïve into the herd from new births or purchases. Additionally, livestock in RVFV endemic countries often live in close proximity to wild ruminant species that also acquire many naïve animals each breeding season. The logistics and current intermittent nature of domestic livestock vaccination campaigns, in the face of unknown baseline herd immunity levels, makes full reliance on reactive livestock vaccination a risky approach to RVFV public health control.

Most livestock animal studies have focused on cattle, sheep, and goat seroprevalence. Although some surveys have used power analysis to determine how many animals to sample from each species, many surveys have still opted for convenience sampling, focusing either on sheep and goats, which are easier to handle, or on just cattle alone. The interpretation of such livestock prevalence surveys should be viewed with caution. Animal age is also likely to influence the results--cattle and dairy animals tend to live longer than sheep or goats, so it can appear that their cumulative burden of exposure is higher. However, evidence of infection among younger animals does provide evidence for recent RVFV transmission activity, and increased seroprevalence between years can indicate an unrecognized outbreak. When multiple species are surveyed, rates should be calculated based on animal-years at risk and better attempts should be made to age animals based on their dentition. Factors that confound RVFV exposure risk should be captured, such as the wealth and knowledge of the farmer [114], area livestock density, and location-specific climate factors. We suggest factors such as nearby temporary and permanent water bodies, land use, mobility at peak mosquito biting times, housing conditions, and rearing strategy be combined with climate data to better discern spatial and temporal high-risk periods for livestock. This will be important in defining optimal locations for human vaccination trials.

As the previous systematic review by Clark, et al. [68] has highlighted, and as we found in our review, there are few studies that assess both human and animals from the same site at the same time. The high-risk periods for livestock and humans may not necessarily overlap, as livestock infections are undoubtedly driven by rainfall and mosquito blooms, but human outbreaks can also expand with increased slaughtering of sick animals and the increased presence of secondary mosquito vectors.

It is understood that livestock viral amplification is an important predecessor of human outbreaks. The greater number of acute animal events (N=273), compared to human events (N=132) identified in this review could be due to our increasing ability to detect smaller animal outbreaks or could represent our inability to reliably detect acute human cases associated with smaller animal outbreaks [176, 177]. After animal outbreaks are established, we recommend broad-based human outbreak investigations that include all community members, not just the high-risk individuals, i.e., all individuals who may have had contact with animal products from the affected herd, as well as those who do not own livestock but reside in areas within vector flight range.

**Gap 3: Differences in seasonality across endemic/enzootic locations.** As climate change modifies seasonal weather patterns, differences in outbreak incidence are becoming more prominent among at-risk locations, but these changes may make epidemics harder to predict. Eighty-nine (31%) of our included studies specifically mention flooding at the outbreak site. A defined “rainy season” and “dry season” in tropical locations can guide risk assessment, but it is not just rain that makes an outbreak more likely. Congregation of many susceptible hosts and large volume slaughtering of animals (for example, as part of religious or community festivals [148, 178, 179]), an abundance of mosquito breeding containers (for example, in plastic trash) [180], large-scale animal movement [181], and low baseline herd immunity will increase the chance of RVFV outbreaks. Recent data-driven modeling efforts by Hardcastle and colleagues [59], incorporating environmental and human and animal census data at the district level, predict that outbreaks in southern and eastern Africa are most likely to occur in December through July, whereas risk in the Sahelian zone will be higher in September through November [182]. Even within countries, seasonal rainfall can vary widely. For example, in western Kenya, August typically has above average rainfall while the remainder of the country remains dry [183]. Real-time regional level weather information must be considered to obtain adaptive implementation of mitigation efforts.

Pastoralist herds tend to have higher RVFV exposure, and this is influenced by the types of ecozones where the herds circulate. Pastoralists follow water and pasture availability as they become available in a seasonal cycle, which may involve movement through high-risk zones. As animal protein demand increases, mostly in urban centers, more livestock will be passing through areas that have previously experienced RVF cases, and in the presence of competent mosquito vectors. Recent infections are more common in nomadic herds compared to agro-pastoralist herds [184] and in herds with longer distance to travel to their night pens or to the nearest permanent water [83]. Modeling of data from Senegal suggested that nomadic herd movements are sufficient to account for endemic circulation of RVFV, although the co-existence of *Aedes* vertical transmission cannot be ruled out [185].

In addition to climate, the variety of suitable hosts present in those areas will also vary from location to location. All potential wildlife reservoir species should be considered when comparing incidences between two geographically distinct areas because wild animals’ behaviors and range is intimately tied to topography and resource availability. In Botswana, wildlife hunters’ seroprevalence was 27% compared to a 3% community level exposure [35], showing that wildlife exposure may contribute directly to human burden independent of domestic livestock presence. This is further illustrated by human case exposure in Angola without documented livestock exposure [101]. As RVFV can adapt to new ecological niches and utilize a wide variety of mammals to amplify itself, including common white-tail deer [186, 187], we caution not to make the assumption that wild animals’ role in RVFV transmission will be similar between locations. All suitable wild animals, particularly wild ungulates, should be included in calculations of regional herd immunity and their interface with humans should be considered as a likely risk factor.

**Gap 4: Data challenges related to bias based on limited study designs and inconsistent reporting.** A systematic review by Bron and colleagues [188] has summarized the patchwork of data collected on RVFV epidemiology over the last century, and has highlighted the lack of standardized reporting as the major challenge in comparing data between locations. In our current systematic review, which focused solely on studies from the last two decades, we confirm that uneven reporting remains a significant problem. Improving study designs and sampling frameworks for RVFV is challenging, in part due to the lack of funding opportunities during interepidemic periods. In outbreak reports, uncertain levels of ascertainment bias and possible diagnostic misclassification mean that case finding and resulting case counts have unclear accuracy. ‘Population at risk’ numbers used in incidence and prevalence calculations are often chosen based on outdated census data without true understanding of who is at risk within a community. With many smaller, low-budget, short-term studies (rather than large collaborative long-term efforts) the focus turns to identifying enough cases for a meaningful statistical analysis, which means that some studies will test only high-risk individuals working with animals. This likely excludes vulnerable populations who have not yet been identified as being at high-risk.

Hospital-based surveys for hemorrhagic fever surveillance are an attractive tool because they do not require household field visits and blood samples are typically more available for testing [38, 40, 152–154, 189, 190]. Such sero-surveillance can be compared from year to year to gain retrospective insight on when transmission has occurred. However, as with most hospital-based studies, these efforts do not represent the population at large, so they likely underestimate true incidence and overestimate the percentage of cases with severe disease.

**Gap 5: Delineating human risk factors.** Limitations in our knowledge of detailed human risk factors undermines ability to use targeted strategies, such as human anti-RVFV vaccination, for prevention. For vaccine development, the predefined geographic site(s) in which clinical trials will be conducted must contain individuals who: 1) are likely to be at a high infection risk during the trial; 2) are likely to exhibit the primary endpoint upon infection; and 3) represent populations in which the vaccine will likely be distributed if found efficacious. Our meta-analysis of human risk factors across multiple 1999-2021 studies confirms the increased RVFV infection risk associated with male gender, general livestock care, contact with animal abortus, butchering animals, and milking risks identified in previous reviews and meta-analysis [65, 191]. Not all of the studies in this systematic review were population-based, thus the variable reporting of associations having ‘statistical significance’ may be attributed to differences in study design, low sample size (with lack of statistical power), and/or choice of target populations.

In most places where RVFV outbreaks occur, lack of health care infrastructure impedes detailed description of human disease signs and performance of laboratory studies (including assessment of viral load and immunologic responses to infection), which might be predictive of severe disease or death. It is possible that other health comorbidities, such as concurrent HIV or hepatitis, can influence the presentation of RVFV disease, but these associations have not yet been widely studied. Further descriptions of the natural history of clinical and laboratory findings in human RVF are needed, and efforts should be made to fully assess the relative risks from direct (non-vector) animal exposures vs. mosquito vector-borne exposure. Frontline healthcare workers need to recognize when testing for RVFV infection is appropriate, based on patients’ significant risk factors, and they should communicate frequently with local veterinary colleagues to identify high-risk transmission periods.

Most risk factor analyses rely on participant recall, risking recall bias, and it has been difficult to discriminate the risk of living in the presence of susceptible livestock from risk due to direct participation in husbandry activities or from risk due to local mosquito exposure (including variably zoophilic and anthropophilic mosquito species). Aside from animal rearing activities, the association of human risk with consumption of infected animal products is concerning, as this is practiced by a majority of human populations and the true infectivity by consumption is still not well understood. For example, consumption of raw milk is a now well recognized as an RVFV exposure risk factor [65, 191], but no previous studies have isolated RVFV from milk during outbreaks. Ongoing studies in Kenya [192] are conducting risk assessments in urban areas to distinguish consumption risk factors versus risk from livestock rearing activities common to rural areas.

RVF has been perceived as a disease of poverty, but this is likely due to its greater incidence recognized in rural areas. It is not known whether RVFV has an urban transmission cycle, and so far, the risk of RVF related to socioeconomic status (aside from having a pastoralist lifestyle) is poorly defined. Choice of occupation is intimately connected to socioeconomic status and this systematic review confirmed that the highest-risk occupations are associated with livestock exposure. Even within occupational categories, differences in assigned tasks can carry differential risks [193]. We combined common terminology for the same profession, for example, slaughterhouse and abattoir workers for our analysis; however, more specific factors such as size of the slaughterhouse, species slaughtered, and available personal protective equipment (PPE) are likely strong confounding factors.

**Gap 6: Need for consistent naming of RVFV lineages and strains.** A possible explanation for variation in RVF attack rates and disease severity, noted between locations and among different hosts, could be differences in RVFV strain virulence. Laboratory studies suggest some strain-specific differences in mortality in experimental animals [194]. However, the variability observed during natural outbreaks is likely due to site-related diagnostic misclassification of RVFV infection with erratic ability to detect milder cases of disease and variable presentations due to patients’ age and health status. Our systematic review aimed to capture strain- and lineage-specific effects; however, a meaningful analysis was obscured by inconsistent naming of lineages due to differences in the genetic sequences chosen to be studied and different approaches to sequence homology analysis. International harmonization of lineage identity and nomenclature will help to determine differential effects, if any, of individual RVFV lineages and strains. Formal delineation could be applied to study lineage and strain as a potential confounders in RVF outcome measures such as disease severity in larger-scale multi-center vaccination trials. As with SARS-CoV2 variants, a naming scheme that does not rely on location may help to alleviate some of the disincentive and stigma for national governments to report acute cases and identify sequences of circulating genomes to identify sources of local introduction.

**Gap 7: Need for consistent RVFV testing frameworks.** As highlighted earlier, identification of RVFV outbreaks and determination of the endemic state of a country require consideration of the efficacy of surveillance efforts in place. Well organized and implemented surveillance systems provide the basis for understanding the epidemiology of RVFV and provide an opportunity for warning and an opportunity to fill the gaps in current knowledge.

As RVFV diagnostics become more widely available, physicians in hyperendemic regions will be able to consider co-infection with RVFV in their differential diagnoses, especially for pregnant mothers who are at risk for abortion [170]. Additionally, if hospitals are aware of current RVFV circulation in livestock, they can go on high-alert and intensify their screening of patients for RVF. However, the lack of specific medical therapeutics may dissuade clinicians from testing for RVFV and favor testing for more treatable conditions, notably malaria. As the recent experience with Ebola virus in West Africa has taught, compassionate care must be the cornerstone of outbreak response in order for the at-risk population to buy into control measures. Public health messaging that fuels despair results in massive underreporting, particularly if outbreaks become political [195].

There are concerted efforts to create a pan-viral hemorrhagic fever diagnostic assay. The Uganda Virus Research Institute (UVRI) in Entebbe has been implementing laboratory-based surveillance for viral hemorrhagic fevers (VHF) in Uganda since 2010, which includes testing for RVFV [196]. While this may help differentiate RVFV from other concerning viruses, we caution about the grouping of RVFV with other hemorrhagic fevers for surveillance purposes as RVF-related hemorrhage in humans is the rarest clinical manifestation, and the risk factors for other forms of RVFV-related disease are different.

Development of robust RVFV surveillance systems is complex, because both animals and humans display variably detectable disease outcomes when they become infected. Currently, most RVFV surveillance systems are engaged only in response to climate early warning systems and are focused on periods with high-risk for large scale outbreaks. We found that recent re-structuring of the FAO Global Animal Disease Information System (EMPRESi) has provided an open-source, easy to navigate platform for tracking reports of RVFV cases in member countries. Since 2004, the system has recorded 1,186 animal cases of RVFV, of which 1,175 were confirmed. This reporting system relies on mandatory reporting of acute cases by national authorities, the OIE, WHO, FAO field officers, and published reports such as those included in this review. However, this database excludes longitudinal seroconversions, and therefore, excludes evidence of sub-clinical transmission during interepidemic periods and so does not capture the full extent of RVFV burden. Future modeling approaches using only these data should consider this key missing piece in their assumptions. The lack of consistent and systematic open-source data capture currently impedes the design of both animal and human vaccine trials. Although RVFV is a reportable disease in many of the affected countries, a system that relies on passive reporting is likely to miss areas of new emergence when viral activity is below the threshold to detect excess numbers of patients with the more obvious clinical signs of RVF. It should be noted that the partnership of a strong national veterinary service with close ties to a fully equipped national laboratory is crucial to any successful surveillance effort. If farmer-based cellphone reporting is to be successful at identifying RVF cases, farmers need to be sensitized to the greater goals of the system and incentivized to report. Overall, the ideal surveillance system for RVFV would utilize aspects of each of the discussed surveillance types (Fig 10), focusing on sentinels and enhanced active case finding, including during interepidemic periods, and take into account regional differences in animal and human health systems. Since RVFV is a vector-transmitted disease, and a large proportion of the world has the potential for RVFV emergence, the management of RVFV surveillance efforts should be regionally collaborated and all RVFV-naive countries with large numbers of livestock should take into account past lessons about recent introductions into new geographic areas as they prepare prevention and mitigation efforts for themselves [197–202].

Post-epidemic performance analyses have found that the lead time given by early warning systems may not be sufficiently long to prepare and vaccinate enough livestock to contain an outbreak. Just as reliance on a passive surveillance system for RVFV is likely to not be enough to detect outbreaks, sole reliance on a remotely sensed early warning systems (EWS) should also be avoided. One major limitation of climatic EWS is that they do not account for the effect of existing herd immunity, obtained either through natural infection or prior livestock vaccination. Livestock seroprevalence as high as 60% have been captured in high risk areas [203] and up to 38% in areas that are not typically classified as high risk, such as Chad [204]. The lack of outbreak detection, despite seemingly high-risk weather conditions, can be explained both by the effect of herd immunity and limitations of existing surveillance systems. With this current level of performance, at-risk countries should not rely solely on early warning systems and instead use them as part of integrated decision support tools, similar to those developed for the greater Horn of Africa [205]. As RVFV outbreaks have often had an insidious onset, we recommend active case finding in high-risk areas in response to any of the previously mentioned early warning signals.

**Gap 8: Evolving better approaches to diagnostics.** Because arboviral infections such as RVF often result in self-limited febrile syndromes with non-specific signs and symptoms, isolated cases of infection are frequently misdiagnosed as malaria in sub-Saharan Africa. This highlights the critical need for affordable and accurate point-of-care diagnostics for RVFV [206–209]. In the majority of recent transmission surveys, anti-RVFV IgM serology was used alone to define recent infection, and more humans than animals were found to be positive. As commercial IgM ELISA tests are now more widely available for livestock, this gap likely represents a lack of available resources, either financial or logistical, for testing livestock for recent infection. For suspect acute cases, IgM allows a wider window of detection than discovery of circulating virus or viral RNA [210].

Among the 95 studies that reported human serology, the methods for human anti-RVFV antibody detection exhibited significant variability, with most studies relying on tertiary reference laboratories for final diagnosis. Future efforts should focus on increasing the number and capacity of local laboratories so that the burden of shipping time and costs can be alleviated. The Covid-19 pandemic response has led to expansion of molecular diagnostic capacity in many regions. In the aftermath of the pandemic, these important logistical and laboratory resources should be leveraged to include other viral diseases such as RVF.

Serology serves as the basic approach for investigating individual and community level risk factors as it can monitor changes in prevalence over time. Both enzyme-linked immunosorbent assays (ELISA) and viral neutralization tests (VNT) have advantages and disadvantages. Identifying IgG-positive livestock by ELISA has often been the first step in recognizing new areas of emergence [81, 211]. However, follow up VNT should be used to confirm positive ELISA results in new locations, especially if the seroprevalence is low (less than 1-2%) when misclassification due to false-positive tests is of concern. For future vaccination trials, if seroconversion is the primary measure of local RVF occurrence in livestock, greater emphasis on the availability of diagnostic techniques of differentiating infected from vaccinated animals (DIVA) will be needed.

**Gap 9: The need for One Health approaches for RVFV detection and control.** Expanding RVFV surveys to include human community populations at-large will identify risk associated with socioeconomic standing, while simultaneously detecting patients with incident infections but without presently known risk factors. To fully understand the ecology of local transmission such broad-based human studies need to be conducted concurrently with robust animal surveys. Recent reviews and policy papers in the human and animal health RVFV literature call for a greater emphasis on a One Health management approach, both in research and control efforts [128, 212, 213]. For vaccination strategies, application of a One Health approach would focus on the combined use of veterinary and human vaccinations. A better understanding of viral transmission to all species during interepidemic periods, with an improvement of diagnostic sampling frameworks, are within reach in the next decade. Reaching an understanding of the herd immunity threshold for outbreak initiation, the variability in case fatality risks, the contributions of vectors to both asymptomatic and symptomatic human and animal infections, and the most influential transmission pathways will take a collaborative effort among multiple disciplines and public health sectors.

### Summary

The gaps presented here are based on a collated estimates of recent RVFV transmission. The available case counts and serosurveys in this systematic review contribute to a body of evidence that can guide the way forward to clinical testing of novel preventive and curative interventions.

The identified gaps will all affect development of human vaccines for RVF, acting as potential impediments to planning and preparing clinical field trials. Trial feasibility will depend on understanding incidence with sufficient granularity and identification of suitable target populations. Site selection faces challenges in forecasting--timing, duration and location of outbreaks, and study population selection will depend on existing seroprevalence and the duration of protection after prior natural exposure and infection. Definition of clinical endpoints will depend on good characterization of clinical presentations, and availability of diagnostic tests for differentiating non-specific febrile syndromes.

RVFV continues to transmit across Africa and nearby Indian Ocean islands at a high rate, but at an irregular frequency. As previously noted, because of its typical mosquito-borne re-emergent transmission, RVFV outbreaks often follow periods of excess rainfall and local flooding, particularly in semi-arid regions. During the 1999-2021 period studied in this review, human outbreaks of RVF occurred at an average rate of 5-6 events per year across the region. The range of transmission has expanded, with human cases documented for the first time in Saudi Arabia, Yemen, Comoros, Mayotte, Burundi, Niger, and Mali, and with re-emergence in Uganda after multiple decades of no activity. Reported human RVF events were most common in Mauritania, Kenya, Sudan, Madagascar, and South Africa, and each of these countries experienced multiple epidemics since 1999. In districts or counties where human cases were reported, the median attack rate was 2.5 per 100,000 (range 0.1 to 91), and median case fatality was 14.3% of severe cases, highlighting the significant population health burden of RVF.

Experience with veterinary vaccines indicates that pre-event vaccination can mitigate the spread of RVFV infection. Now that human vaccines are under development, their clinical testing and implementation should be guided by particular focus on the high-risk subgroups (based on location and exposures) identified in this review. Effective vaccine trials will depend on achieving accurate outbreak predictions combined with efficient animal and human case surveillance systems.

The feasibility of such trials can be assessed using the present summary of recent epidemiology, while approaches to trial design will be strengthened by simultaneously addressing the gaps we have discussed here. Recommendations for vaccine trial study design will be addressed in greater detail in a subsequent paper by the CEPI RVF study group.

## Supporting information

S1_Table Prisma checklist

S2_fig Impact of sheltering animals

S2_Table Included studies

S3_fig impact of milking

S3_Table Excluded studies

S4_Table countries with RVFV by year

S5_Table Acute human cases 1999-2021

S6_Table RVFV lineage studies

S7_Table Insect studies for RVFV

S8_Table Effect of human animal exposure

S9_Table effect of Human SES

S10_Table Effect of occupation

S11_Table Livestock risk factors for RVFV

S12_Table Wildlife RVFV seroprevalence

S13_Table surveillance systems for RVF

## Data Availability

Data collected in this systematic review and meta-analysis are available in the supporting information files posted with this article. Data were obtained using search strategies fully detailed in S1 Text, and articles chosen for full review are listed in S2 and S3 Tables. Information on outbreak locations and timing, and on the risk factors associated with human and animal RVFV infection are detailed in S4, S5, and S7-S11 Tables. Additional extracted data are: S13 Table, with local and national RVF surveillance systems reported in included studies, and S6 Table, listing studies of RVFV lineage geographic distribution based on genetic sequence analysis.

## Acknowledgments

The authors would like to acknowledge the very helpful guidance provided by members of the CEPI literature review advisory group: Dr. Brian Bird, Dr. Baptiste Dungu, Roice Fulton, Dr. Jeroen Kortekaas, Dr. Paul Oloo, Dr. Janusz Paweska, Dr. Melinda Rostal, and Dr. George Warimwe.

## Supporting Information

### Text

**S1 Text.** Information sources and detailed scripts used in running the searches on the online databases

### Figures

**S1 Figure.** Forest Plot for meta-analysis of the impact of butchering on odds of RVFV exposure

**S2 Figure.** Forest Plot for meta-analysis of the impact of sheltering animals on odds of RVFV exposure

**S3 Figure.** Forest Plot for meta-analysis of the impact of milking animals on odds of RVFV exposure

### Tables

**S1 Table.** PRISMA checklist

**S2 Table.** Listing of included studies with full citations

**S3 Table.** Listing of excluded studies with full citations

**S4 Table.** Listing by year of countries experiencing human or animal exposure to RVFV during 1999-2021

**S5 Table.** Listing of acute human cases reported during 1999-2021. Outbreaks are indicated by year, country, region, and geographic location, with estimated population size, incidence, and case fatality rates

**S6 Table.** Studies of RVFV lineage distribution based on genetic sequence analysis

**S7 Table.** 1999-2021 studies of local arthropod vector presence and RVFV infection

**S8 Table.** Studies assessing human risk factors related to animal exposures and severe RVFV disease

**S9 Table.** Studies of the effects of socioeconomic status on human RVFV exposure risk

**S10 Table.** Studies of the effects of occupation on human RVFV exposure risk

**S11 Table.** Livestock factors related to animal RVFV exposure risk

**S12 Table.** Wildlife anti-RVFV seroprevalence reported in included studies

**S13 Table.** Local and national RVF surveillance systems reported in included studies

## References

1. Gubler DJ. The global emergence/resurgence of arboviral diseases as public health problems. Arch Med Res. 2002;33(4):330–342. Epub 2002/09/18. doi: 10.1016/s0188-4409(02)00378-8.

2. CDC. Rift Valley Fever. Viral Hemorrhagic Fevers: Fact Sheets. 2002:1–3.

3. Isaäcson M. Viral hemorrhagic fever hazards for travelers in Africa. Clin Infect Dis. 2001;33(10):1707–1712. Epub 2001/10/12. doi: 10.1086/322620.

4. WHO. Rift Valley Fever. WHO Fact Sheet No 207. 2018:1–5. Epub 19 Feb 2018.

5. Munyua P, Murithi RM, Wainwright S, Githinji J, Hightower A, Mutonga D, et al. Rift Valley fever outbreak in livestock in Kenya, 2006-2007. Am J Trop Med Hyg. 2010;83(2 Suppl):58–64. Epub 2010/08/13. doi: 10.4269/ajtmh.2010.09-0292.

6. CDC. Outbreak of Rift Valley fever--Yemen, August-October 2000. MMWR. 2000;49(47):1065–1066.

7. Woods CW, Karpati AM, Grein T, McCarthy N, Gaturuku P, Muchiri E, et al. An outbreak of Rift Valley fever in Northeastern Kenya, 1997-98. Emerg Infect Dis. 2002;8(2):138–144. Epub 2002/03/19. doi: 10.3201/eid0802.010023.

8. Daubney R, Hudson JR, Garnham PC. Enzootic hepatitis or Rift Valley fever: an undescribed virus disease of sheep, cattle and man from East Africa. Journal of Pathology and Bacteriology. 1931;34:545–579.

9. CDC. Update: Outbreak of Rift Valley Fever - Saudi Arabia, August-November 2000. MMWR. 2000;49(43):982–985.

10. Golnar AJ, Turell MJ, LaBeaud AD, Kading RC, Hamer GL. Predicting the mosquito species and vertebrate species involved in the theoretical transmission of Rift Valley fever virus in the United States. PLoS Negl Trop Dis. 2014;8(9):e3163. doi: 10.1371/journal.pntd.0003163.

11. Iranpour M, Turell MJ, Lindsay LR. Potential for Canadian mosquitoes to transmit Rift Valley fever virus. J Am Mosq Control Assoc. 2011;27(4):363–369. Epub 2012/02/15. doi: 10.2987/11-6169.1.

12. Turell MJ, Dohm DJ, Geden CJ, Hogsette JA, Linthicum KJ. Potential for stable flies and house flies (Diptera: Muscidae) to transmit Rift Valley fever virus. J Am Mosq Control Assoc. 2010;26(4):445–448. Epub 2011/02/05. doi: 10.2987/10-6070.1.

13. Turell MJ, Wilson WC, Bennett KE. Potential for North American mosquitoes (Diptera: Culicidae) to transmit Rift Valley fever virus. J Med Entomol. 2010;47(5):884–889. Epub 2010/10/14. doi: 10.1603/me10007.

14. Gargan TP, 2nd, Clark GG, Dohm DJ, Turell MJ, Bailey CL. Vector potential of selected North American mosquito species for Rift Valley fever virus. Am J Trop Med Hyg. 1988;38(2):440–446. doi: 10.4269/ajtmh.1988.38.440.

15. Turell MJ, Bailey CL, Beaman JR. Vector competence of a Houston, Texas strain of *Aedes albopictus* for Rift Valley fever virus. J Am Mosq Control Assoc. 1988;4(1):94–96.

16. Chengula AA, Mdegela RH, Kasanga CJ. Socio-economic impact of Rift Valley fever to pastoralists and agro pastoralists in Arusha, Manyara and Morogoro regions in Tanzania. Springerplus. 2013;2:549. Epub 2013/11/21. doi: 10.1186/2193-1801-2-549.

17. Holleman C. The socio-economic implications of the livestock ban in Somaliland. 2002.

18. Muga GO, Onyango-Ouma W, Sang R, Affognon H. Sociocultural and economic dimensions of Rift Valley fever. Am J Trop Med Hyg. 2015;92(4):730–738. Epub 2015/02/18. doi: 10.4269/ajtmh.14-0363.

19. Rich KM, Wanyoike F. An assessment of the regional and national socio-economic impacts of the 2007 Rift Valley fever outbreak in Kenya. Am J Trop Med Hyg. 2010;83(2 Suppl):52–57. doi: 10.4269/ajtmh.2010.09-0291.

20. Riou O, Phillipe B, Jouan A, Coulibaly I, Mondo M, Digoutte JP. Les formes neurologiques et neurosensorielles de la fievre de Valley du Rift en Mauritanie. Bulletin de la Societe de Phologie Exotiques. 1989;82:605–610.

21. Kahlon SS, Peters CJ, Leduc J, Muchiri EM, Muiruri S, Njenga MK, et al. Severe Rift Valley fever may present with a characteristic clinical syndrome. Am J Trop Med Hyg. 2010;82(3):371–375. doi: 10.4269/ajtmh.2010.09-0669.

22. Kim Y, Metras R, Dommergues L, Youssouffi C, Combo S, Le Godais G, et al. The role of livestock movements in the spread of Rift Valley fever virus in animals and humans in Mayotte, 2018-19. PLoS Negl Trop Dis. 2021;15(3):e0009202. doi: 10.1371/journal.pntd.0009202.

23. OIE. Rift Valley Fever (RVF): OIE; 2021. Available from: https://www.oie.int/en/animal-health-in-the-world/animal-diseases/rift-valley-fever/.

24. Davies FG, Linthicum KJ, James AD. Rainfall and epizootic Rift Valley Fever. Bull World Health Organ. 1985;63:941–943.

25. Linthicum KJ, Anyamba A, Tucker CJ, Kelley PW, Myers MF, Peters CJ. Climate and satellite indicators to forecast Rift Valley Fever epidemics in Kenya. Science. 1999;285:397–400.

26. Linthicum KJ, Bailey CL, Davies FG, Tucker CJ. Detection of Rift Valley fever viral activity in Kenya by satellite remote sensing imagery. Science. 1987;235(4796):1656–1659. Epub 1987/03/27.

27. Linthicum KJ, Bailey CL, Tucker CJ, Mitchell KD, Logan TM, Davies FG, et al. Application of polar-orbiting, meteorological satellite data to detect flooding of Rift Valley fever virus vector mosquito habitats in Kenya. Med Vet Entomol. 1990;4(4):433–438. Epub 1990/10/01.

28. Anyamba A, Chretien JP, Formenty PB, Small J, Tucker CJ, Malone JL, et al. Rift Valley fever potential, Arabian Peninsula. Emerg Infect Dis. 2006;12(3):518–520. Epub 2006/05/23. doi: 10.3201/eid1203.050973.

29. Anyamba A, Chretien JP, Small J, Tucker CJ, Formenty PB, Richardson JH, et al. Prediction of a Rift Valley fever outbreak. Proc Natl Acad Sci U S A. 2009;106(3):955–959. doi: 10.1073/pnas.0806490106.

30. Anyamba A, Chretien JP, Small J, Tucker CJ, Linthicum KJ. Developing global climate anomalies suggest potential disease risks for 2006-2007. Int J Health Geogr. 2006;5:60. Epub 2006/12/30. doi: 10.1186/1476-072x-5-60.

31. Anyamba A, Linthicum KJ, Small J, Britch SC, Pak E, de La Rocque S, et al. Prediction, assessment of the Rift Valley fever activity in East and southern Africa 2006-2008 and possible vector control strategies. Am J Trop Med Hyg. 2010;83(2 Suppl):43–51. doi: 10.4269/ajtmh.2010.09-0289.

32. Anyamba A, Linthicum KJ, Tucker CJ. Climate-disease connections: Rift Valley fever in Kenya. Cad Saude Publica. 2001;17 Suppl:133-140. Epub 2001/06/27. doi: 10.1590/s0102-311x2001000700022.

33. Ahmed A, Makame J, Robert F, Julius K, Mecky M. Sero-prevalence and spatial distribution of Rift Valley fever infection among agro-pastoral and pastoral communities during interepidemic period in the Serengeti ecosystem, northern Tanzania. BMC Infect Dis. 2018;18(1):276. Epub 2018/06/15. doi: 10.1186/s12879-018-3183-9.

34. Tigoi C, Sang R, Chepkorir E, Orindi B, Arum SO, Mulwa F, et al. High risk for human exposure to Rift Valley fever virus in communities living along livestock movement routes: A cross-sectional survey in Kenya. PLoS Negl Trop Dis. 2020;14(2):e0007979. Epub 2020/02/23. doi: 10.1371/journal.pntd.0007979.

35. Sanderson CE, Jori F, Moolla N, Paweska JT, Oumer N, Alexander KA. Silent circulation of Rift Valley fever in humans, Botswana, 2013-2014. Emerg Infect Dis. 2020;26(10):2453–2456. Epub 2020/09/19. doi: 10.3201/eid2610.191837.

36. Schoepp RJ, Rossi CA, Khan SH, Goba A, Fair JN. Undiagnosed acute viral febrile illnesses, Sierra Leone. Emerging Infectious Diseases. 2014;20(7):1176–1182. doi: 10.3201/eid2007.131265.

37. Bosworth A, Ghabbari T, Dowall S, Varghese A, Fares W, Hewson R, et al. Serologic evidence of exposure to Rift Valley fever virus detected in Tunisia. New Microbes New Infect. 2016;9:1–7. doi: 10.1016/j.nmni.2015.10.010.

38. Dellagi K, Salez N, Maquart M, Larrieu S, Yssouf A, Silai R, et al. Serological evidence of contrasted exposure to arboviral infections between islands of the Union of Comoros (Indian Ocean). Plos Neglected Tropical Diseases. 2016;10(12). doi: 10.1371/journal.pntd.0004840.

39. Durand JP, Bouloy M, Richecoeur L, Peyrefitte CN, Tolou H. Rift Valley fever virus infection among French troops in Chad. Emerg Infect Dis. 2003;9(6):751–752. Epub 2003/06/05. doi: 10.3201/eid0906.020647.

40. Guillebaud J, Bernardson B, Randriambolamanantsoa TH, Randrianasolo L, Randriamampionona JL, Marino CA, et al. Study on causes of fever in primary healthcare center uncovers pathogens of public health concern in Madagascar. PLoS Negl Trop Dis. 2018;12(7):e0006642. Epub 2018/07/17. doi: 10.1371/journal.pntd.0006642.

41. Gudo ES, Pinto G, Weyer J, le Roux C, Mandlaze A, José AF, et al. Serological evidence of rift valley fever virus among acute febrile patients in Southern Mozambique during and after the 2013 heavy rainfall and flooding: implication for the management of febrile illness. Virol J. 2016;13:96. Epub 2016/06/10. doi: 10.1186/s12985-016-0542-2.

42. Sow A, Faye O, Faye O, Diallo D, Sadio BD, Weaver SC, et al. Rift Valley fever in Kedougou, southeastern Senegal, 2012. Emerg Infect Dis. 2014;20(3):504–506. Epub 2014/02/26. doi: 10.3201/eid2003.131174.

43. Sumaye RD, Abatih EN, Thiry E, Amuri M, Berkvens D, Geubbels E. Inter-epidemic acquisition of Rift Valley fever virus in humans in Tanzania. PLoS Negl Trop Dis. 2015;9(2):e0003536. Epub 2015/02/28. doi: 10.1371/journal.pntd.0003536.

44. Liu J, Sun Y, Shi W, Tan S, Pan Y, Cui S, et al. The first imported case of Rift Valley fever in China reveals a genetic reassortment of different viral lineages. Emerg Microbes Infect. 2017;6(1):e4. Epub 2017/01/18. doi: 10.1038/emi.2016.136.

45. Oltmann A, Kämper S, Staeck O, Schmidt-Chanasit J, Günther S, Berg T, et al. Fatal outcome of hepatitis A virus (HAV) infection in a traveler with incomplete HAV vaccination and evidence of Rift Valley fever virus infection. J Clin Microbiol. 2008;46(11):3850–3852. Epub 2008/09/05. doi: 10.1128/jcm.01102-08.

46. Evans A, Gakuya F, Paweska JT, Rostal M, Akoolo L, Van Vuren PJ, et al. Prevalence of antibodies against Rift Valley fever virus in Kenyan wildlife. Epidemiology and Infection. 2008;136(9):1261–1269. doi: 10.1017/S0950268807009806.

47. LaBeaud AD, Cross PC, Getz WM, Glinka A, King CH. Rift Valley fever virus infection in African Buffalo (*Syncerus caffer*) herds in rural South Africa: Evidence of interepidemic transmission. American Journal of Tropical Medicine and Hygiene. 2011;84(4):641–646. doi: 10.4269/ajtmh.2011.10-0187.

48. Olive MM, Goodman SM, Reynes JM. The role of wild mammals in the maintenance of Rift Valley fever virus. J Wildl Dis. 2012;48(2):241–266. doi: 10.7589/0090-3558-48.2.241.

49. Rostal MK, Evans AL, Sang R, Gikundi S, Wakhule L, Munyua P, et al. Identification of potential vectors of and detection of antibodies against Rift Valley fever virus in livestock during interepizootic periods. American Journal of Veterinary Research. 2010;71(5):522–526. doi: 10.2460/ajvr.71.5.522.

50. Pittman PR, Liu CT, Cannon TL, Makuch RS, Mangiafico JA, Gibbs PH, et al. Immunogenicity of an inactivated Rift Valley fever vaccine in humans: a 12-year experience. Vaccine. 1999;18(1-2):181–189. Epub 1999/09/29. doi: 10.1016/s0264-410x(99)00218-2.

51. Page MJ, McKenzie JE, Bossuyt PM, Boutron I, Hoffmann TC, Mulrow CD, et al. The PRISMA 2020 statement: an updated guideline for reporting systematic reviews. Bmj. 2021;372:n71. doi: 10.1136/bmj.n71.

52. García-Bocanegra I, Paniagua J, Cano-Terriza D, Arenas-Montes A, Fernández-Morente M, Napp S. Absence of Rift Valley fever virus in domestic and wild ruminants from Spain. Vet Rec. 2016;179(2):48. Epub 2016/05/11. doi: 10.1136/vr.103696.

53. Mentaberre G, Gutierrez C, Rodriguez NF, Joseph S, Gonzalez-Barrio D, Cabezon O, et al. A transversal study on antibodies against selected pathogens in dromedary camels in the Canary Islands, Spain. Veterinary Microbiology. 2013;167(3-4):468–473. doi: 10.1016/j.vetmic.2013.07.029.

54. Bazanow BA, Stygar D, Romuk E, Skrzep-Poloczek B, Pacon J, Gadzala L, et al. Preliminary serological investigation of Rift Valley fever in Poland. Journal of Vector Borne Diseases. 2018;55(4):324–326. doi: 10.4103/0972-9062.256570.

55. Kim HJ, Park JY, Jeoung HY, Yeh JY, Cho YS, Choi JS, et al. Serological surveillance studies confirm the Rift Valley fever virus free status in South Korea. Trop Anim Health Prod. 2015;47(7):1427–1430. doi: 10.1007/s11250-015-0858-8.

56. Batieha A, Saliba EK, Graham R, Mohareb E, Hijazi Y, Wijeyaratne P. Seroprevalence of West Nile, Rift Valley, and sandfly arboviruses in Hashimiah, Jordan. Emerg Infect Dis. 2000;6(4):358–362. Epub 2000/07/25. doi: 10.3201/eid0604.000405.

57. Freeman MC, Garn JV, Sclar GD, Boisson S, Medlicott K, Alexander KT, et al. The impact of sanitation on infectious disease and nutritional status: A systematic review and meta-analysis. Int J Hyg Environ Health. 2017;220(6):928–949. doi: 10.1016/j.ijheh.2017.05.007.

58. Puzzolo E, Pope D, Stanistreet D, Rehfuess EA, Bruce NG. Clean fuels for resource-poor settings: A systematic review of barriers and enablers to adoption and sustained use. Environ Res. 2016;146:218–234. doi: 10.1016/j.envres.2016.01.002.

59. Hardcastle AN, Osborne JCP, Ramshaw RE, Hulland EN, Morgan JD, Miller-Petrie MK, et al. Informing Rift Valley fever preparedness by mapping seasonally varying environmental suitability. Int J Infect Dis. 2020;99:362–372. Epub 2020/08/02. doi: 10.1016/j.ijid.2020.07.043.

60. Redding DW, Tiedt S, Lo Iacono G, Bett B, Jones KE. Spatial, seasonal and climatic predictive models of Rift Valley fever disease across Africa. Philos Trans R Soc Lond B Biol Sci. 2017;372(1725). Epub 2017/06/07. doi: 10.1098/rstb.2016.0165.

61. Williams R, Malherbe J, Weepener H, Majiwa P, Swanepoel R. Anomalous high rainfall and soil saturation as combined risk indicator of Rift Valley fever outbreaks, South Africa, 2008-2011. Emerg Infect Dis. 2016;22(12):2054–2062. Epub 2016/07/13. doi: 10.3201/eid2212.151352.

62. Hightower A, Kinkade C, Nguku PM, Anyangu A, Mutonga D, Omolo J, et al. Relationship of climate, geography, and geology to the incidence of Rift Valley fever in Kenya during the 2006-2007 outbreak. American Journal of Tropical Medicine and Hygiene. 2012;86(2):373–380. doi: 10.4269/ajtmh.2012.11-0450.

63. Verster AM, Liang JE, Rostal MK, Kemp A, Brand RF, Anyamba A, et al. Selected wetland soil properties correlate to Rift Valley fever livestock mortalities reported in 2009-10 in central South Africa. PLoS One. 2020;15(5):e0232481. Epub 2020/05/19. doi: 10.1371/journal.pone.0232481.

64. Sindato C, Pfeiffer DU, Karimuribo ED, Mboera LE, Rweyemamu MM, Paweska JT. A spatial analysis of Rift Valley fever virus seropositivity in domestic ruminants in Tanzania. PLoS One. 2015;10(7):e0131873. Epub 2015/07/15. doi: 10.1371/journal.pone.0131873.

65. Nicholas DE, Jacobsen KH, Waters NM. Risk factors associated with human Rift Valley fever infection: systematic review and meta-analysis. Trop Med Int Health. 2014;19(12):1420–1429. doi: 10.1111/tmi.12385.

66. Higgins JP, Thompson SG. Quantifying heterogeneity in a meta-analysis. Stat Med. 2002;21(11):1539–1558. Epub 2002/07/12. doi: 10.1002/sim.1186.

67. Deeks JJ, Altman DG, Bradburn MJ. Statistical methods for examining heterogeneity and combining results from several studies in meta-analysis. In: Egger M, Smith GD, Altman DG, editors. Systematic Reviews in Health Care: Meta-analysis in Context. London: BMJ Books; 2001. p. 285–312.

68. Clark MHA, Warimwe GM, Di Nardo A, Lyons NA, Gubbins S. Systematic literature review of Rift Valley fever virus seroprevalence in livestock, wildlife and humans in Africa from 1968 to 2016. Plos Neglected Tropical Diseases. 2018;12(7). doi: 10.1371/journal.pntd.0006627.

69. LaBeaud AD, Muchiri EM, Ndzovu M, Mwanje MT, Muiruri S, Peters CJ, et al. Interepidemic Rift Valley fever virus seropositivity, northeastern Kenya. Emerg Infect Dis. 2008;14(8):1240–1246. doi: 10.3201/eid1408.080082.

70. McElroy AK, Albariño CG, Nichol ST. Development of a RVFV ELISA that can distinguish infected from vaccinated animals. Virol J. 2009;6:125. Epub 2009/08/15. doi: 10.1186/1743-422x-6-125.

71. Paweska JT, Burt FJ, Swanepoel R. Validation of IgG-sandwich and IgM-capture ELISA for the detection of antibody to Rift Valley fever virus in humans. J Virol Methods. 2005;124(1-2):173–181. Epub 2005/01/25. doi: 10.1016/j.jviromet.2004.11.020.

72. Paweska JT, Jansen van Vuren P, Swanepoel R. Validation of an indirect ELISA based on a recombinant nucleocapsid protein of Rift Valley fever virus for the detection of IgG antibody in humans. J Virol Methods. 2007;146(1-2):119–124. Epub 2007/07/25. doi: 10.1016/j.jviromet.2007.06.006.

73. Jansen van Vuren P, Paweska JT. Laboratory safe detection of nucleocapsid protein of Rift Valley fever virus in human and animal specimens by a sandwich ELISA. J Virol Methods. 2009;157(1):15–24. Epub 2009/01/07. doi: 10.1016/j.jviromet.2008.12.003.

74. Wichgers Schreur PJ, Paweska JT, Kant J, Kortekaas J. A novel highly sensitive, rapid and safe Rift Valley fever virus neutralization test. J Virol Methods. 2017;248:26–30. doi: 10.1016/j.jviromet.2017.06.001.

75. Sobarzo A, Paweska JT, Herrmann S, Amir T, Marks RS, Lobel L. Optical fiber immunosensor for the detection of IgG antibody to Rift Valley fever virus in humans. Journal of Virological Methods. 2007;146(1-2):327–334. doi: 10.1016/j.jviromet.2007.07.017.

76. van der Wal FJ, Achterberg RP, de Boer SM, Boshra H, Brun A, Maassen CB, et al. Bead-based suspension array for simultaneous detection of antibodies against the Rift Valley fever virus nucleocapsid and Gn glycoprotein. J Virol Methods. 2012;183(2):99–105. doi: 10.1016/j.jviromet.2012.03.008.

77. Petrova V, Kristiansen P, Norheim G, Yimer SA. Rift valley fever: diagnostic challenges and investment needs for vaccine development. BMJ Glob Health. 2020;5(8). Epub 2020/08/21. doi: 10.1136/bmjgh-2020-002694.

78. Beechler BR, Manore CA, Reininghaus B, O’Neal D, Gorsich EE, Ezenwa VO, et al. Enemies and turncoats: bovine tuberculosis exposes pathogenic potential of Rift Valley fever virus in a common host, African buffalo (*Syncerus caffer*). Proc Biol Sci. 2015;282(1805). Epub 2015/03/20. doi: 10.1098/rspb.2014.2942.

79. Capobianco Dondona A, Aschenborn O, Pinoni C, Di Gialleonardo L, Maseke A, Bortone G, et al. Rift Valley fever virus among wild ruminants, Etosha National Park, Namibia, 2011. Emerg Infect Dis. 2016;22(1):128–130. doi: 10.3201/eid2201.150725.

80. Sindato C, Swai ES, Karimuribo ED, Dautu G, Pfeiffer DU, Mboera LEG, et al. Spatial distribution of non-clinical Rift Valley fever viral activity in domestic and wild ruminants in northern Tanzania. Tanzania Veterinary Journal. 2013;28 (Special):21–38.

81. Selmi R, Mamlouk A, Ben Said M, Ben Yahia H, Abdelaali H, Ben Chehida F, et al. First serological evidence of the Rift Valley fever Phlebovirus in Tunisian camels. Acta Trop. 2020;207:105462. Epub 2020/04/24. doi: 10.1016/j.actatropica.2020.105462.

82. Di Nardo A, Rossi D, Saleh SM, Lejlifa SM, Hamdi SJ, Di Gennaro A, et al. Evidence of Rift Valley fever seroprevalence in the Sahrawi semi-nomadic pastoralist system, Western Sahara. BMC Vet Res. 2014;10:92. Epub 2014/04/25. doi: 10.1186/1746-6148-10-92.

83. Kanouté YB, Gragnon BG, Schindler C, Bonfoh B, Schelling E. Epidemiology of brucellosis, Q Fever and Rift Valley fever at the human and livestock interface in northern Côte d’Ivoire. Acta Trop. 2017;165:66–75. Epub 2016/02/24. doi: 10.1016/j.actatropica.2016.02.012.

84. Andayi F, Charrel RN, Kieffer A, Richet H, Pastorino B, Leparc-Goffart I, et al. A sero-epidemiological study of arboviral fevers in Djibouti, Horn of Africa. PLoS Negl Trop Dis. 2014;8(12):e3299. Epub 2014/12/17. doi: 10.1371/journal.pntd.0003299.

85. Umuhoza T, Berkvens D, Gafarasi I, Rukelibuga J, Mushonga B, Biryomumaisho S. Seroprevalence of Rift Valley fever in cattle along the Akagera-Nyabarongo rivers, Rwanda. J S Afr Vet Assoc. 2017;88(0):e1–e5. doi: 10.4102/jsava.v88i0.1379.

86. Dutuze MF, Ingabire A, Gafarasi I, Uwituze S, Nzayirambaho M, Christofferson RC. Identification of Bunyamwera and possible other Orthobunyavirus infections and disease in cattle during a Rift Valley fever outbreak in Rwanda in 2018. Am J Trop Med Hyg. 2020;103(1):183–189. Epub 2020/04/22. doi: 10.4269/ajtmh.19-0596.

87. Bonney JH, Osei-Kwasi M, Adiku TK, Barnor JS, Amesiya R, Kubio C, et al. Hospital-based surveillance for viral hemorrhagic fevers and hepatitides in Ghana. PLoS Negl Trop Dis. 2013;7(9):e2435. Epub 2013/09/27. doi: 10.1371/journal.pntd.0002435.

88. Bird BH, Githinji JW, Macharia JM, Kasiiti JL, Muriithi RM, Gacheru SG, et al. Multiple virus lineages sharing recent common ancestry were associated with a large Rift Valley fever outbreak among livestock in Kenya during 2006-2007. J Virol. 2008;82(22):11152–11166. doi: 10.1128/JVI.01519-08.

89. Miller BR, Godsey MS, Crabtree MB, Savage HM, Al-Mazrao Y, Al-Jeffri MH, et al. Isolation and genetic characterization of Rift Valley fever virus from *Aedes vexans arabiensis*, Kingdom of Saudi Arabia. Emerg Infect Dis. 2002;8(12):1492–1494. Epub 2002/12/25. doi: 10.3201/eid0812.020194.

90. Nderitu L, Lee JS, Omolo J, Omulo S, O’Guinn ML, Hightower A, et al. Sequential Rift Valley fever outbreaks in eastern Africa caused by multiple lineages of the virus. J Infect Dis. 2011;203(5):655–665. doi: 10.1093/infdis/jiq004.

91. Maquart M, Pascalis H, Abdouroihamane S, Roger M, Abdourahime F, Cardinale E, et al. Phylogeographic reconstructions of a Rift Valley fever virus strain reveals transboundary animal movements from eastern continental Africa to the Union of the Comoros. Transbound Emerg Dis. 2016;63(2):e281–285. Epub 2014/09/13. doi: 10.1111/tbed.12267.

92. Ratovonjato J, Olive MM, Tantely LM, Andrianaivolambo L, Tata E, Razainirina J, et al. Detection, isolation, and genetic characterization of Rift Valley fever virus from *Anopheles (Anopheles) coustani, Anopheles (Anopheles) squamosus*, and *Culex (Culex) antennatus* of the Haute Matsiatra region, Madagascar. Vector Borne Zoonotic Dis. 2011;11(6):753–759. Epub 2010/10/30. doi: 10.1089/vbz.2010.0031.

93. Andriamandimby SF, Randrianarivo-Solofoniaina AE, Jeanmaire EM, Ravololomanana L, Razafimanantsoa LT, Rakotojoelinandrasana T, et al. Rift Valley fever during rainy seasons, Madagascar, 2008 and 2009. Emerg Infect Dis. 2010;16(6):963–970. doi: 10.3201/eid1606.091266.

94. Faye O, Ba H, Ba Y, Freire CC, Faye O, Ndiaye O, et al. Reemergence of Rift Valley fever, Mauritania, 2010. Emerg Infect Dis. 2014;20(2):300–303. Epub 2014/01/23. doi: 10.3201/eid2002.130996.

95. Sow A, Faye O, Ba Y, Diallo D, Fall G, Faye O, et al. Widespread Rift Valley fever emergence in Senegal in 2013-2014. Open Forum Infect Dis. 2016;3(3):ofw149. Epub 2016/10/06. doi: 10.1093/ofid/ofw149.

96. Sow A, Faye O, Ba Y, Ba H, Diallo D, Faye O, et al. Rift Valley fever outbreak, southern Mauritania, 2012. Emerg Infect Dis. 2014;20(2):296–299. Epub 2014/01/23. doi: 10.3201/eid2002.131000.

97. Lagare A, Fall G, Ibrahim A, Ousmane S, Sadio B, Abdoulaye M, et al. First occurrence of Rift Valley fever outbreak in Niger, 2016. Vet Med Sci. 2019;5(1):70–78. Epub 2018/11/10. doi: 10.1002/vms3.135.

98. Grobbelaar AA, Weyer J, Leman PA, Kemp A, Paweska JT, Swanepoel R. Molecular epidemiology of Rift Valley fever virus. Emerg Infect Dis. 2011;17(12):2270–2276. Epub 2011/12/17. doi: 10.3201/eid1712.111035.

99. van Schalkwyk A, Gwala S, Schuck KN, Quan M, Davis AS, Romito M, et al. Retrospective phylogenetic analyses of formalin-fixed paraffin-embedded samples from the 2011 Rift Valley fever outbreak in South Africa, through sequencing of targeted regions. J Virol Methods. 2020;287:114003. Epub 2020/11/10. doi: 10.1016/j.jviromet.2020.114003.

100. van Schalkwyk A, Romito M. Genomic characterization of Rift Valley fever virus, South Africa, 2018. Emerg Infect Dis. 2019;25(10):1979–1981. Epub 2019/09/21. doi: 10.3201/eid2510.181748.

101. Liu W, Sun FJ, Tong YG, Zhang SQ, Cao WC. Rift Valley fever virus imported into China from Angola. Lancet Infect Dis. 2016;16(11):1226. Epub 2016/10/30. doi: 10.1016/s1473-3099(16)30401-7.

102. Grobbelaar AA, Weyer J, Leman P, Kemp A, Paweska JT. Phylogeny of Rift Valley fever virus isolates recovered from humans during 2008-2011 disease outbreaks in South Africa. International Journal of Infectious Diseases. 2014;21:224–224. doi: 10.1016/j.ijid.2014.03.887.

103. Bett B, Said MY, Sang R, Bukachi S, Wanyoike S, Kifugo SC, et al. Effects of flood irrigation on the risk of selected zoonotic pathogens in an arid and semi-arid area in the eastern Kenya. PLoS One. 2017;12(5). doi: 10.1371/journal.pone.0172626.

104. LaBeaud AD, Muiruri S, Sutherland LJ, Dahir S, Gildengorin G, Morrill J, et al. Postepidemic analysis of Rift Valley fever virus transmission in northeastern Kenya: a village cohort study. PLoS Negl Trop Dis. 2011;5(8):e1265. doi: 10.1371/journal.pntd.0001265.

105. Arishi H, Ageel A, Rahman MA, Al Hazmi A, Arishi AR, Ayoola B, et al. Update: Outbreak of Rift Valley fever - Saudi Arabia, August-November 2000 (Reprinted from MMWR, vol 49, pg 982-986, 2000). Journal of the American Medical Association. 2000;284(23):2989–2990.

106. Dobson MC, Kellndorfer JM, Williams RE, Kwarteng A, editors. SAR-based land-cover classification of Kuwait. International Geoscience and Remote Sensing Symposium; 2000; Honolulu.

107. Outbreak of Rift Valley fever, Yemen, August-October 2000. Wkly Epidemiol Rec. 2000;75(48):392–395. Epub 2001/01/06.

108. Hanafi HA, Fryauff DJ, Saad MD, Soliman AK, Mohareb EW, Medhat I, et al. Virus isolations and high population density implicate *Culex antennatus* (Becker) (Diptera: Culicidae) as a vector of Rift Valley fever virus during an outbreak in the Nile Delta of Egypt. Acta Trop. 2011;119(2-3):119–124. Epub 2011/05/17. doi: 10.1016/j.actatropica.2011.04.018.

109. Faye O, Diallo M, Diop D, Bezeid OE, Bâ H, Niang M, et al. Rift Valley fever outbreak with East-Central African virus lineage in Mauritania, 2003. Emerg Infect Dis. 2007;13(7):1016–1023. Epub 2008/01/25. doi: 10.3201/eid1307.061487.

110. Sindato C, Karimuribo ED, Pfeiffer DU, Mboera LE, Kivaria F, Dautu G, et al. Spatial and temporal pattern of Rift Valley fever outbreaks in Tanzania; 1930 to 2007. PLoS One. 2014;9(2):e88897. Epub 2014/03/04. doi: 10.1371/journal.pone.0088897.

111. El Mamy AB, Baba MO, Barry Y, Isselmou K, Dia ML, El Kory MO, et al. Unexpected Rift Valley fever outbreak, northern Mauritania. Emerg Infect Dis. 2011;17(10):1894–1896. Epub 2011/10/18. doi: 10.3201/eid1710.110397.

112. El Mamy AB, Kane Y, El Arbi AS, Barry Y, Bernard C, Lancelot R, et al. L’épidémie de la fièvre de la Vallé du Rift en 2012 en Mauritanie. Revue Africaine de Santé et Productions Animales. 2014;12(3-4):169–173.

113. Shoemaker T, Balinandi S, Nyakarahuka L, Ojwang J, Tumusiime A, Mulei S, et al. First laboratory confirmation of an outbreak of Rift Valley fever virus in 50 Years in Kabale District, southwestern Uganda. American Journal of Tropical Medicine and Hygiene. 2017;95(5):437–437.

114. Hassan A, Muturi M, Mwatondo A, Omolo J, Bett B, Gikundi S, et al. Epidemiological investigation of a Rift Valley fever outbreak in humans and livestock in Kenya, 2018. Am J Trop Med Hyg. 2020;103(4):1649–1655. Epub 2020/08/05. doi: 10.4269/ajtmh.20-0387.

115. Youssouf H, Subiros M, Dennetiere G, Collet L, Dommergues L, Pauvert A, et al. Rift Valley fever outbreak, Mayotte, France, 2018-2019. Emerg Infect Dis. 2020;26(4):769–772. Epub 2020/03/19. doi: 10.3201/eid2604.191147.

116. van Vuren PJ, Kgaladi J, Patharoo V, Ohaebosim P, Msimang V, Nyokong B, et al. Human cases of Rift Valley fever in South Africa, 2018. Vector-Borne and Zoonotic Diseases. 2018;18(12):713–715. doi: 10.1089/vbz.2018.2357.

117. Youssef HM, Ghoneim MA, al. e. Recent trends for diagnosis of Rift Valley fever in animals and mosquitoes in Egypt with special reference to the carrier. Global Veterinaria. 2008;2(1).

118. Jeffries CL, Tantely LM, Raharimalala FN, Hurn E, Boyer S, Walker T. Diverse novel resident *Wolbachia* strains in Culicine mosquitoes from Madagascar. Scientific Reports. 2018;8. doi: 10.1038/s41598-018-35658-z.

119. Ndiaye EH, Diallo D, Fall G, Ba Y, Faye O, Dia I, et al. Arboviruses isolated from the Barkedji mosquito-based surveillance system, 2012-2013. BMC Infect Dis. 2018;18(1):642. Epub 2018/12/14. doi: 10.1186/s12879-018-3538-2.

120. Abdelgadir DM, Bashab HMM, Mohamed RAE, Abuelmaali SA. Risk factor analysis for outbreak of Rift Valley fever in Khartoum State of Sudan. Journal of Entomological Science. 2010;45(3):239–251. doi: 10.18474/0749-8004-45.3.239.

121. Mbanzulu KM, Mboera LEG, Luzolo FK, Wumba R, Misinzo G, Kimera SI. Mosquito-borne viral diseases in the Democratic Republic of the Congo: a review. Parasit Vectors. 2020;13(1):103. Epub 2020/02/28. doi: 10.1186/s13071-020-3985-7.

122. Mohamed RAE, Mohamed N, Aleanizy FS, Alqahtani FY, Al Khalaf A, Al-Keridis LA. Investigation of hemorrhagic fever viruses inside wild populations of ticks: One of the pioneer studies in Saudi Arabia. Asian Pacific Journal of Tropical Disease. 2017;7:299–303.

123. Mohamed M, Mosha F, Mghamba J, Zaki SR, Shieh WJ, Paweska J, et al. Epidemiologic and clinical aspects of a Rift Valley fever outbreak in humans in Tanzania, 2007. Am J Trop Med Hyg. 2010;83(2 Suppl):22–27. doi: 10.4269/ajtmh.2010.09-0318.

124. LaBeaud AD, Pfeil S, Muiruri S, Dahir S, Sutherland LJ, Traylor Z, et al. Factors associated with severe human Rift Valley fever in Sangailu, Garissa County, Kenya. PLoS Negl Trop Dis. 2015;9(3):e0003548. Epub 2015/03/13. doi: 10.1371/journal.pntd.0003548.

125. Anyangu AS, Gould LH, Sharif SK, Nguku PM, Omolo JO, Mutonga D, et al. Risk factors for severe Rift Valley fever infection in Kenya, 2007. Am J Trop Med Hyg. 2010;83(2 Suppl):14–21. doi: 10.4269/ajtmh.2010.09-0293.

126. Seufi AM, Galal FH. Role of *Culex* and *Anopheles* mosquito species as potential vectors of Rift Valley fever virus in Sudan outbreak, 2007. BMC Infect Dis. 2010;10:65. Epub 2010/03/13. doi: 10.1186/1471-2334-10-65.

127. Oyas H, Holmstrom L, Kemunto NP, Muturi M, Mwatondo A, Osoro E, et al. Enhanced surveillance for Rift Valley fever in livestock during El Niño rains and threat of RVF outbreak, Kenya, 2015-2016. PLoS Negl Trop Dis. 2018;12(4):e0006353. Epub 2018/04/27. doi: 10.1371/journal.pntd.0006353.

128. Bashir RSE, Hassan OA. A One Health perspective to identify environmental factors that affect Rift Valley fever transmission in Gezira state, Central Sudan. Tropical Medicine and Health. 2019;47(1). doi: 10.1186/s41182-019-0178-1.

129. van Vuren PJ, Shalekoff S, Grobbelaar AA, Archer BN, Thomas J, Tiemessen CT, et al. Serum levels of inflammatory cytokines in Rift Valley fever patients are indicative of severe disease. Virology Journal. 2015;12. doi: 10.1186/s12985-015-0392-3.

130. Outbreak news. Rift Valley fever, Kenya. Wkly Epidemiol Rec. 2007;82(3):17–18. Epub 2007/01/24.

131. Madani TA, Al-Mazrou YY, Al-Jeffri MH, Mishkhas AA, Al-Rabeah AM, Turkistani AM, et al. Rift Valley fever epidemic in Saudi Arabia: epidemiological, clinical, and laboratory characteristics. Clin Infect Dis. 2003;37(8):1084–1092. doi: 10.1086/378747.

132. Njenga MK, Paweska J, Wanjala R, Rao CY, Weiner M, Omballa V, et al. Using a field quantitative real-time PCR test to rapidly identify highly viremic rift valley fever cases. J Clin Microbiol. 2009;47(4):1166–1171. Epub 2009/01/28. doi: 10.1128/jcm.01905-08.

133. Chengula AA, Kasanga CJ, Mdegela RH, Sallu R, Yongolo M. Molecular detection of Rift Valley fever virus in serum samples from selected areas of Tanzania. Trop Anim Health Prod. 2014;46(4):629–634. Epub 2014/01/28. doi: 10.1007/s11250-014-0540-6.

134. Bett B, Lindahl J, Sang R, Wainaina M, Kairu-Wanyoike S, Bukachi S, et al. Association between Rift Valley fever virus seroprevalences in livestock and humans and their respective intra-cluster correlation coefficients, Tana River County, Kenya. Epidemiology and Infection. 2019;147. doi: 10.1017/S0950268818003242.

135. Nicolas G, Chevalier V, Tantely LM, Fontenille D, Durand B. A spatially explicit metapopulation model and cattle trade analysis suggests key determinants for the recurrent circulation of Rift Valley fever virus in a pilot area of Madagascar highlands. PLoS Negl Trop Dis. 2014;8(12):e3346. doi: 10.1371/journal.pntd.0003346.

136. Metras R, Cavalerie L, Dommergues L, Merot P, Edmunds WJ, Keeling MJ, et al. The epidemiology of Rift Valley fever in Mayotte: insights and perspectives from 11 years of data. Plos Neglected Tropical Diseases. 2016;10(6). doi: 10.1371/journal.pntd.0004783.

137. Alhaj M. Surveillance study on Rift Valley fever in Jazan region, Saudi Arabia. Int J Adv Sci Tech Res. 2015;5(4):1–13.

138. Ngoshe YB, Avenant A, Rostal MK, Karesh WB, Paweska JT, Bagge W, et al. Patterns of Rift Valley fever virus seropositivity in domestic ruminants in central South Africa four years after a large outbreak. Scientific Reports. 2020;10(1). doi: 10.1038/s41598-020-62453-6.

139. Moiane B, Mapaco L, Thompson P, Berg M, Albihn A, Fafetine J. High seroprevalence of Rift Valley fever phlebovirus in domestic ruminants and African Buffaloes in Mozambique shows need for intensified surveillance. Infect Ecol Epidemiol. 2017;7(1):1416248. Epub 2018/01/13. doi: 10.1080/20008686.2017.1416248.

140. Caron A, Cornelis D, Foggin C, Hofmeyr M, de Garine-Wichatitsky M. African Buffalo movement and zoonotic disease risk across transfrontier conservation areas, Southern Africa. Emerg Infect Dis. 2016;22(2):277–280. Epub 2016/01/27. doi: 10.3201/eid2202.140864.

141. Byomi AM, Samaha HA, Zidan SA, Hadad GA. Some associated risk factors with the occurence of Rift Valley fever in animals and man in certain localities of Nile delta, Egypt. Assiut Veterinary Medical Journal. 2015;61:10–17.

142. Mohamed AM, Ashshi AM, Asghar AH, Abd El-Rahim IH, El-Shemi AG, Zafar T. Seroepidemiological survey on Rift Valley fever among small ruminants and their close human contacts in Makkah, Saudi Arabia, in 2011. Rev Sci Tech. 2014;33(3):903–915. doi: 10.20506/rst.33.3.2328.

143. Wensman JJ, Lindahl J, Wachtmeister N, Torsson E, Gwakisa P, Kasanga C, et al. A study of Rift Valley fever virus in Morogoro and Arusha regions of Tanzania - serology and farmers’ perceptions. Infect Ecol Epidemiol. 2015;5:30025. doi: 10.3402/iee.v5.30025.

144. Elfadil AA, Hasab-Allah KA, Dafa-Allah OM. Factors associated with Rift Valley fever in south-west Saudi Arabia. Rev Sci Tech. 2006;25(3):1137–1145.

145. Mdlulwa Z, Ngwane CB. Evaluating the impact of 2010 Rift Valley fever outbreaks on sheep numbers in three provinces of South Africa. . African Journal of Agricultural Research. 2017;12(11):979–986.

146. Mroz C, Gwida M, El-Ashker M, Ziegler U, Homeier-Bachmann T, Eiden M, et al. Rift Valley fever virus infections in Egyptian cattle and their prevention. Transbound Emerg Dis. 2017;64(6):2049–2058. Epub 2017/01/25. doi: 10.1111/tbed.12616.

147. Abbas B, Yousif MA, M. NH. Animal health constraints to livestock exports from the Horn of Africa. Rev Sci Tech. 2014;33(3): 711–721.

148. Mohamed AM, Ghazi H, Ashshi AM, Faidah HS, Clinical EIA. Serological survey of Rift Valley fever among sacrifice animals in holy Mecca during pilgrimage season. . Int J Trop Med. 2011;6(4):85–89.

149. Tong C, Javelle E, Grard G, Dia A, Lacrosse C, Fourié T, et al. Tracking Rift Valley fever: From Mali to Europe and other countries, 2016. Euro Surveill. 2019;24(8). Epub 2019/02/28. doi: 10.2807/1560-7917.es.2019.24.8.1800213.

150. Liu B, Ma J, Jiao Z, Gao X, Xiao J, Wang H. Risk assessment for the Rift Valley fever occurrence in China: Special concern in south-west border areas. Transbound Emerg Dis. 2020. Epub 2020/06/23. doi: 10.1111/tbed.13695.

151. Grossi-Soyster EN, LaBeaud AD. Rift Valley fever: Important considerations for risk mitigation and future outbreaks. Trop Med Infect Dis. 2020;5(2). doi: 10.3390/tropicalmed5020089.

152. O’Hearn AE, Voorhees MA, Fetterer DP, Wauquier N, Coomber MR, Bangura J, et al. Serosurveillance of viral pathogens circulating in West Africa. Virol J. 2016;13(1):163. Epub 2016/10/08. doi: 10.1186/s12985-016-0621-4.

153. LaBeaud AD, Ochiai Y, Peters CJ, Muchiri EM, King CH. Spectrum of Rift Valley fever virus transmission in Kenya: Insights from three distinct regions. American Journal of Tropical Medicine and Hygiene. 2007;76(5):795–800. doi: 10.4269/ajtmh.2007.76.795.

154. Grossi-Soyster EN, Banda T, Teng CY, Muchiri EM, Mungai PL, Mutuku FM, et al. Rift Valley fever seroprevalence in coastal Kenya. Am J Trop Med Hyg. 2017;97(1):115–120. Epub 2017/07/19. doi: 10.4269/ajtmh.17-0104.

155. Cook EAJ, Grossi-Soyster EN, de Glanville WA, Thomas LF, Kariuki S, Bronsvoort BMD, et al. The sero-epidemiology of Rift Valley fever in people in the Lake Victoria Basin of western Kenya. Plos Neglected Tropical Diseases. 2017;11(7). doi: 10.1371/journal.pntd.0005731.

156. Gray GC, Anderson BD, LaBeaud AD, Heraud JM, Fevre EM, Andriamandimby SF, et al. Seroepidemiological study of interepidemic Rift Valley fever virus infection among persons with intense ruminant exposure in Madagascar and Kenya. Am J Trop Med Hyg. 2015;93(6):1364–1370. doi: 10.4269/ajtmh.15-0383.

157. LaBeaud AD, Bashir F, King CH. Measuring the burden of arboviral diseases: the spectrum of morbidity and mortality from four prevalent infections. Population Health Metrics. 2011;9. doi: 10.1186/1478-7954-9-1.

158. Muiruri S, Kabiru EW, Muchiri EM, Hussein H, Kagondu F, LaBeaud AD, et al. Cross-sectional survey of Rift Valley fever virus exposure in Bodhei village located in a transitional coastal forest habitat in Lamu County, Kenya. Am J Trop Med Hyg. 2015;92(2):394–400. Epub 2014/12/24. doi: 10.4269/ajtmh.14-0440.

159. Nanyingi MO, Munyua P, Kiama SG, Muchemi GM, Thumbi SM, Bitek AO, et al. A systematic review of Rift Valley fever epidemiology 1931-2014. Infect Ecol Epidemiol. 2015;5:28024. Epub 2015/08/04. doi: 10.3402/iee.v5.28024.

160. Jori F, Godfroid J, Michel AL, Potts AD, Jaumally MR, Sauzier J, et al. An assessment of zoonotic and production limiting pathogens in rusa deer (*Cervus timorensis rusa*) from Mauritius. Transbound Emerg Dis. 2014;61 Suppl 1:31–42. doi: 10.1111/tbed.12206.

161. Elfving K, Shakely D, Andersson M, Baltzell K, Ali AS, Bachelard M, et al. Acute uncomplicated febrile illness in children aged 2-59 months in Zanzibar - aetiologies, antibiotic treatment and outcome. PLoS One. 2016;11(1):e0146054. Epub 2016/01/29. doi: 10.1371/journal.pone.0146054.

162. Nicolas G, Durand B, Duboz R, Rakotondravao R, Chevalier V. Description and analysis of the cattle trade network in the Madagascar highlands: potential role in the diffusion of Rift Valley fever virus. Acta Trop. 2013;126(1):19–27. doi: 10.1016/j.actatropica.2012.12.013.

163. Abdo-Salem S, Tran A, Grosbois V, Gerbier G, Al-Qadasi M, Saeed K, et al. Can environmental and socioeconomic factors explain the recent emergence of Rift Valley fever in Yemen, 2000-2001? Vector Borne Zoonotic Dis. 2011;11(6):773–779. Epub 2011/02/03. doi: 10.1089/vbz.2010.0084.

164. Xiao Y, Beier JC, Cantrell RS, Cosner C, DeAngelis DL, Ruan S. Modelling the effects of seasonality and socioeconomic impact on the transmission of Rift Valley fever virus. PLoS Negl Trop Dis. 2015;9(1):e3388. Epub 2015/01/09. doi: 10.1371/journal.pntd.0003388.

165. Apolloni A, Nicolas G, Coste C, El Mamy AB, Yahya B, El Arbi AS, et al. Towards the description of livestock mobility in Sahelian Africa: Some results from a survey in Mauritania. PLoS One. 2018;13(1). doi: 10.1371/journal.pone.0191565.

166. Bergren NA, Borland EM, Hartman DA, Kading RC. Laboratory demonstration of the vertical transmission of Rift Valley fever virus by *Culex tarsalis* mosquitoes. PLoS Negl Trop Dis. 2021;15(3):e0009273. doi: 10.1371/journal.pntd.0009273.

167. Antonis AF, Kortekaas J, Kant J, Vloet RP, Vogel-Brink A, Stockhofe N, et al. Vertical transmission of Rift Valley fever virus without detectable maternal viremia. Vector Borne Zoonotic Dis. 2013;13(8):601–606. doi: 10.1089/vbz.2012.1160.

168. Adam I, Karsany MS. Case report: Rift Valley fever with vertical transmission in a pregnant Sudanese woman. J Med Virol. 2008;80(5):929. Epub 2008/03/25. doi: 10.1002/jmv.21132.

169. Arishi HM, Aqeel AY, Al Hazmi MM. Vertical transmission of fatal Rift Valley fever in a newborn. Ann Trop Paediatr. 2006;26(3):251–253. doi: 10.1179/146532806X120363.

170. Baudin M, Jumaa AM, Jomma HJE, Karsany MS, Bucht G, Näslund J, et al. Association of Rift Valley fever virus infection with miscarriage in Sudanese women: a cross-sectional study. Lancet Glob Health. 2016;4(11):e864–e871. Epub 2016/10/22. doi: 10.1016/s2214-109x(16)30176-0.

171. Pienaar NJ, Thompson PN. Temporal and spatial history of Rift Valley fever in South Africa: 1950 to 2011. Onderstepoort J Vet Res. 2013;80(1):384. Epub 2013/05/31. doi: 10.4102/ojvr.v80i1.384.

172. Ndengu M, Matope G, Tivapasi M, Pfukenyi DM, Cetre-Sossah C, De Garine-Wichatitsky M. Seroprevalence and associated risk factors of Rift Valley fever in cattle and selected wildlife species at the livestock/wildlife interface areas of Gonarezhou National Park, Zimbabwe. Onderstepoort J Vet Res. 2020;87(1):e1–e7. Epub 2020/05/07. doi: 10.4102/ojvr.v87i1.1731.

173. Nyakarahuka L, de St Maurice A, Purpura L, Ervin E, Balinandi S, Tumusiime A, et al. Prevalence and risk factors of Rift Valley fever in humans and animals from Kabale district in Southwestern Uganda, 2016. PLoS Negl Trop Dis. 2018;12(5):e0006412. Epub 2018/05/04. doi: 10.1371/journal.pntd.0006412.

174. FAO. Rift Valley fever vaccine development, progress and constraints. Proceedings of the GF-TADs meeting, January 2011. Rome, Italy: 2011.

175. Bird BH, Nichol ST. Breaking the chain: Rift Valley fever virus control via livestock vaccination. Curr Opin Virol. 2012;2(3):315–323. Epub 2012/04/03. doi: 10.1016/j.coviro.2012.02.017.

176. Shoemaker TR, Nyakarahuka L, Balinandi S, Ojwang J, Tumusiime A, Mulei S, et al. First laboratory-confirmed outbreak of human and animal Rift Valley fever virus in Uganda in 48 years. Am J Trop Med Hyg. 2019;100(3):659–671. Epub 2019/01/25. doi: 10.4269/ajtmh.18-0732.

177. Archer BN, Weyer J, Paweska J, Nkosi D, Leman P, Tint KS, et al. Outbreak of Rift Valley fever affecting veterinarians and farmers in South Africa, 2008. S Afr Med J. 2011;101(4):263–266. Epub 2011/07/27. doi: 10.7196/samj.4544.

178. Tambo E, Olalubi OA, Sacko M. Rift Valley fever epidemic in Niger near border with Mali. Lancet Infectious Diseases. 2016;16(12):1319–1320. doi: 10.1016/S1473-3099(16)30477-7.

179. Davies FG. Risk of a Rift Valley fever epidemic at the Haj in Mecca, Saudi Arabia. Rev Sci Tech. 2006;25(1):137–147. doi: 10.20506/rst.25.1.1648.

180. Krystosik A, Njoroge G, Odhiambo L, Forsyth JE, Mutuku F, LaBeaud AD. Solid wastes provide breeding sites, burrows, and food for biological disease vectors, and urban zoonotic reservoirs: A call to action for solutions-based research. Front Public Health. 2019;7:405. doi: 10.3389/fpubh.2019.00405.

181. Salmon-Rousseau A, Piednoir E, Cattoir V, de La Blanchardiere A. Hajj-associated infections. Medecine Et Maladies Infectieuses. 2016.

182. Cecilia H, Métras R, Fall AG, Lo MM, Lancelot R, Ezanno P. It’s risky to wander in September: Modelling the epidemic potential of Rift Valley fever in a Sahelian setting. Epidemics. 2020;33:100409. Epub 2020/11/03. doi: 10.1016/j.epidem.2020.100409.

183. Obiero JPO, Onyando JO. Climate. Developments in Earth Surface Processes. 162013. p. 39–50.

184. Alhaji NB, Aminu J, Lawan MK, Babalobi OO, Ghali-Mohammed I, Odetokun IA. Seropositivity and associated intrinsic and extrinsic factors for Rift Valley fever virus occurrence in pastoral herds of Nigeria: a cross sectional survey. BMC Vet Res. 2020;16(1):243. Epub 2020/07/16. doi: 10.1186/s12917-020-02455-8.

185. Durand B, Lo Modou M, Tran A, Ba A, Sow F, Belkhiria J, et al. Rift Valley fever in northern Senegal: A modelling approach to analyse the processes underlying virus circulation recurrence. Plos Neglected Tropical Diseases. 2020;14(6). doi: 10.1371/journal.pntd.0008009.

186. Kakani S, LaBeaud AD, King CH. Planning for Rift Valley fever virus: use of geographical information systems to estimate the human health threat of white-tailed deer (*Odocoileus virginianus*)-related transmission. Geospat Health. 2010;5(1):33–43. doi: 10.4081/gh.2010.185.

187. Wilson WC, Kim IJ, Trujillo JD, Sunwoo SY, Noronha LE, Urbaniak K, et al. Susceptibility of white-tailed deer to Rift Valley fever virus. Emerg Infect Dis. 2018;24(9):1717–1719. Epub 2018/08/21. doi: 10.3201/eid2409.180265.

188. Bron GM, Strimbu K, Cecilia H, Lerch A, Moore SM, Tran Q, et al. Over 100 Years of Rift Valley fever: A patchwork of data on pathogen spread and spillover. Pathogens. 2021;10(6). doi: 10.3390/pathogens10060708.

189. Schwarz NG, Girmann M, Randriamampionona N, Bialonski A, Maus D, Krefis AC, et al. Seroprevalence of antibodies against chikungunya, dengue, and Rift Valley fever viruses after febrile illness outbreak, Madagascar. Emerg Infect Dis. 2012;18(11):1780–1786. doi: 10.3201/eid1811.111036.

190. Al-Qabati AG, Al-Afaleq AI. Cross-sectional, longitudinal and prospective epidemiological studies of Rift Valley fever in Al-Hasa Oasis, Saudi Arabia. Journal of Animal and Veterinary Advances. 2010;9(2):258–265.

191. Grossi-Soyster EN, Lee J, King CH, LaBeaud AD. The influence of raw milk exposures on Rift Valley fever virus transmission. PLoS Negl Trop Dis. 2019;13(3):e0007258. doi: 10.1371/journal.pntd.0007258.

192. Gerken KN, Migliore E, Malumbo S, Shaita KN, Agola G, Fabre E, et al. Seroprevalence of Rift Valley fever virus in urban Kenya: a potential public health burden hiding in plain sight. Am J Trop Med Hyg. 2021:(in press).

193. Cook EAJ, de Glanville WA, Thomas LF, Kariuki S, Bronsvoort B, Fevre EM. Working conditions and public health risks in slaughterhouses in western Kenya. BMC Public Health. 2017;17. doi: 10.1186/s12889-016-3923-y.

194. Ikegami T, Balogh A, Nishiyama S, Lokugamage N, Saito TB, Morrill JC, et al. Distinct virulence of Rift Valley fever phlebovirus strains from different genetic lineages in a mouse model. PLoS One. 2017;12(12):e0189250. Epub 2017/12/22. doi: 10.1371/journal.pone.0189250.

195. WHO. Factors that contributed to undetected spread of the Ebola virus and impeded rapid containment Geneva, Switzerland: World Health Organization; 2015 [cited 2021 9/7/2021].

196. de St Maurice A, Nyakarahuka L, Purpura L, Ervin E, Tumusiime A, Balinandi S, et al. Notes from the field: Rift Valley fever response - Kabale District, Uganda, March 2016. MMWR Morb Mortal Wkly Rep. 2016;65(43):1200–1201. Epub 2016/11/05. doi: 10.15585/mmwr.mm6543a5.

197. Hartley DM, Rinderknecht JL, Nipp TL, Clarke NP, Snowder GD, National Center for Foreign A, et al. Potential effects of Rift Valley fever in the United States. Emerg Infect Dis. 2011;17(8):e1. doi: 10.3201/eid1708.101088.

198. Brugere-Picoux J, Chomel B. Importation of tropical diseases to Europe via animals and animal products: risks and pathways. Bulletin De L Academie Nationale De Medecine. 2009;193(8):1805–1819. doi: 10.1016/S0001-4079(19)32415-X.

199. Cito F, Narcisi V, Danzetta ML, Iannetti S, Sabatino DD, Bruno R, et al. Analysis of surveillance systems in place in European Mediterranean countries for West Nile virus (WNV) and Rift Valley fever (RVF). Transbound Emerg Dis. 2013;60 Suppl 2:40–44. Epub 2014/03/05. doi: 10.1111/tbed.12124.

200. Nielsen SS, Alvarez J, Bicout DJ, Calistri P, Depner K, Drewe JA, et al. Rift Valley fever - epidemiological update and risk of introduction into Europe. EFSAJ. 2020;18(3):e06041. Epub 2020/10/07. doi: 10.2903/j.efsa.2020.6041.

201. Rolin AI, Berrang-Ford L, Kulkarni MA. The risk of Rift Valley fever virus introduction and establishment in the United States and European Union. Emerg Microbes Infect. 2013;2(12):e81. Epub 2013/12/01. doi: 10.1038/emi.2013.81.

202. Zeller H, Marrama L, Sudre B, Van Bortel W, Warns-Petit E. Mosquito-borne disease surveillance by the European Centre for Disease Prevention and Control. Clin Microbiol Infect. 2013;19(8):693–698. Epub 2013/04/24. doi: 10.1111/1469-0691.12230.

203. Lagerqvist N, Moiane B, Mapaco L, Fafetine J, Vene S, Falk KI. Antibodies against Rift Valley fever virus in cattle, Mozambique. Emerg Infect Dis. 2013;19(7):1177–1179. Epub 2013/06/15. doi: 10.3201/eid1907.130332.

204. Abakar MF, Nare NB, Schelling E, Hattendorf J, Alfaroukh IO, Zinsstag J. Seroprevalence of Rift Valley fever, Q Fever, and brucellosis in ruminants on the southeastern shore of Lake Chad. Vector-Borne and Zoonotic Diseases. 2014;14(10):757–762. doi: 10.1089/vbz.2014.1585.

205. Consultative Group for RVFDS. Decision-support tool for prevention and control of Rift Valley fever epizootics in the Greater Horn of Africa. Am J Trop Med Hyg. 2010;83(2 Suppl):75–85. doi: 10.4269/ajtmh.2010.83s2a03.

206. Shah MM, Ndenga BA, Mutuku FM, Vu DM, Grossi-Soyster EN, Okuta V, et al. High dengue burden and circulation of 4 virus serotypes among children with undifferentiated fever, Kenya, 2014-2017. Emerg Infect Dis. 2020;26(11):2638–2650. doi: 10.3201/eid2611.200960.

207. Vu DM, Mutai N, Heath CJ, Vulule JM, Mutuku FM, Ndenga BA, et al. Unrecognized dengue virus infections in children, western Kenya, 2014-2015. Emerg Infect Dis. 2017;23(11):1915–1917. doi: 10.3201/eid2311.170807.

208. Hortion J, Mutuku FM, Eyherabide AL, Vu DM, Boothroyd DB, Grossi-Soyster EN, et al. Acute flavivirus and alphavirus infections among children in two different areas of Kenya, 2015. Am J Trop Med Hyg. 2019;100(1):170–173. doi: 10.4269/ajtmh.18-0297.

209. Hertz JT, Munishi OM, Ooi EE, Howe S, Lim WY, Chow A, et al. Chikungunya and dengue fever among hospitalized febrile patients in northern Tanzania. Am J Trop Med Hyg. 2012;86(1):171–177. doi: 10.4269/ajtmh.2012.11-0393.

210. Pepin M, Bouloy M, Bird BH, Kemp A, Paweska J. Rift Valley fever virus (Bunyaviridae: Phlebovirus): an update on pathogenesis, molecular epidemiology, vectors, diagnostics and prevention. Vet Res. 2010;41(6):61. Epub 2010/12/29. doi: 10.1051/vetres/2010033.

211. Fakour S, Naserabadi S, Ahmadi E. The first positive serological study on Rift Valley fever in ruminants of Iran. J Vector Borne Dis. 2017;54(4):348–352. Epub 2018/02/21. doi: 10.4103/0972-9062.225840.

212. Amato L, Dente MG, Calistri P, Declich S. Integrated early warning surveillance: Achilles’ Heel of One Health? Microorganisms. 2020;8(1). doi: 10.3390/microorganisms8010084.

213. Hassan OA, Ahlm C, Evander M. A need for One Health approach – lessons learned from outbreaks of Rift Valley fever in Saudi Arabia and Sudan. Infect Ecol Epidemiol. 2014;4. Epub 2014/02/08. doi: 10.3402/iee.v4.20710.

