## Supplementary material for "Paving the way for human vaccination against Rift Valley fever virus: A systematic literature review of RVFV epidemiology from 1999 to 2021": S2_fig Impact of sheltering animals

### Odds Ratio of RVFV Infection via Sheltering Livestock

| Study name | Period | Time Interval | Odds ratio | Lower limit | Upper limit | p-Value |
|---|---|---|---|---|---|---|
| LaBeaud (2008) | IEP | 2006 | 6.040 | 1.400 | 26.049 | 0.016 |
| Anyangu (2010) | Outbreak | 2007 | 1.945 | 1.230 | 3.076 | 0.004 |
| Al-Azraqi (2012) | IEP | 2008 | 1.961 | 1.073 | 3.582 | 0.028 |
| LaBeaud (2011) | IEP | 2009 | 3.810 | 1.544 | 9.403 | 0.004 |
| Cook (2017) | IEP | 2010-2012 | 4.993 | 1.087 | 22.924 | 0.039 |
| LaBeaud (2015) | IEP | 2011 | 6.100 | 3.593 | 10.357 | 0.000 |
| Memish (2015) | IEP | 2012 | 4.000 | 1.109 | 14.422 | 0.034 |
| **Pooled OR (random effects)** | | | 3.370 | 2.115 | 5.369 | 0.000 |

Odds ratio and 95% CI

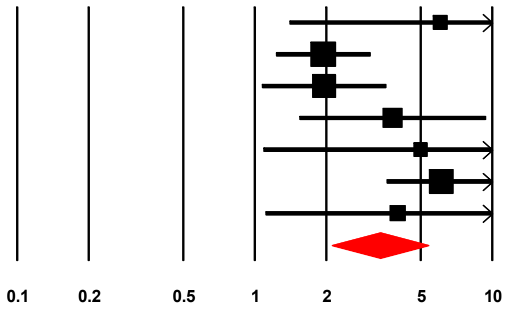


**Less likely** **More likely**
