## Supplementary material for "Paving the way for human vaccination against Rift Valley fever virus: A systematic literature review of RVFV epidemiology from 1999 to 2021": S3_fig impact of milking

### Odds Ratio of RVFV Infection via Milking Livestock

| Study name | Period | Time Interval | Odds ratio | Lower limit | Upper limit | p-Value |
|---|---|---|---|---|---|---|
| | | | Statistics for each study | | | |
| LaBeaud (2008) | IEP | 2006 | 12.550 | 3.711 | 42.447 | 0.000 |
| Muiruri (2015) | IEP | 2006 | 3.833 | 1.603 | 9.164 | 0.003 |
| Anyangu (2010) | Outbreak | 2007 | 8.039 | 5.289 | 12.218 | 0.000 |
| LaBeaud (2015) | IEP | 2011 | 4.920 | 2.972 | 8.145 | 0.000 |
| Sumaye (2015) | IEP | 2012 | 2.212 | 1.334 | 3.668 | 0.002 |
| **Pooled OR (random effects)** | | | 4.967 | 2.790 | 8.844 | 0.000 |

Odds ratio and 95%CI

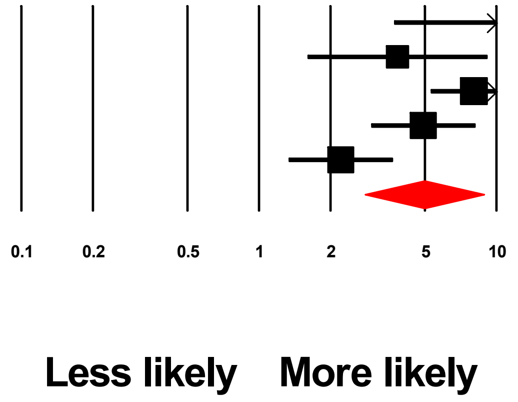
