## Supplementary material for "Paving the way for human vaccination against Rift Valley fever virus: A systematic literature review of RVFV epidemiology from 1999 to 2021": S7_Table Insect studies for RVFV

| **Summary of Vector Study Results from Included Studies 1999-2021 Grouped by Country** | | | | |
| --- | --- | --- | --- | --- |
| **Epidemic period** | **Country, study year** | **Vector Testing Results Summary** | **RVFV-affected species detected** | **Reference** |
| **Interepidemic** | Botswana 2011 and 2012 | A total of 18,259 mosquitoes divided in mosquito pools were processed to obtain supernatant fluid, which was inoculated in infant mice. Those mice were tested by RT-PCR and were all negative |  | [1] |
| **Interepidemic** | Egypt 2004-2005 | Mosquitoes tested in monthly pools. Kafr El Sheikh had positive pools every month from Jan to Nov 2004. El Beheira had positive pools in Jun Aug Sept 2004 | Domestic animals, Insects | [2] |
| **Active Epidemic/Epizootic** | Kenya 2006-2007 | RVFV positive *Ae. ochraceus* had fed on human (1) cattle (1), sheep(4), goats (3), and an unidentified host (1). Disseminated RVFV infections, as indicated by presence of virus in the insect heads, were observed in mosquitoes with blood meals from sheep (3), a goat, and one unidentified host. A single RVFV disseminated infection was detected in *Ae. mcintoshi,* which had fed on a donkey. In Baringo RVFV-positive *Ma. uniformis* had fed on sheep (4), a goat, and an unidentified host. | Domestic animals, Insects | [3] |
| **Active Epidemic/Epizootic** | Kenya 2007 | Rift Valley fever virus was detected in 77 of the 3,003 mosquito pools tested by RT-PCR. The virus was detected in mosquitoes from Garissa, Kilifi, and Baringo districts but not from Kirinyaga district. Each district was found to contain a unique set of RVFV-infected mosquito species | Insects | [4] |
| **Interepidemic** | Korea 2012 and 2013 | In 2012, the most frequently collected species were *Culex* (Cx.) *pipiens* (79.2%), followed by *Aedes*( Ae.) *vexans* (12.6%), *Ae. albopictus* (2.7%),*Cx. tritaeniorhynchus* (2.2%), and *Cx. inatomii* (2.2%). In 2013, the most frequently collected species was *Cx. pipiens* (55.9%), followed by *Ae. vexans* (12.1%), *Anopheles* (An.) *sinensis* (11.6%), *Ae. albopictus* (8.7%), and *Cx. tritaeniorhynchus* (4.3%). All 1827 pools were negative by RT-PCR. |  | [5] |
| **Active Epidemic/Epizootic** | Madagascar 2008-2009 | Tested a total of 319 pools for RVFV, including 59, 126, and 134 pools from Fianarantsoa I, Fianarantsoa II, and Ambalavao, respectively. RVFV RNA was detected in eight pools: two pools of *Cx. antennatus* from Fianarantsoa I, one pool of *An. coustani* and two pools of *An. squamosus* from Fianarantsoa II, and three pools of *An. squamosus* from Ambalavao | Insects | [6] |
| **Interepidemic** | Madagascar 2016 | Rift Valley fever virus detected in 17/704 tested in just three *Culex* species: *Culex tritaeniorhynchus*, *Culex antennatus* and *Culex decens* | Insects | [7] |
| **Active Epidemic/Epizootic** | Mauritania 2003 | *Culex poicilipes* was the most frequent species (43.8%), followed by *Cx. Antennatus* (23%) and Mansonia *uniformis* (9%). A total of 544 monospecific pools were submitted for viral isolation and only *Cx. poicilipes* had RVFV. Three RVFV strains (ArD 174367, ArD 174303, and ArD 174347) were isolated from the 146 pools constituted in Guimi Province | Humans, Domestic animals, Insects | [8] |
| **Active Epidemic/Epizootic** | Mauritania 2012 | RVFV strains detected but vector work done after vector control campaign. *Aedes vexans*, *Culex poicilipes*, *Culex antennatus*, and *Mansonia uniformis* comprised 52.6% of the mosquitoes collected; | Humans, Insects | [9] |
| **Active Epidemic/Epizootic** | Mauritania, Senegal 1998-1999 | 7 positive pools from Mauritania in 1999. These results are the first field evidence of *Cx. poicilipes* naturally infected with RVFV, and the first isolations of this virus from mosquitoes in Mauritania | Insects | [10] |
| **Active Epidemic/Epizootic** | Niger 2016 | 181 pools of mosquitoes, sandflies and *Culicoides* were captured. The mosquitoes were mainly *Anopheles* and *Culex* species with *Aedes* species found only rarely. Analysis by RT-PCR did not reveal any positive results for RVFV. | Humans, Domestic animals | [11] |
| **Active Epidemic/Epizootic** | Saudi Arabia 2000 | A single pool yielded a virus isolate. Rt-PCR on M segment fragment | Insects | [12] |
| **Active Epidemic/Epizootic** | Saudi Arabia 2000 | Large numbers of 2 species of mosquitos, *Culex tritaeniorhynchus* and *Aedes caspius*, in Al Ardah district, the epicenter of the outbreak where the first human cases were reported. Preliminary laboratory studies have already yielded isolates of RVF virus from both of these species. | Humans, Domestic animals, Insects | [13] |
| **Active Epidemic/Epizootic** | Senegal 2012 | No RVFV RNA was detected from 519 mosquito pools sampled in the Kedougou region during October 2012, although these pools included 7 species previously found associated with RVFV and which represented 26.6 % of the pools | Human | [14] |
| **Interepidemic** | Senegal 2013 | A single strain of RVFV was isolated from a pool of *Ae. ochraceus* collected from a pond in September 2013. 110 pools also tested at this time. Minimum field infection rate was 0.8 | Insects | [15] |
| **Active Epidemic/Epizootic** | Senegal 2013 and 2014 | No RVFV strains were isolated from mosquitoes in Mbour, but 1 pool (*Aedes ochraceus*) of mosquitoes of 437 collected in the Linguere district tested positive by PCR and virus isolation in November 2013. | Humans, Domestic animals, Insects | [16] |
| **Active Epidemic/Epizootic** | Sudan 2008 and 2009 | Llat KuKu (Site D) had 300 *Culex* and were positive for RVFV with a CT value of 39; Soba West (Site E) had 177 *Aedes* positive for RVFV with a CT value of 33. *Culex* at site E were negative; Sites A-C were negative | Insects | [17] |
| **Interepidemic** | Democratic Republic of the Congo 2014 | RVFV positive 2 *Aedes* pools. 2922 mosquitoes were collected from the study sites, comprising 1986 *Aedes* spp, 631 *Culex* spp, 283 *Anopheles* spp, and 22 *Mansonia* spp. | Insects | [18] |
| **Active Epidemic/Epizootic** | Egypt 2003 | Three isolates of RVF virus (RVFV) were obtained from 297 tested pools of female mosquitoes and all three RVFV isolates came from *Cx. antennatus* | Humans, Domestic animals, Insects | [19] |
| **Interepidemic** | Egypt 2009-2010 | All negative for RVFV 872 mosquitoes were collected by using CDC light trap and identified into 32 mosquito pool according to genus *Culex*, *Aedes* and *Anopheles*, *Culex* constituted the higher percentages of 71.8%. | Humans, Domestic animals | [20] |
| **Active Epidemic/Epizootic** | Kenya, Tanzania, Somalia 2006-2007 | The Kenya 1 lineage viruses consisted of 2 sublineages and 1 mosquito isolate (KEN/Bar-Msq 187-09/07). The Kenya 1b sublineage consisted of 2 mosquito isolates | Humans, Domestic animals, Insects | [21] |
| **Active Epidemic/Epizootic** | Mauritania 2010 | A total of 2,741 mosquitoes belonging to 5 genera and 11 species were collected for entomologic investigation. *Culex antennatus* was the most abundant mosquito species. Three RVFV strains were isolated from *Cx. antennatus* mosquitoes collected in Safia | Humans, Domestic animals, Insects | [22] |
| **Active Epidemic/Epizootic** | Saudi Arabia 2000 | Among 23,699 mosquito females tested, isolations of RVF virus were made from six of 15,428 *Culex tritaeniorhynchus* and from seven of 8091 *Aedes vexans arabiensis*. | Insects | [23] |
| **Interepidemic** | Saudi Arabia 2016 | RVFV detected by PCR in *H. schulzei*, *Hyalomma onatoli*, and *H. dromedarii* ticks (594 total tested in groups) | Insects | [24] |
| **Active Epidemic/Epizootic** | Senegal 2002,2003 | In 2002 rainy season, primary species *Ae vexans* that had 10 different strains and 2nd most was *Culex poicilipes* which had 29 different RVF strains. In 2003 rainy season, Other species were captured more and no *Aedes vexans* tested positive but *Culex poicilipes* had second highest abundance and had 3 different RVF strains | Insects | [25] |
| **Active Epidemic/Epizootic** | Sudan 2007 | RVFV was successfully detected in larvae and females of *An. gambiae arabiensis*, *An. coustani*, *Cx. pipiens* complex and *Ae. aegypti* collected from White Nile state. RVFV was successfully detected in both larvae and females of *Cx. pipiens* complex and *Cx. poicilipes* collected from Khartoum stat | Humans, Insects | [26] |
| **Interepidemic** | Tanzania 2013 | All pools tested negative for RVFV by PCR | Domestic animals, Insects | [27] |
| **Interepidemic** | Tanzania 2015 | 480 blood fed Aedes (24 pools) tested for RVFV all were negative | Insects | [28] |
| **Interepidemic** | Turkey before 2019 | all mosquito and tick pools negative |  | [29] |
| **Active Epidemic/Epizootic** | Uganda 2016 | Only three (1%) mosquito pools were found positive for RVFV by RT-PCR. One positive pool was *A. gibbinsi* (1.4%) trapped in Mushenyi village, home of the first probable case, PC1. The second positive pool was unspecified *Aedes* spp. (12.5%) trapped in Kazigizigi, Southern division, Kabale town. The third positive pool was *C. fuscopennata* (2.5%) trapped near the home of the second acute case | Humans, Domestic animals, Insects | [30] |
| **Active Epidemic/Epizootic** | Union of Comoros 2010-2011 | No RVFV RNA was detected in any of the 442 pools of blood sucking insects | Domestic animals | [31] |

**Cited References:**

1. Pachka H, Annelise T, Alan K, Power T, Patrick K, Véronique C, et al. Rift Valley fever vector diversity and impact of meteorological and environmental factors on Culex pipiens dynamics in the Okavango Delta, Botswana. Parasit Vectors. 2016;9(1):434. Epub 2016/08/10. doi: 10.1186/s13071-016-1712-1.

2. Youssef HM, Ghoneim MA, al. e. Recent trends for diagnosis of Rift Valley Fever in animals and mosquitoes in Egypt with special reference to the carrier. Global Veterinaria. 2008;2(1).

3. Lutomiah J, Omondi D, Masiga D, Mutai C, Mireji PO, Ongus J, et al. Blood meal analysis and virus detection in blood-fed mosquitoes collected during the 2006-2007 Rift Valley fever outbreak in Kenya. Vector Borne Zoonotic Dis. 2014;14(9):656-664. doi: 10.1089/vbz.2013.1564.

4. Sang R, Kioko E, Lutomiah J, Warigia M, Ochieng C, O'Guinn M, et al. Rift Valley fever virus epidemic in Kenya, 2006/2007: the entomologic investigations. Am J Trop Med Hyg. 2010;83(2 Suppl):28-37. doi: 10.4269/ajtmh.2010.09-0319.

5. Kim HJ, Lyoo HR, Park JY, Choi JS, Lee JY, Jeoung HY, et al. Surveillance of Rift Valley Fever Virus in Mosquito Vectors of the Republic of Korea. Vector Borne Zoonotic Dis. 2016;16(2):131-135. Epub 2016/01/16. doi: 10.1089/vbz.2015.1843.

6. Ratovonjato J, Olive MM, Tantely LM, Andrianaivolambo L, Tata E, Razainirina J, et al. Detection, isolation, and genetic characterization of Rift Valley fever virus from Anopheles (Anopheles) coustani, Anopheles (Anopheles) squamosus, and Culex (Culex) antennatus of the Haute Matsiatra region, Madagascar. Vector Borne Zoonotic Dis. 2011;11(6):753-759. Epub 2010/10/30. doi: 10.1089/vbz.2010.0031.

7. Jeffries CL, Tantely LM, Raharimalala FN, Hurn E, Boyer S, Walker T. Diverse novel resident Wolbachia strains in Culicine mosquitoes from Madagascar. Scientific Reports. 2018;8. doi: 10.1038/s41598-018-35658-z.

8. Faye O, Diallo M, Diop D, Bezeid OE, Bâ H, Niang M, et al. Rift Valley fever outbreak with East-Central African virus lineage in Mauritania, 2003. Emerg Infect Dis. 2007;13(7):1016-1023. Epub 2008/01/25. doi: 10.3201/eid1307.061487.

9. Sow A, Faye O, Ba Y, Ba H, Diallo D, Faye O, et al. Rift Valley fever outbreak, southern Mauritania, 2012. Emerg Infect Dis. 2014;20(2):296-299. Epub 2014/01/23. doi: 10.3201/eid2002.131000.

10. Diallo M, Nabeth P, Ba K, Sall AA, Ba Y, Mondo M, et al. Mosquito vectors of the 1998-1999 outbreak of Rift Valley Fever and other arboviruses (Bagaza, Sanar, Wesselsbron and West Nile) in Mauritania and Senegal. Med Vet Entomol. 2005;19(2):119-126. Epub 2005/06/17. doi: 10.1111/j.0269-283X.2005.00564.x.

11. Lagare A, Fall G, Ibrahim A, Ousmane S, Sadio B, Abdoulaye M, et al. First occurrence of Rift Valley fever outbreak in Niger, 2016. Vet Med Sci. 2019;5(1):70-78. Epub 2018/11/10. doi: 10.1002/vms3.135.

12. Miller BR, Godsey MS, Crabtree MB, Savage HM, Al-Mazrao Y, Al-Jeffri MH, et al. Isolation and genetic characterization of Rift Valley fever virus from Aedes vexans arabiensis, Kingdom of Saudi Arabia. Emerg Infect Dis. 2002;8(12):1492-1494. Epub 2002/12/25. doi: 10.3201/eid0812.020194.

13. Rift Valley fever, Saudi Arabia, August-October 2000. Wkly Epidemiol Rec. 2000;75(46):370-371. Epub 2001/01/06.

14. Sow A, Faye O, Faye O, Diallo D, Sadio BD, Weaver SC, et al. Rift Valley fever in Kedougou, southeastern Senegal, 2012. Emerg Infect Dis. 2014;20(3):504-506. Epub 2014/02/26. doi: 10.3201/eid2003.131174.

15. Ndiaye EH, Diallo D, Fall G, Ba Y, Faye O, Dia I, et al. Arboviruses isolated from the Barkedji mosquito-based surveillance system, 2012-2013. BMC Infect Dis. 2018;18(1):642. Epub 2018/12/14. doi: 10.1186/s12879-018-3538-2.

16. Sow A, Faye O, Ba Y, Diallo D, Fall G, Faye O, et al. Widespread Rift Valley Fever Emergence in Senegal in 2013-2014. Open Forum Infect Dis. 2016;3(3):ofw149. Epub 2016/10/06. doi: 10.1093/ofid/ofw149.

17. Abdelgadir DM, Bashab HMM, Mohamed RAE, Abuelmaali SA. Risk Factor Analysis for Outbreak of Rift Valley Fever in Khartoum State of Sudan. Journal of Entomological Science. 2010;45(3):239-251. doi: 10.18474/0749-8004-45.3.239.

18. Mbanzulu KM, Mboera LEG, Luzolo FK, Wumba R, Misinzo G, Kimera SI. Mosquito-borne viral diseases in the Democratic Republic of the Congo: a review. Parasit Vectors. 2020;13(1):103. Epub 2020/02/28. doi: 10.1186/s13071-020-3985-7.

19. Hanafi HA, Fryauff DJ, Saad MD, Soliman AK, Mohareb EW, Medhat I, et al. Virus isolations and high population density implicate Culex antennatus (Becker) (Diptera: Culicidae) as a vector of Rift Valley Fever virus during an outbreak in the Nile Delta of Egypt. Acta Trop. 2011;119(2-3):119-124. Epub 2011/05/17. doi: 10.1016/j.actatropica.2011.04.018.

20. Byomi AM, Samaha HA, Zidan SA, Hadad GA. Some associated risk factors with the occurence of Rift Valley fever in animals and man in certain localities of Nile delta, Egypt. Assiut Veterinary Medical Journal. 2015;61:10-17.

21. Nderitu L, Lee JS, Omolo J, Omulo S, O'Guinn ML, Hightower A, et al. Sequential Rift Valley fever outbreaks in eastern Africa caused by multiple lineages of the virus. J Infect Dis. 2011;203(5):655-665. doi: 10.1093/infdis/jiq004.

22. Faye O, Ba H, Ba Y, Freire CC, Faye O, Ndiaye O, et al. Reemergence of Rift Valley fever, Mauritania, 2010. Emerg Infect Dis. 2014;20(2):300-303. Epub 2014/01/23. doi: 10.3201/eid2002.130996.

23. Jupp PG, Kemp A, Grobbelaar A, Lema P, Burt FJ, Alahmed AM, et al. The 2000 epidemic of Rift Valley fever in Saudi Arabia: mosquito vector studies. Med Vet Entomol. 2002;16(3):245-252. Epub 2002/09/24. doi: 10.1046/j.1365-2915.2002.00371.x.

24. Mohamed RAE, Mohamed N, Aleanizy FS, Alqahtani FY, Al Khalaf A, Al-Keridis LA. Investigation of hemorrhagic fever viruses inside wild populations of ticks: One of the pioneer studies in Saudi Arabia. Asian Pacific Journal of Tropical Disease. 2017;7:299-303.

25. Ba Y, Sall AA, Diallo D, Mondo M, Girault L, Dia I, et al. Re-emergence of Rift Valley fever virus in Barkedji (Senegal, West Africa) in 2002-2003: identification of new vectors and epidemiological implications. J Am Mosq Control Assoc. 2012;28(3):170-178. Epub 2013/07/10. doi: 10.2987/12-5725.1.

26. Seufi AM, Galal FH. Role of Culex and Anopheles mosquito species as potential vectors of rift valley fever virus in Sudan outbreak, 2007. BMC Infect Dis. 2010;10:65. Epub 2010/03/13. doi: 10.1186/1471-2334-10-65.

27. Mhina AD, Kasanga CJ, Sindato C, Karimuribo ED, Mboera LE. Rift Valley fever potential mosquito vectors and their infection status in Ngorongoro District in northern Tanzania Tanzan J Health Res. 2015;17(4):1-9.

28. Bisimwa NP, Angwenyi S, Kinimi E, Shayo M, Bwihangane BA, Kasanga CJ. Molecular detection of arboviruses in Aedes mosquitoes collected from Kyela district, Tanzania. Revue De Medecine Veterinaire. 2016;167(5-6):138-144.

29. Bilgin Z, Turan N, Cizmecigil UY, Altan E, Esatgil MU, Yilmaz A, et al. Investigation of Vector-Borne Viruses in Ticks, Mosquitos, and Ruminants in the Thrace District of Turkey. Vector Borne Zoonotic Dis. 2020;20(9):670-679. Epub 2020/05/14. doi: 10.1089/vbz.2019.2532.

30. Shoemaker TR, Nyakarahuka L, Balinandi S, Ojwang J, Tumusiime A, Mulei S, et al. First Laboratory-Confirmed Outbreak of Human and Animal Rift Valley Fever Virus in Uganda in 48 Years. Am J Trop Med Hyg. 2019;100(3):659-671. Epub 2019/01/25. doi: 10.4269/ajtmh.18-0732.

31. Roger M, Beral M, Licciardi S, Soulé M, Faharoudine A, Foray C, et al. Evidence for circulation of the rift valley fever virus among livestock in the union of Comoros. PLoS Negl Trop Dis. 2014;8(7):e3045. Epub 2014/08/01. doi: 10.1371/journal.pntd.0003045.
