## Supplementary material for "Paving the way for human vaccination against Rift Valley fever virus: A systematic literature review of RVFV epidemiology from 1999 to 2021": S8_Table Effect of human animal exposure

**Human Risk Factors Summary**

84 included papers assessed human risk factors either for RVFV exposure or acute infection. The most common factors assessed were occupation, gender, age, proximity to water sources, behaviors related to mosquitoes, and contact with animals.

We conducted a separate analysis of studies that assessed animal contact related risk factors. The following table summarizes risk factors commonly assessed for RVFV exposure.

| **Human Risk Factor** | **Number of Studies that Assessed** |
| --- | --- |
| Travel | 15 |
| Behaviors related to mosquito exposure | 29 |
| Proximity to water sources | 12 |
| Gender | 50 |
| Contact with a dead human | 1 |
| Education level | 5 |
| Wealth | 1 |

**Animal Exposure Risk factors for Human Disease assessed**

39 Studies assessed human risk factors related to animal exposures and 17 of those studies linked animal exposures to acute infections.

We assessed the following common animal exposures and determined how many studies had assessed for the risk factor and how many studies had found statistically significant results. The statistics included as significant were based on bivariate or multivariate analyses. Not all studies separate the species of animals within each exposure risk factor, so this has been reported here as “not subdivided.” Not subdividing the risk factors by species limited the ability to compare between studies. More studies that assessed risk factors related to sheep detected significant results when they assessed sheep exposure separately. Twenty-one other risk factors were found significant, with consumption and preparation of raw meat found significant in four studies. The percent of studies that found statistical significance does not represent the likelihood of that risk factor leading to human disease.

Overall, human contact with animals is measured in a variety of ways and is most commonly not subdivided by species.

| **Animal Related Risk Factor** | **# of studies/ # with significance (% of total)** | **# studies assessed by species** | **# of studies with significant results** |
| --- | --- | --- | --- |
| Milking | 14/4 (29%) | 9 not subdivided by species  4 sheep  2 camels | 2 not subdivided  2 sheep  1 camel |
| Assisting birth | 19/10 (53%) | 14 not subdivided  4 sheep  1 camel  2 goats | 8 not subdivided  2 sheep |
| Herding | 28/8 (29%) | 20 not subdivided by species  7 sheep  2 camels  4 goats  2 Cows/Cattle  1 Other | 6 not subdivided  2 sheep |
| Feeding and other general care | 38/6 (16%) | 30 not subdivided  5 sheep  4 goats  1 camel  3 Cows/Cattle | 4 not subdivided  1 sheep  1 cattle |
| Housing Animals Inside the home | 14/7 (50%) | 10 not subdivided  3 sheep  3 goats  3 Cows/Cattle | 4 not subdivided  3 sheep  3 goats  2 Cows/Cattle |
| Skinning Animal | 16/3 (19%) | 14 not subdivided  1 sheep  1 Cows/Cattle | 2 not subdivided  1 Cows/Cattle |
| Butchering an animal | 40/10 (25%) | 33 not subdivided  5 sheep  1 camel  3 goats  1 Other | 7 not subdivided  2 Sheep  2 goats  3 Cows/Cattle |
| Drinking raw milk | 22/6 (27%) | 16 not subdivided  1 sheep  2 camels  2 Cows/Sheep  1 Other | 4 not subdivided  1 sheep  1 camel  1 cows/cattle |
| Disposal of aborted fetus | 31/13 (42%) | 21 not subdivided  7 sheep  1 camels  4 goats  2 Cows/Cattle  1 Other | 9 not subdivided  2 sheep  2 goats  2 Cows/Cattle  1 Other |
| Other | 27/21 | 16 not subdivided  4 Sheep  2 goats  6 Cattle cows  1 Other (donkey) | 21** |
| **Owning goats, living in close proximity to goats in urban areas, transporting aborted animals, performing autopsy, giving injections and specimen collection, hunting wild animals, skinned and had a recent fever, handling raw meat, eating raw meat, cooking with meat, contact with raw milk, cared for sick animal, disposal of animal carcass, contact with cattle x3, contact with donkeys, exposure to dead animals, prepared meat for cooking, | | | |

**Factors Associated with Severe Human Disease**

Factors most commonly associated with severe RVFV disease include animal contact exposures and ecological features.

| **Country** | **Study Year** | **Risk Factor(s) for Severe Disease** | **Reference** |
| --- | --- | --- | --- |
| Kenya (Germany) | 2007 | Co-infection with hepatitis A | 1. Oltmann, 2008 |
| Kenya | 2011 | Sheltered livestock, disposed of livestock fetuses; older age, village, recent illness, and death of a family member | 2. LaBeaud, 2005 |
| Kenya | 2007 | 4-person case series: animal contact (75%), no animal contact (25%) | 3. Kahlon, 2010 |
| Kenya | 2006-2007 | Geographic Location | 4. Outbreak news, 2007 |
| Kenya | 2015-2016 | Rainfall, flooding, mosquito swarms | 5. Oyas, 2018 |
| Kenya | 2007 | Animal contact, herding, caring for animals during birthing, touching aborted animal fetus  Death: Consuming or handling products from sick animals | 6. Anyangu, 2010 |
| Sudan | 2007 | local NDVI variation, local muddy soil | 7. Bashir, 2019 |
| Sudan | 2007 | males, age 15-29 | 8. Seufi, 2010 |

1. Oltmann A, Kämper S, Staeck O, Schmidt-Chanasit J, Günther S, Berg T, et al. Fatal outcome of hepatitis A virus (HAV) infection in a traveler with incomplete HAV vaccination and evidence of Rift Valley fever virus infection. J Clin Microbiol. 2008;46(11):3850-3852. Epub 2008/09/05. doi: 10.1128/jcm.01102-08.
2. LaBeaud AD, Pfeil S, Muiruri S, Dahir S, Sutherland LJ, Traylor Z, et al. Factors associated with severe human Rift Valley fever in Sangailu, Garissa County, Kenya. PLoS Negl Trop Dis. 2015;9(3):e0003548. Epub 2015/03/13. doi: 10.1371/journal.pntd.0003548.
3. Kahlon SS, Peters CJ, Leduc J, Muchiri EM, Muiruri S, Njenga MK, et al. Severe Rift Valley fever may present with a characteristic clinical syndrome. Am J Trop Med Hyg. 2010;82(3):371-375. doi: 10.4269/ajtmh.2010.09-0669.
4. Outbreak news. Rift Valley fever, Kenya. Wkly Epidemiol Rec. 2007;82(3):17-18. Epub 2007/01/24.
5. Oyas H, Holmstrom L, Kemunto NP, Muturi M, Mwatondo A, Osoro E, et al. Enhanced surveillance for Rift Valley fever in livestock during El Niño rains and threat of RVF outbreak, Kenya, 2015-2016. PLoS Negl Trop Dis. 2018;12(4):e0006353. Epub 2018/04/27. doi: 10.1371/journal.pntd.0006353.
6. Anyangu AS, Gould LH, Sharif SK, Nguku PM, Omolo JO, Mutonga D, et al. Risk factors for severe Rift Valley fever infection in Kenya, 2007. Am J Trop Med Hyg. 2010;83(2 Suppl):14-21. doi: 10.4269/ajtmh.2010.09-0293.
7. Bashir RSE, Hassan OA. A One Health perspective to identify environmental factors that affect Rift Valley fever transmission in Gezira state, Central Sudan. Tropical Medicine and Health. 2019;47(1). doi: 10.1186/s41182-019-0178-1.
8. Seufi AM, Galal FH. Role of *Culex* and *Anopheles* mosquito species as potential vectors of Rift Valley fever virus in Sudan outbreak, 2007. BMC Infect Dis. 2010;10:65. Epub 2010/03/13. doi: 10.1186/1471-2334-10-65.
