## Supplementary material for "Paving the way for human vaccination against Rift Valley fever virus: A systematic literature review of RVFV epidemiology from 1999 to 2021": S9_Table effect of Human SES

**Socioeconomic Risk Factors**

Socioeconomic factors found statistically significant. Only 17 studies assessed socioeconomic factors beyond age and gender. The significant results are summarized below.

| **SES Factor** | **Country** | **Significant Results** | **Reference** |
| --- | --- | --- | --- |
| **Wealth** | Kenya | Most no primary school | 1. Ochieng, 2015 |
|  | Kenya | Seropositive less likely to own land or motor vehicle | 2. Grossi-Soyster, 2017 |
|  | Saudi Arabia | Electricity in home, housing animals in home | 3. Al-Azraqi,2012 |
| **Location of home** | Kenya | Rural village location | 4. LaBeaud, 2011 |
| **Education** | Niger | Primary school only 4.5%, secondary or more 1.8% | 5. Alhaji, 2020 |
|  | Kenya | No formal education 64%, Some primary school 24%, Secondary School 12% | 6. Hassan, 2020 |
|  | Djbouti | Household size and education | 7. Andayi, 2014 |
| **Ethnicity** | Djbouti | Ethnicity Arab OR 3.4 | 7. Andayi, 2014 |

1. *Ochieng et al 2015. Seroprevalence of Infections with Dengue, Rift Valley Fever and Chikungunya Viruses in Kenya, 2007. PLoS ONE. 2015; 10(7):e0132645. doi:10.1371/journal.pone.0132645
2. Grossi-Soyster EN, Banda T, Teng CY, Muchiri EM, Mungai PL, Mutuku FM, et al. Rift Valley fever seroprevalence in coastal Kenya. Am J Trop Med Hyg. 2017;97(1):115-120. Epub 2017/07/19. doi: 10.4269/ajtmh.17-0104.
3. *Al-Azraqi et al. Rift Valley Fever in Southwestern Saudi Arabia: a sero-epidemiological study seven years after the outbreak of 2000-2001. Acta Trop. 2012; 123(2):111-116. 10.1016/j.actatropica.2012.04.007
4. *LaBeaud et al. Postepidemic Analysis of Rift Valley Fever Virus Transmission in Northeastern Kenya: A Village Cohort Study. PLOS Neglected Tropical Diseases. 2011; 5(8):1-9. doi:10.1371/journal.pntd.0001265
5. Alhaji NB, Aminu J, Lawan MK, Babalobi OO, Ghali-Mohammed I, Odetokun IA. Seropositivity and associated intrinsic and extrinsic factors for Rift Valley fever virus occurrence in pastoral herds of Nigeria: a cross sectional survey. BMC Vet Res. 2020;16(1):243. Epub 2020/07/16. doi: 10.1186/s12917-020-02455-8.
6. Hassan A, Muturi M, Mwatondo A, Omolo J, Bett B, Gikundi S, et al. Epidemiological investigation of a Rift Valley fever outbreak in humans and livestock in Kenya, 2018. Am J Trop Med Hyg. 2020;103(4):1649-1655. Epub 2020/08/05. doi: 10.4269/ajtmh.20-0387.
7. Andayi F, Charrel RN, Kieffer A, Richet H, Pastorino B, Leparc-Goffart I, et al. A sero-epidemiological study of arboviral fevers in Djibouti, Horn of Africa. PLoS Negl Trop Dis. 2014;8(12):e3299. Epub 2014/12/17. doi: 10.1371/journal.pntd.0003299.
