## Supplementary material for "Paving the way for human vaccination against Rift Valley fever virus: A systematic literature review of RVFV epidemiology from 1999 to 2021": S12_Table Wildlife RVFV seroprevalence

| **Wildlife Seroprevalence Among Included Studies 1999-2021** | | | |
| --- | --- | --- | --- |
| **Country name, study year** | **Wild Animal reported seroprevalence** | **Ongoing Transmission detected? (Method)** | **Unique Study ID**  **(see S2 and S3 Tables)** |
| Senegal_2013 and 2014 | Dorcas gazellas 24.2% | Yes (IgM or PCR +) | 277 |
| South Africa_2000-2006 | African buffalo 20.9% | Yes (seroconversion during cohort observation) | 141 |
| South Africa_2008 | African buffalo 36.2% | Yes ('clinical signs' and abortion) | 142 |
| South Africa_1996-2007 (X-sectional) South Africa_2008-12 (Cohort) | Buffalo (cohort) 33.5% | Yes (q6 months surveillance) | 140 |
| Kenya_2006-2007 | Buffalo_15.4%; Giraffe_43%; others were 0% | Yes (IgM or PCR +) | 173 |
| Tanzania_2007 (livestock)_2002-2006 (wildlife) | Buffalo_41% (9/22); elephant_33.3% (1/3) | Yes (IgM+) | 377 |
| Namibia_2011 | Springbok_35%, wildebeest_24% | Yes (IgM or PCR+) | 209 |
| Zimbabwe_before 2017 | African Buffalo_11.7%; Kudu_0.0%; Impala_0.0% | No | 479 |
| Kenya_2005 | African buffalo_16%, elephant_6%, warthog_2.5%, black rhino_32.6%, zebra_1%, giraffe_0%, kongoni_0%, eland_0%, leopard_0%, lion_0%, Thomson's gazelle_87.5%, lesser kudu_50%, impala_62.5%, waterbuck_20% | No | 50 |
| Mozambique_2013-2014 | African buffalo_30.4% | No | 470 |
| Kenya_2008-2015 | Baboon_0%, Black rhino_38.6%, Buffalo_17.9%, Elephant_22.2%, Giraffe_0%, Warthog_8.8%, Wildebeest_4.8%, Zebra_0% | No | 355 |
| Mayotte_2016 | Brown lemur_0.0% | No | 424 |
| South Africa_2003-2004 | Buffalo (6.0%) | No | 256 |
| Kenya_2000-2006 | buffalo_1/82 = 1.2%; waterbuck_2/17 = 11.8%; gazelle_1/11 = 9.1%; eland_0/15 = 0%; giraffe_0/16 = 0%; kongoni_0/14 = 0% | No | 191A |
| Botswana_2010, 2011 | Buffalo_12.7% | No | 322 |
| Kenya_2008-2009 | Buffalo_12/192 = 6.3%; waterbuck_2/13 = 15.4%; eland_0/3 = 0%; giraffe_1/48 = 2.1%; warthog_3/96 = 3.1%; gerenuk_0/4 = 0%; lesser kudu_0/2 = 0% | No | 191C |
| Kenya_2007 | Buffalo_35/163 = 21.5%; waterbuck_2/11 = 18.1%; gazelle_0/27 = 0%; eland_1/4 = 25.0%; giraffe_2/15 = 13.3%; warthog_32/43 = 74.4%; gerenuk_4/6 = 75%; impala_2/2 = 100% | No | 191B |
| Zimbabwe_2007-2009 | Buffalo_5.3% | No | 210 |
| South Africa_2016-2018 | Nyala_33.6%, Impala_45.9% | No | 63 |
| Spain_2009-2015 | Red deer_0.0%; Fallow deer_0.0%; Mouflon_0.0% | No | 287 |
| South Africa, Namibia, Kenya_1987-1997 | Rhinoceros_0.0% | No | 286 |
| Senegal_1996-1998 | Rodents:_3.8% (11/290) | No | 288 |
| Mauritius_2007 | Rusa deer_0% | No | 318 |
| South Africa_1999-2016 | Warthogs_1.87% | No | 10 |
| South Africa_2007 | White Rhinoceros_49% | No | 423 |
