## Supplementary material for "Paving the way for human vaccination against Rift Valley fever virus: A systematic literature review of RVFV epidemiology from 1999 to 2021": S13_Table surveillance systems for RVF

| **Rift Valley Fever virus Surveillance Systems Identified During Literature Review 1999-2021** | | | | | | | |
| --- | --- | --- | --- | --- | --- | --- | --- |
| **Country** | **Surveillance System Identified^a^** | **Year** | **Active Cases Detected?** | **Species** | **Study Type** | **Unique Study ID (see S2 and S3 tables)** | **Trigger for response?** |
| South Africa | National notifiable or reportable disease | 1950-present | Yes | Animal | Passive Surveillance | ID443 |  |
| South Africa | RVFV BioPortal Daily web searches in five languages for RVFV news (not able to find) | 07/ 2008 – link not active | No; web link is no longer active | Human | Web searches | ID404 | South Africa hosting the World Cup |
| South Africa | Alerting healthcare centers of high-risk period and periodic site visits, promotional and enhanced case finding activities | 2008-2011 | Yes; 302 cases and 25 deaths; CFR 8% | Human (engaged veterinary authorities in case finding) | Hospital based alerting | ID108 | A cluster of sick vet and farm workers (13) |
| South Africa | Testing of culled animals for routine herd thinning |  | No, serology only but neutralizing Abs in KwaZulu-Natal antelope indicating recent circulation | Wild Animals- | Testing Culled Wild Animals | ID63 |  |
| South Africa | Dip tank surveillance for FMD | 2016-2018 | 1Yes, 17 (20 goats, 66 cattle). First time seroconversion in animals was validated with gold standard SNT | Cattle, Goats | Cross-sectional survey, Longitudinal | ID73 |  |
| Mauritania | Reporting to local authorities in Aoejeft area and testing done at Laboratoire National d’Elevage et de Recherches Vétérinaires, Dakar, Senegal | 10_2010-12_2010 | 205 IgG and IgM, 123 IgM | Human and Animal | Prospective Cohort | ID239 | Reactive |
| Mauritania | Multi-sector task force has been jointly established by the MOH and the Ministry of Rural Development to respond to this outbreak | November 2012 | Yes, 25 confirmed, 17 deaths | Human | Prospective Cohort, Outbreak investigation | ID115 |  |
| Kenya | Global Outbreak Alert and Response Network (GOARN) 11-member team from GOARN partner institutions, and WHO (Country Office, Regional Office for Africa, and Headquarters) supporting the Ministry of Health | 2006-2007 | Yes |  | Hospital based Outpatients, Hospital based Inpatients | ID492 |  |
| Kenya | Kenya Ministry of Health (MoH) established nation-wide surveillance for RVF, initially with intensified efforts in Northeastern Province. Integrated Disease Surveillance and Response (IDSR) | 2006-2007 | Yes | Humans | Serosurvey accompanying outbreak cases | ID446 |  |
| Kenya | US National Aeronautics and Space Administration, US Department of Agriculture, and the Global Emerging Infection Surveillance program of US Department of Defense, predicted a RVF outbreak | 2006-2007 | 126 Human IgM+ | Targeted livestock surveillance | Biobanked sampled |  | Reactive to Early Warning Sign |
| Kenya | Nationwide HIV surveillance samples | 2007 |  |  | Cross sectional | ID411 | After outbreak leveraged samples from HIV |
| Kenya | Detection of Rift Valley Fever Virus Interepidemic Activity in Some Hotspot Areas of Kenya by Sentinel Animal Surveillance, | 2009-2012 |  | Yes, detected interepidemic seroconversions | Sentinel herds | ID365 |  |
| Kenya | Surveillance enhanced during high risk period using farmer phone reports of syndromic events suspected to be RVF in livestock. | 2015-2016 | 362 suspects (deaths in young animals, abortions, bleeding)-no IgM confirmed | No laboratory confirmed | Prospective Cohort | ID449 | In response to early warning during El Nino year |
| Kenya | Kenya Meteorological Department alerted that NE regions received 3x expected annual rainfall, outbreak detected using animal health syndromic surveillance | 2018 | 22/84 tested samples; 30 lab confirmed cases, 7 human deaths | Livestock and Humans | Snowball sampling, door to door, key informants |  |  |
| Kenya | Risk-based decision support tool development | 2009-now |  | Humans, animals, vectors |  |  | Created as a debriefing of ‘06/’07 outbreak |
| Madagascar | Madagascar instituted national RVF surveillance for humans in 2007 | 2007-now | Trapped mosquitoes in districts with animal and human RVF in 2008 and 2009. Positive pools of Anopheles, Culex | Mosquitoes | Mosquito trapping in RVF hotspots | ID398 |  |
| Madagascar | Sentinel surveillance system for early outbreak detection in Madagascar alerted and increase of cases of fever with joint pain initially and then 3 months after the outbreak | 2010 | A retrospective serology study for other arbovirus in pregnant women 2-3 months after the peak of a CHIKV outbreak and | Pregnant humans | Retrospective cohort | ID161 | Recent report of RVFV infections in humans and animals |
| Madagascar | Fever Sentinel Surveillance Network of Madagascar |  | 1 case of rift in a large study on non-malaria febrile disease. 7 year old girl that had been clinically diagnosed with mumps | Humans | Cross-sectional survey | ID268 |  |
| Mayotte | Active surveillance: Serological surveys conducted every year in livestock (cattle, goats and sheep). Passive surveillance: Sudden deaths or abortions reported by farmers. | 2013-2015 | No acute cases detected, monitor serology over time: 122 IgG+ in 2013-14; 29 IgG+ in 2014-15 | Animals | Multiple cross-sectional surveys | ID457 |  |

*^a^Early Warning Alerts***.** A shared goal of most early warning systems is to increase the lead time for prediction of RVF events. In the past 20 years, major advances in predictive disease modeling have led to the development of early warning systems that can use remote and location-specific monitoring to predict when environmental conditions are favorable for an outbreak. These signals most often rely on measures of rainfall, temperature, changes in vegetation (normalized difference vegetation index, NDVI), and local population structure to forecast possible RVF events on regional, national or local scales [1-7]. This signal is then meant to trigger active surveillance and RVFV case identification. In 2008, a joint International Livestock Research Institute (ILRI) and Government of Kenya Division of Veterinary Services participatory assessment of the 2006-2007 Kenya outbreak found that although the FAO EMPRES early warning was issued in November 2006, the earliest cases in livestock had already occurred in mid-October 2006 in North Eastern Province [8].

Various levels of success have been reported in early warning systems. In Kenya in 2015 and 2016, an early warning system confirmed an El Niño year and surveillance efforts were augmented by initiating farmer phone reports of syndromic events in livestock [9]. This identified 362 suspect cases from young animal deaths, abortions, and bleeding; however, none of those cases were laboratory confirmed with IgM antibody testing. Three years later in 2018, the Kenya Meteorological Department alerted that the high-risk northeast had region received three times the expected annual rainfall, which again triggered farmer reporting but also included snowball sampling and testing strategies and door-to-door visits for humans [10]. These initiatives confirmed 22/84 livestock samples and 30 lab confirmed human cases for RVFV infections, including 7 human deaths.

*Detection in hospital-based surveys, including leverage of other disease surveillance systems to identify RVFV circulation***.** During 1999-2021, hospital-based serosurveys and/or PCR testing have identified unsuspected RVF cases through additional, non-routine testing after samples from febrile patients have tested negative for other arboviruses during local surveillance programs [11-17]. In Sierra Leone between 2006-2008, all samples sent to the national Kenema Government Hospital Lassa Diagnostic Laboratory that were Lassa virus- (LASV-) and malaria-negative were tested for RVFV [12, 18]. This identified 5 acute human cases retrospectively. These efforts confirmed that RVFV had been circulating acutely in the LASV hyperendemic area. Retesting of febrile patients enrolled in surveillance for dengue and chikungunya in Mayotte help to identify an acute outbreak of RVF in 2018-2019 [19]. In more pro-active surveillance system, 302 human cases were identified in South Africa between 2008-2011 by referral to local health facilities after regional site visit interviews screened for suspect case criteria. These included clinical signs consistent with RVFV infection as well as work exposure in high-risk occupations (having recent contact with livestock or game animals) [20].

*Use of sentinel herds for surveillance.* Sentinel herds assess real-time ongoing transmission in livestock and can provide insight for disease presence in surrounding herds while providing baseline data for more comprehensive field investigations. Sentinel herds are groups of livestock placed in areas considered to be high-risk, and animals are sampled intermittently to monitor for anti-RVFV seroconversions [16, 21-23]. Livestock that seroconvert during the interepidemic period do not always show clinical signs, which is concerning, as viremic adults may be exported or slaughtered for human consumption. Sentinel herds set up in three high-risk districts of Kenya were sampled every 4-6 weeks in 2009 up to June 2012 [24]. This study detected evidence of RVFV circulation during the interepidemic period of 2009 to 2012 in Ijara and Marigat. These sites had previously detected RVFV-affected livestock and humans during the 2006-2007 outbreak, yet, when interepidemic transmission was documented, there were no reports of concurrent human RVF cases, and no signs of increased abortion. This study also conducted ‘snowball’ target sampling near sentinel herds of animals with abortion, mortality in lambs, and other clinical signs of RVFV in livestock, which led to further case identification.

In Saudi Arabia, sentinel herds may have documented ongoing interepidemic transmission in 2003-2004, three years after the last overt outbreak. However, without confirmatory serologic testing (plaque reduction neutralization testing, PRNT) such results are not completely certain [25]. This study captured four cases of ongoing transmission based on animal IgM positivity (IgM) or seroconversion (IgG) during the study period. Again in Saudi Arabia in 2013, the use of sentinel herds allowed for detection of further ongoing low level transmission by identifying two additional acute cases [22]. Sentinel herds have also been used in response to reports of increase abortion incidence in Senegal in 2003, when sentinel herds were set up after farmers reported cases to the veterinary field services in collaboration with the National Veterinary Research Laboratory (ISRA-LNERV) and the Pasteur Institute of Dakar [21]. However, sentinel herds set up at each of the 12 veterinary posts that were involved in the previous Senegalese outbreak did not detect any acute cases, but they did find an existing anti-RVFV seroprevalence of 3% among study animals.

*Farmer reported syndromic surveillance.* The current widespread availability of basic cellular telephone service has provided an opportunity for farmers and veterinary services in more remote areas (without direct access to diagnostics) to report upticks in the occurrence of RVF-compatible animal syndromes. In Kenya, the use of m-Health helped detect the 2018 outbreak of RVF [9]. Although changes in abortion rates and young animal deaths over time may provide insight for RVFV circulation, there are many other infectious and non-infectious causes of livestock abortion in countries endemic for RVFV [26, 27].

Reliance on farmers to report clinical signs can be negatively affected by their economic disincentives to report RVF to authorities at all levels. In a qualitative community survey in Sudan, participants reported they were reluctant to report because they knew they would not receive financial compensation for their livestock, and they believed it was the responsibility of human health authorities to control RVFV rather than veterinary authorities [28].

1. Anyamba A, Chretien JP, Formenty PB, Small J, Tucker CJ, Malone JL, et al. Rift Valley Fever potential, Arabian Peninsula. Emerg Infect Dis. 2006;12(3):518-520. Epub 2006/05/23.

2. Anyamba A, Linthicum KJ, Small J, Britch SC, Pak E, de La Rocque S, et al. Prediction, assessment of the Rift Valley fever activity in East and Southern Africa 2006-2008 and possible vector control strategies. Am J Trop Med Hyg. 2010;83(2 Suppl):43-51. doi: 10.4269/ajtmh.2010.09-0289.

3. Anyamba A, Linthicum KJ, Small J, Britch SC, Tucker CJ. Remote sensing contributions to prediction and risk assessment of natural disasters caused by large-scale Rift Valley fever outbreaks. Proceedings of the Ieee. 2012;100(10):2824-2834. doi: 10.1109/JPROC.2012.2194469.

4. Redding DW, Tiedt S, Lo Iacono G, Bett B, Jones KE. Spatial, seasonal and climatic predictive models of Rift Valley fever disease across Africa. Philos Trans R Soc Lond B Biol Sci. 2017;372(1725). Epub 2017/06/07. doi: 10.1098/rstb.2016.0165.

5. Hardcastle AN, Osborne JCP, Ramshaw RE, Hulland EN, Morgan JD, Miller-Petrie MK, et al. Informing Rift Valley fever preparedness by mapping seasonally varying environmental suitability. Int J Infect Dis. 2020;99:362-372. Epub 2020/08/02. doi: 10.1016/j.ijid.2020.07.043.

6. Caminade C, Ndione JA, Kebe CMF, Jones AE, Danuor S, Tay S, et al. Mapping Rift Valley fever and malaria risk over West Africa using climatic indicators. Atmospheric Science Letters. 2011;12(1):96-103. doi: 10.1002/asl.296.

7. Lacaux JP, Tourre YM, Vignolles C, Ndione JA, Lafaye M. Classification of ponds from high-spatial resolution remote sensing: Application to Rift Valley fever epidemics in Senegal. Remote Sensing of Environment. 2007;106(1):66-74. doi: 10.1016/j.rse.2006.07.012.

8. Consultative Group for RVFDS. Decision-support tool for prevention and control of Rift Valley fever epizootics in the Greater Horn of Africa. Am J Trop Med Hyg. 2010;83(2 Suppl):75-85. doi: 10.4269/ajtmh.2010.83s2a03.

9. Oyas H, Holmstrom L, Kemunto NP, Muturi M, Mwatondo A, Osoro E, et al. Enhanced surveillance for Rift Valley fever in livestock during El Niño rains and threat of RVF outbreak, Kenya, 2015-2016. PLoS Negl Trop Dis. 2018;12(4):e0006353. Epub 2018/04/27. doi: 10.1371/journal.pntd.0006353.

10. Hassan A, Muturi M, Mwatondo A, Omolo J, Bett B, Gikundi S, et al. Epidemiological investigation of a Rift Valley fever outbreak in humans and livestock in Kenya, 2018. Am J Trop Med Hyg. 2020;103(4):1649-1655. Epub 2020/08/05. doi: 10.4269/ajtmh.20-0387.

11. Dellagi K, Salez N, Maquart M, Larrieu S, Yssouf A, Silai R, et al. Serological evidence of contrasted exposure to arboviral infections between islands of the Union of Comoros (Indian Ocean). Plos Neglected Tropical Diseases. 2016;10(12). doi: 10.1371/journal.pntd.0004840.

12. O'Hearn AE, Voorhees MA, Fetterer DP, Wauquier N, Coomber MR, Bangura J, et al. Serosurveillance of viral pathogens circulating in West Africa. Virol J. 2016;13(1):163. Epub 2016/10/08. doi: 10.1186/s12985-016-0621-4.

13. LaBeaud AD, Ochiai Y, Peters CJ, Muchiri EM, King CH. Spectrum of Rift Valley fever virus transmission in Kenya: Insights from three distinct regions. American Journal of Tropical Medicine and Hygiene. 2007;76(5):795-800. doi: 10.4269/ajtmh.2007.76.795.

14. Grossi-Soyster EN, Banda T, Teng CY, Muchiri EM, Mungai PL, Mutuku FM, et al. Rift Valley fever seroprevalence in coastal Kenya. Am J Trop Med Hyg. 2017;97(1):115-120. Epub 2017/07/19. doi: 10.4269/ajtmh.17-0104.

15. Schwarz NG, Girmann M, Randriamampionona N, Bialonski A, Maus D, Krefis AC, et al. Seroprevalence of antibodies against Chikungunya, Dengue, and Rift Valley fever viruses after febrile illness outbreak, Madagascar. Emerg Infect Dis. 2012;18(11):1780-1786. doi: 10.3201/eid1811.111036.

16. Al-Qabati AG, Al-Afaleq AI. Cross-sectional, longitudinal and prospective epidemiological studies of Rift Valley fever in Al-Hasa Oasis, Saudi Arabia. Journal of Animal and Veterinary Advances. 2010;9(2):258-265.

17. Guillebaud J, Bernardson B, Randriambolamanantsoa TH, Randrianasolo L, Randriamampionona JL, Marino CA, et al. Study on causes of fever in primary healthcare center uncovers pathogens of public health concern in Madagascar. PLoS Negl Trop Dis. 2018;12(7):e0006642. Epub 2018/07/17. doi: 10.1371/journal.pntd.0006642.

18. Schoepp RJ, Rossi CA, Khan SH, Goba A, Fair JN. Undiagnosed acute viral febrile illnesses, Sierra Leone. Emerging Infectious Diseases. 2014;20(7):1176-1182. doi: 10.3201/eid2007.131265.

19. Youssouf H, Subiros M, Dennetiere G, Collet L, Dommergues L, Pauvert A, et al. Rift Valley fever outbreak, Mayotte, France, 2018-2019. Emerg Infect Dis. 2020;26(4):769-772. Epub 2020/03/19. doi: 10.3201/eid2604.191147.

20. Archer BN, Thomas J, Weyer J, Cengimbo A, Landoh DE, Jacobs C, et al. Epidemiologic Investigations into Outbreaks of Rift Valley Fever in Humans, South Africa, 2008-2011. Emerging Infectious Diseases. 2013;19(12):1918-1925. doi: 10.3201/eid1912.121527.

21. Chevalier V, Lancelot R, Thiongane Y, Sall B, Diaité A, Mondet B. Rift Valley fever in small ruminants, Senegal, 2003. Emerg Infect Dis. 2005;11(11):1693-1700. Epub 2005/12/02. doi: 10.3201/eid1111.050193.

22. Alhaj M. Surveillance study on Rift Valley fever in Jazan region, Saudi Arabia. Int JAdv Sci Tech Res. 2015;5(4):1-13.

23. Lancelot R. Animaux sentinelles en milieu tropical: vers un système intégré de surveillance. . Epidémiol Santé Anim. 2009;56:27-34.

24. Lichoti JK, Kihara A, Oriko AA, Okutoyi LA, Wauna JO, Tchouassi DP, et al. Detection of Rift Valley fever virus interepidemic activity in some hotspot areas of Kenya by sentinel animal surveillance, 2009-2012. Vet Med Int. 2014;2014:379010. doi: 10.1155/2014/379010.

25. Elfadil AA, Hasab-Allah KA, Dafa-Allah OM. Factors associated with Rift Valley fever in south-west Saudi Arabia. Rev Sci Tech. 2006;25(3):1137-1145.

26. Kanouté YB, Gragnon BG, Schindler C, Bonfoh B, Schelling E. Epidemiology of brucellosis, Q Fever and Rift Valley fever at the human and livestock interface in northern Côte d'Ivoire. Acta Trop. 2017;165:66-75. Epub 2016/02/24. doi: 10.1016/j.actatropica.2016.02.012.

27. Abakar MF, Nare NB, Schelling E, Hattendorf J, Alfaroukh IO, Zinsstag J. Seroprevalence of Rift Valley fever, Q Fever, and brucellosis in ruminants on the southeastern shore of Lake Chad. Vector-Borne and Zoonotic Diseases. 2014;14(10):757-762. doi: 10.1089/vbz.2014.1585.

28. Hassan OA, Affognon H, Rocklöv J, Mburu P, Sang R, Ahlm C, et al. The One Health approach to identify knowledge, attitudes and practices that affect community involvement in the control of Rift Valley fever outbreaks. PLoS Negl Trop Dis. 2017;11(2):e0005383. Epub 2017/02/17. doi: 10.1371/journal.pntd.0005383.
